# Report 46: Factors driving extensive spatial and temporal fluctuations in COVID-19 fatality rates in Brazilian hospitals

**DOI:** 10.1101/2021.11.01.21265731

**Authors:** Andrea Brizzi, Charles Whittaker, Luciana M. S. Servo, Iwona Hawryluk, Carlos A. Prete, William M. de Souza, Renato S. Aguiar, Leonardo J. T. Araujo, Leonardo S. Bastos, Alexandra Blenkinsop, Lewis F. Buss, Darlan Candido, Marcia C. Castro, Silvia F. Costa, Julio Croda, Andreza Aruska de Souza Santos, Christopher Dye, Seth Flaxman, Paula L. C. Fonseca, Victor E. V. Geddes, Bernardo Gutierrez, Philippe Lemey, Anna S. Levin, Thomas Mellan, Diego M. Bonfim, Xenia Miscouridou, Swapnil Mishra, Mélodie Monod, Filipe R. R. Moreira, Bruce Nelson, Rafael H. M. Pereira, Otavio Ranzani, Ricardo P. Schnekenberg, Elizaveta Semenova, Raphael Sonnabend, Renan P. Souza, Xiaoyue Xi, Ester C. Sabino, Nuno R. Faria, Samir Bhatt, Oliver Ratmann

**Affiliations:** Department of Mathematics, Imperial College London, London, United Kingdom; MRC Centre for Global Infectious Disease Analysis, Jameel Institute, School of Public Health, Imperial College London, United Kingdom; Institute for Applied Economic Research - IPEA, Brasília, Brazil; Departamento de Engenharia de Sistemas Eletrônicos, Escola Politécnica da Universidade de São Paulo, São Paulo, Brazil; World Reference Center for Emerging Viruses and Arboviruses and Department of Microbiology and Immunology, University of Texas Medical Branch, Galveston, TX, USA; Departamento de Genética, Ecologia e Evolução, Instituto de Ciências Biológicas, Universidade Federal de Minas Gerais, Belo Horizonte, Brazil; Instituto D’Or de Pesquisa e Ensino (IDOR), Rio de Janeiro, Brazil; Laboratory of Quantitative Pathology, Center of Pathology, Adolfo Lutz Institute, São Paulo, Brazil; Programa de Computação Científica, Fundação Oswaldo Cruz, Rio de Janeiro, Brazil; Departamento de Moléstias Infecciosas e Parasitárias e Instituto de Medicina Tropical da Faculdade de Medicina da Universidade de São Paulo, São Paulo, Brasil; Department of Zoology, University of Oxford, Oxford, United Kingdom; Department of Global Health and Population, Harvard T.H. Chan School of Public Health, Boston, United States; Department of Epidemiology of Microbial Diseases, Yale School of Public Health, New Haven, United States; Latin American Centre, University of Oxford, Oxford, United Kingdom; Department of Computer Science, University of Oxford, Oxford, United Kingdom; Department of Microbiology, Immunology and Transplantation, KU Leuven – University of Leuven, Leuven, Belgium; Department of Infectious Disease Epidemiology, Imperial College London, London, United Kingdom; Section of Epidemiology, School of Public Health, University of Copenhagen, Denmark, Copenhagen; Departamento de Genética, Instituto de Biologia, Universidade Federal do Rio de Janeiro, Rio de Janeiro, Brazil; Environmental Dynamics, INPA, National Institute for Amazon Research, Bairro Petropolis, Brazil; Barcelona Institute for Global Health, ISGlobal, Barcelona, Spain; Nuffield Department of Clinical Neurosciences, University of Oxford, Oxford, United Kingdom; Instituto de Medicina Tropical, Faculdade de Medicina da Universidade de São Paulo, São Paulo, Brazil; Section of Epidemiology, School of Public Health, University of Copenhagen

## Abstract

The SARS-CoV-2 Gamma variant spread rapidly across Brazil, causing substantial infection and death waves. We use individual-level patient records following hospitalisation with suspected or confirmed COVID-19 to document the extensive shocks in hospital fatality rates that followed Gamma’s spread across 14 state capitals, and in which more than half of hospitalised patients died over sustained time periods. We show that extensive fluctuations in COVID-19 in-hospital fatality rates also existed prior to Gamma’s detection, and were largely transient after Gamma’s detection, subsiding with hospital demand. Using a Bayesian fatality rate model, we find that the geographic and temporal fluctuations in Brazil’s COVID-19 in-hospital fatality rates are primarily associated with geographic inequities and shortages in healthcare capacity. We project that approximately half of Brazil’s COVID-19 deaths in hospitals could have been avoided without pre-pandemic geographic inequities and without pandemic healthcare pressure. Our results suggest that investments in healthcare resources, healthcare optimization, and pandemic preparedness are critical to minimize population wide mortality and morbidity caused by highly transmissible and deadly pathogens such as SARS-CoV-2, especially in low- and middle-income countries.

**Note:** The following manuscript has appeared as ‘Report 46 - Factors driving extensive spatial and temporal fluctuations in COVID-19 fatality rates in Brazilian hospitals’ at https://spiral.imperial.ac.uk:8443/handle/10044/1/91875.

**One sentence summary:** COVID-19 in-hospital fatality rates fluctuate dramatically in Brazil, and these fluctuations are primarily associated with geographic inequities and shortages in healthcare capacity.

## Introduction

Since late 2020, emerging variants of concern of severe acute respiratory syndrome coronavirus 2 (SARS-CoV-2) have led to substantial epidemic waves and marked increases in COVID-19 deaths [1, 2, 3, 4, 5]. In Brazil, the SARS-CoV-2 Gamma variant of concern (also known as P.1, 20J/501Y.V3, GR/501Y.V3, VOC-21JAN-02 or VOC-202101/02) was first detected in December 2020 in Manaus, Amazonas state, north Brazil [3, 6, 7]. Gamma is defined by 17 unique mutations, including the N501Y, E484K and K417N in the spike protein that have been associated with increased transmissibility and immune escape [3, 8, 9]. Three weeks after its detection, Gamma was the dominant lineage circulating in Manaus as measured by variant frequency [3], and by the end of March 2021, SARS-CoV-2 sequence data belonging to the Gamma variant were available from 14 of 27 Brazilian federal states (Figure 1A). Gamma’s rapid spread through the country was followed by death waves suggesting increased disease severity following infection with Gamma [10], but these data have not been examined in the context of extensive inequities in baseline development and healthcare capacity across Brazil, which are common in low- and middle-income countries [11]. Here, we use individual-level patient histories following hospitalisation with suspected or confirmed COVID-19 (Figure 2) [12, 13] to describe how Gamma’s expansion was followed by shocks in COVID-19 fatality rates in Brazilian hospitals, and show that in-hospital fatality rates also fluctuated extensively before Gamma’s emergence. We introduce pandemic healthcare pressure indices that measure and monitor mismatches between healthcare demand and available resources. We find that these pandemic healthcare pressure indices are strongly correlated with variations in COVID-19 fatality rates. Using a Bayesian model, we then assess the factors driving the extensive fluctuations in COVID-19 in-hospital fatality rates, from pre-pandemic geographic inequities in economic development, healthcare resources and vulnerable populations [14, 15, 16, 17, 18], to pandemic healthcare pressures [15, 19, 20], and variant-specific disease severity as measured by in-hospital fatality rates [21, 22].

**Figure 1:**
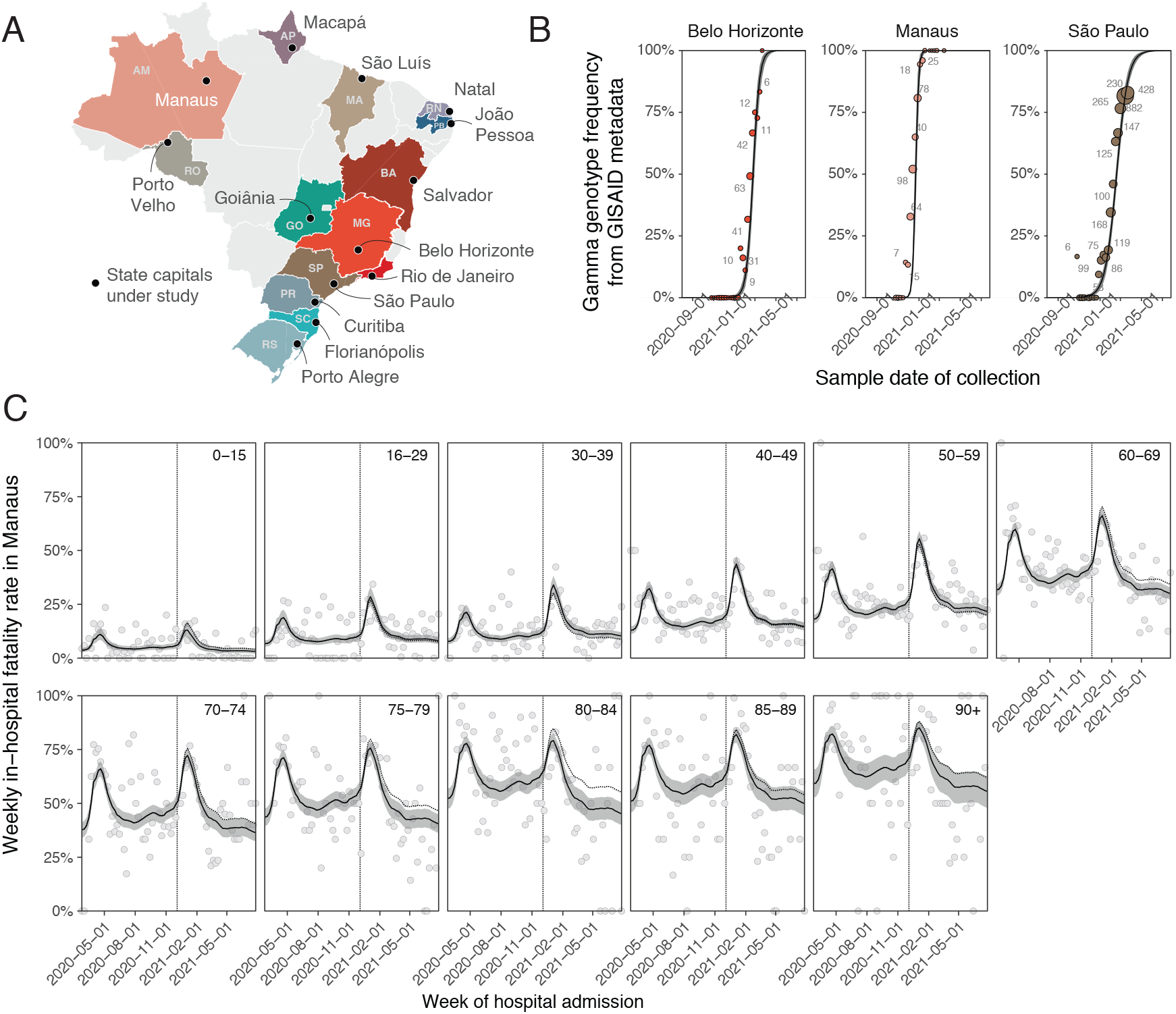
Spatio-temporal expansion of SARS-CoV-2 Gamma in Brazil and associated shocks in COVID-19 fatality rates in hospitals. (A) The 14 states and state capitals in which Gamma was detected by March 31, 2021 and which were included in the analysis. (B) Time evolution of SARS-CoV-2 Gamma variant frequencies in three locations, suggesting rapid expansion. Data from GISAID [23] (dots) are shown along with the number of sequenced SARS-CoV-2 samples (text), and posterior median model fits (line) and associated 95% credible intervals (CrI) (grey ribbon). (C) Weekly COVID-19 in-hospital fatality rates among hospitalised residents in Manaus with no evidence of vaccination prior to admission (dots), by age group (facets). Posterior median estimates (line) of the Bayesian multi-strain fatality model are shown along with 95% CrIs (grey ribbon), and the expected in-hospital fatality rates of non-Gamma variants (dotted line). The date of Gamma’s first detection in each location is indicated as a vertical dotted line.

**Figure 2:**
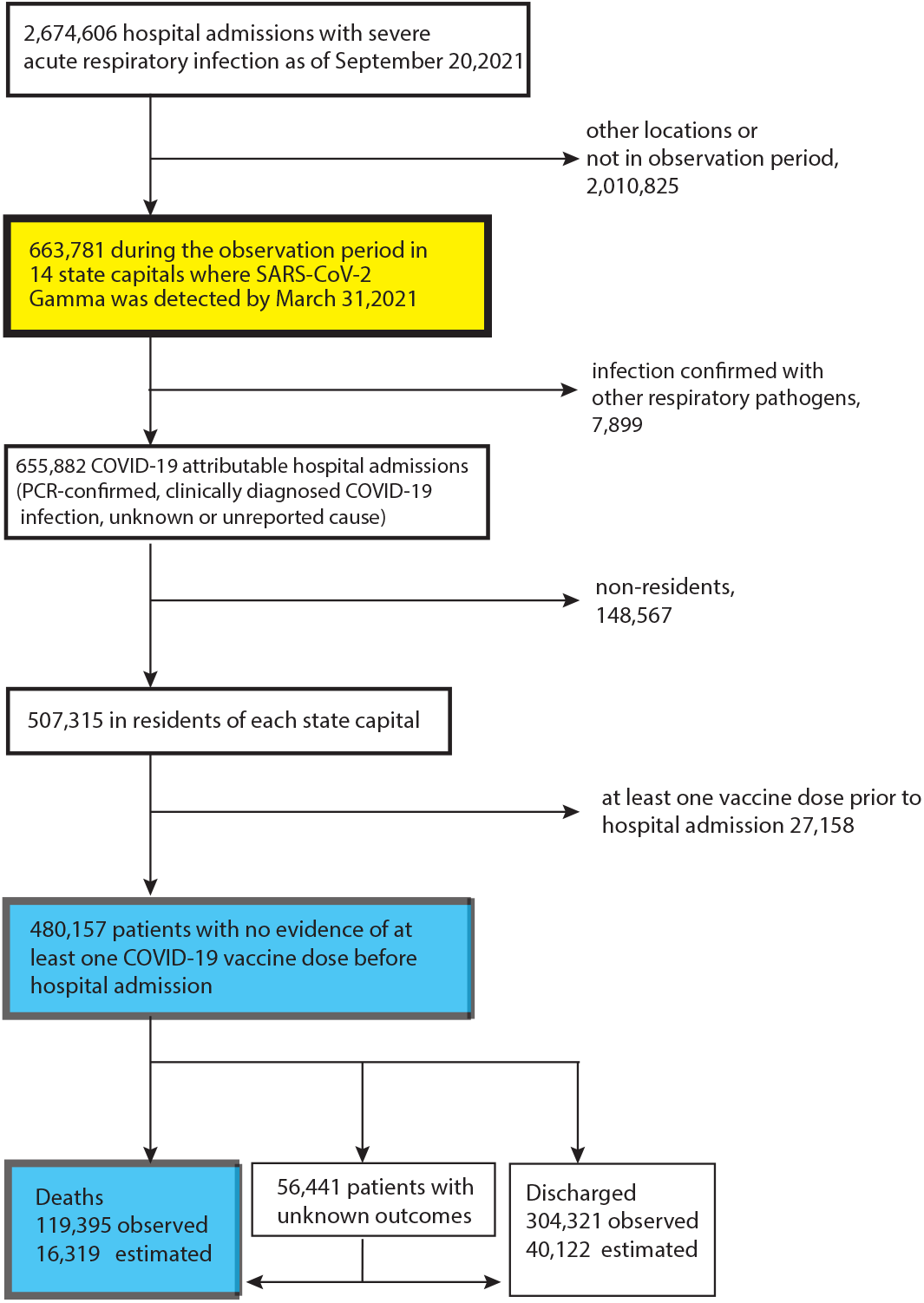
Analysis flow chart. Individual-level records on hospital admissions with severe acute respiratory infection across Brazil are mandatory to report to the SIVEP-Gripe (Sistema de Informação da Vigilância Epidemiológica da Gripe) database [12, 13], and publicly available records between 20 January 2020 and 26 July 2021 were downloaded on 20 September 2021. Data used to derive COVID-19 in-hospital fatality rates are shown in blue, and data used to derive the healthcare pressure indices are shown in yellow.

## Gamma’s expansion across Brazil was followed by transient shocks in in-hospital fatality rates

To assess the spatio-temporal expansion of Gamma in each of the 14 cities, we analyze publicly available SARS-CoV-2 genome sequences from GISAID [23] along with confirmed lineage assignments (Supplementary Text, page 6) [24]. We considered that the sequence data, which is labeled by federal unit or state, primarily captures SARS-CoV-2 spread in the corresponding state capitals (Supplementary Text, page 20). Figure 1B and Supplementary Figure S1 show Gamma’s variant frequency over time, and highlight Gamma’s swift expansion in all 14 state capitals. Concurrently, COVID-19 attributable hospital admissions and deaths rose in tandem with Gamma’s expansion in each city (Supplementary Figure S2, S3).

Over the full study period, from 20 January 2020 to 26 July 2021, 655,882 patients with PCR-confirmed, clinically diagnosed, or suspected COVID-19 infection, in short COVID-19 attributable infection [16], were admitted to hospitals across the 14 cities (Figure 2 and Supplementary Text, page 11). To match these data to city-level transmission dynamics, we here focus on the 507,315 COVID-19 attributable hospital admissions among residents in each state capital. We further excluded 27,158 resident patients who were administered at least one vaccine dose before hospitalisation to avoid confounding of time trends in fatality rates with vaccine rollout, which occurred during the same period. Thus, our study population comprised the remaining 480,157 COVID-19 attributable hospital admissions among residents. Among those, 119,395 (24.87%) outcomes were fatal. However, 56,441 (11.8%) admissions had unreported clinical outcomes, which occurred primarily after Gamma’s detection (Supplementary Figure S2), and so it is important to account for under-reporting of COVID-19 deaths [16, 25, 26]. We estimate 16,319 additional fatalities by considering the proportion of deaths among patients with known outcomes, stratified by age and observation week (Supplementary Text, page 24). Figure 1C shows for Manaus the resulting, empirical COVID-19 in-hospital fatality rates, defined as the proportion of under-reporting adjusted deaths in weekly COVID-19-attributable hospital admissions among residents with no evidence of vaccination prior to admission. We observe marked increases in COVID-19 in-hospital fatality rates after Gamma was first detected in Manaus, a pattern that is consistent across all age groups considered, 0-15, 16-29, 30-39, 40-49, 50-59, 60-69, 70-74, 75-79, 80-84, 85-89 and 90+, and across the other 13 state capitals (Supplementary Figure S4) [21]. Yet, importantly, we find COVID-19 in-hospital fatality rates also fluctuated markedly prior to Gamma’s first detection, and that increases after Gamma’s detection were transient, declining in tandem with fewer admissions in Manaus (Figure 1C).

## In-hospital fatality rates fluctuated extensively also prior to Gamma’s emergence

The observed, age-dependent patterns in COVID-19 in-hospital fatality rates are consistent with earlier observations that COVID-19 fatality rates increase with age [27]. Moreover, the within age band variation reveals stark geographical differences and temporal fluctuations since the beginning of the pandemic, and reinforces previous findings on geographical heterogeneity in the first three months of the pandemic [17]. For example, in Belo Horizonte no age group experienced shocks of COVID-19 in-hospital fatality rates above 50% that lasted at least four consecutive weeks, whereas in Porto Velho all patients of age 50 or older experienced such fatality shocks (Supplementary Table S3).

To obtain a simple measure on the extent of spatio-temporal variation, we first estimated smoothed, non-parametric trends through the weekly, age-specific in-hospital fatality rates (Supplementary Text, page 27). Then we weighted each age-specific trend in a location by the proportion of the 14 cities’ populations in the corresponding age band, resulting in an age-standardised estimate of COVID-19 in-hospital fatality rate trends. Figure 3A shows the age-standardised COVID-19 in-hospital fatality rates since the beginning of the pandemic across locations in black lines, and Table 1 reports the minimum and maximum observed values in each location. In Belo Horizonte, the minimum age-standardised COVID-19 in-hospital fatality rate before Gamma’s detection was 7.7%, and the maximum value over the entire study period was 12.2%, a 1.59-fold increase. We observed higher fold increases in all other state capitals, apart from Rio de Janeiro in which age-standardised in-hospital fatality rates were very high throughout. The minimum rates occurred either at the start of the observation period or between waves of hospital admissions in each location, tend to reach similar baseline values between shock periods in each location, and were lowest in the South and South East regions of Brazil. The maximum rates occurred in most locations after Gamma’s first detection, except for João Pessoa, Macapá, and Rio de Janeiro where they occurred preceding Gamma’s first detection. Overall, rates tended to be highest in the North, North East, and Center West regions of Brazil.

**Table 1:** Temporal fluctuations in COVID-19 attributable in-hospital fatality rate, and avoidable COVID-19 attributable deaths in hospitals.

| Location | Observation Period | COVID-19 attributable hospital admissions in unvaccinated residents |  | Age-standardised weekly COVID-19 in-hospital fatality rate (HFR) |  |  | Estimated, avoidable COVID-19 deaths in hospitals assuming the lowest HFR |  |
| --- | --- | --- | --- | --- | --- | --- | --- | --- |
|  |  | Total | Deaths* | Lowest<sup>†</sup> | Highest<sup>†</sup> | Fold increase<sup>‡</sup> | in each city<br>Percent<sup>‡</sup> | across all 14 cities<br>Percent<sup>‡</sup> |
| Belo Horizonte | 06/04/20-26/07/21 | 43,763 | 7,842 | 7.7% | 12.2% | 1.59 | 26.0%<br>(21.7%-30.5%) | 26%<br>(21.7%-30.5%) |
| Curitiba | 02/03/20-26/07/21 | 32,972 | 7,466 | 8.1% | 18.2% | 2.25 | 40.3%<br>(35.2%-45.2%) | 45.8%<br>(42.6%-49.2%) |
| Florianópolis | 09/03/20-26/07/21 | 4,072 | 916 | 7.8% | 17.1% | 2.20 | 23.6%<br>(13.5%-33.2%) | 44%<br>(40.7%-47.5%) |
| Goiânia | 16/03/20-26/07/21 | 20,881 | 6,624 | 11.5% | 27.7% | 2.42 | 47.0%<br>(41.8%-52.1%) | 61.2%<br>(58.9%-63.6%) |
| João Pessoa | 09/03/20-26/07/21 | 11,476 | 3,858 | 15.2% | 29.7% | 1.95 | 19.1%<br>(13.6%-24.8%) | 65.4%<br>(63.3%-67.5%) |
| Macapá | 30/03/20-26/07/21 | 3,209 | 1,031 | 11.2% | 41.7% | 3.73 | 41.6%<br>(30.2%-51.5%) | 68.1%<br>(66.1%-70.2%) |
| Manaus | 24/02/20-26/07/21 | 26,136 | 10,008 | 16.3% | 32.5% | 1.99 | 37.8%<br>(34.5%-41.1%) | 70.1%<br>(68.3%-72%) |
| Natal | 16/03/20-26/07/21 | 10,136 | 3,760 | 11.5% | 31.6% | 2.74 | 34.9%<br>(29.6%-40.3%) | 66.1%<br>(64.1%-68.2%) |
| Porto Alegre | 02/03/20-26/07/21 | 16,301 | 5,296 | 8.6% | 27.2% | 3.17 | 41.3%<br>(37.3%-45.3%) | 58.9%<br>(56.5%-61.4%) |
| Porto Velho | 30/03/20-26/07/21 | 7,189 | 2,618 | 11.4% | 31.9% | 2.78 | 39.1%<br>(32.6%-45.3%) | 69.8%<br>(67.9%-71.8%) |
| Rio de Janeiro | 16/03/20-26/07/21 | 77,020 | 30,194 | 20.1% | 26.5% | 1.32 | 9.0%<br>(7.4%-10.7%) | 67.2%<br>(65.3%-69.2%) |
| Salvador | 16/03/20-26/07/21 | 27,396 | 8,607 | 9.6% | 26.2% | 2.74 | 17.6%<br>(13.0%-22.1%) | 60.5%<br>(58.1%-62.9%) |
| São Luís | 24/02/20-26/07/21 | 9,370 | 2,867 | 7.6% | 26.8% | 3.51 | 35.2%<br>(28.0%-41.8%) | 60.6%<br>(58.2%-63%) |
| São Paulo | 20/01/20-26/07/21 | 190,236 | 44,620 | 8.9% | 19.8% | 2.23 | 35.6%<br>(33.4%-37.7%) | 49.4%<br>(46.4%-52.6%) |
| All 14 cities | 20/01/20-26/07/21 | 480,157 | 135,714 |  |  |  | 28.8%<br>(27.7%-29.8%) | 56.55%<br>(53.95%-59.22%) |

\* Observed deaths plus expected deaths in hospital admissions with unknown outcome. <sup>†</sup> Age-specific COVID-19 attributable in-hospital fatality rates were estimated from paired data on underreporting-adjusted deaths in COVID-19 attributable hospital admissions. Non-parametric loess estimates were obtained and weighted by the population age composition across cities. Lowest fatality rates were calculated in the period prior to Gamma's first detection in each location, and highest fatality rates were calculated including the time after Gamma's first detection. <sup>‡</sup> Estimates are based on hypothetical scenarios evaluated under the Bayesian multi-strain fatality model, assuming the lowest observed in-hospital fatality rates seen in the periods prior to Gamma's first detection in each location.

**Figure 3:**
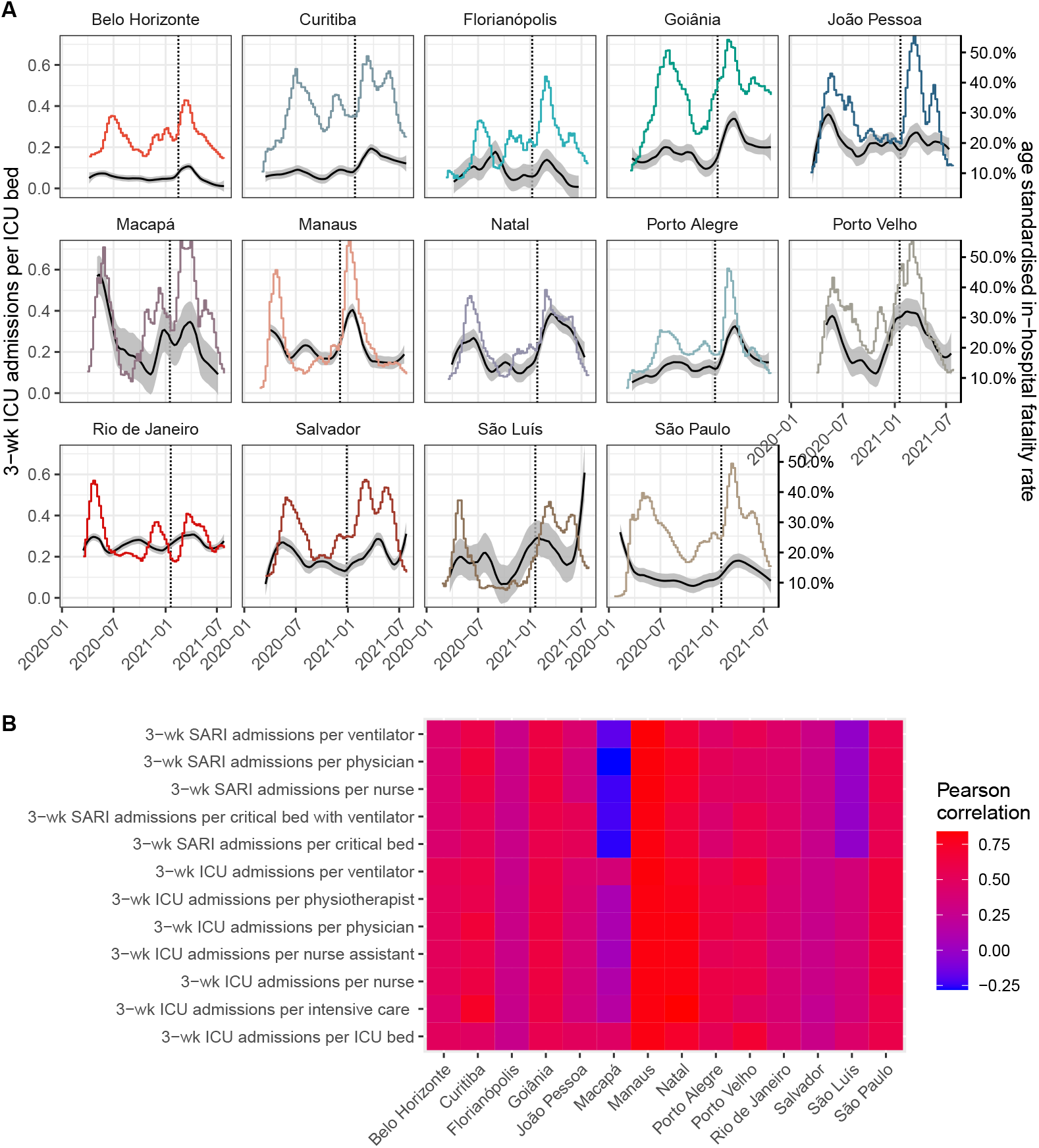
Time trends in age-standardised COVID-19 in-hospital fatality rates and pandemic healthcare pressure.(A) Detailed time evolution for the index of ICU admissions over three weeks per ICU bed. The non-parametric estimates of age-standardised COVID-19 in-hospital fatality rates (black, right hand side axis) are shown against the healthcare pressure index (colour, left hand side axis) and the date of Gamma’s first detection as vertical dashed line. (B) Heatmap of Pearson correlation coefficients between age-standardised in-hospital fatality rates and each pandemic healthcare pressure index. In Macapá, ventilators were particularly scarce at the beginning of the pandemic, resulting in overall poor correlations of all other healthcare pressure indices except those involving ventilators.

## Healthcare pressure indices track COVID-19 in-hospital death rates

Since the early phase of the COVID-19 pandemic, investments to avoid a widespread collapse of Brazil’s healthcare system have resulted in larger availability of equipment such as ventilators or intensive care unit (ICU) beds, as well as trained health care professionals, but with significant geographic differences [19, 28]. Here, in the context of substantial underfunding of Brazil’s unified health system prior to the pandemic [29, 14] and disparities in healthcare resources across and within Brazil’s states [30], we introduce pandemic healthcare pressure indices to monitor in-hospital healthcare load at the city level. We obtained healthcare-facility-level microdata on personnel (nurses, nurse assistants, physiotherapists, physicians and intensive care specialists) and equipment (critical care beds, ICU beds, ventilators) from Brazil’s National Register of Health Facilities (Cadastro Nacional de Estabelecimentos de Saúde (CNES)) [31], which we consolidated into monthly time series for each location (Supplementary Text, page 14).

We find large inequities in healthcare resources across Brazil. In March 2020 the number of available ventilators per 100,000 population ranged from 21.7 in Macapá to 102.2 in Porto Alegre, and the number of physicians per 100,000 population ranged from 124.4 in Macapá to 633.2 in Belo Horizonte (Supplementary Table S2 and Figure S5). As shown in Figure 3, we constructed time-varying healthcare pressure indices by calculating, for example, the moving sum of ICU admissions over three weeks per ICU bed (Supplementary Text, page 29). The healthcare pressure indices thus capture changes in hospital demand per available resource, with demand comprising all hospitalised patients with severe acute respiratory infection, including non-residents and individuals with vaccine breakthrough infections. We find that all healthcare pressure indices are strongly correlated with the age-standardised, weekly COVID-19 in-hospital fatality rates in most cities (Figure 3, Supplementary Figure S6).

## Decomposing effects of infection severity, location and pandemic healthcare pressure

We next developed a Bayesian multi-strain fatality model to disentangle the effects of location-specific inequities, pandemic healthcare pressures and Gamma’s disease severity on fluctuating COVID-19 in-hospital fatality rates. Briefly, the model describes Gamma’s replacement dynamics in each of the 14 state capitals as a logistic function, that is fitted to the weekly SARS-CoV-2 variant frequency data. To characterise when Gamma’s expansion started in each location, we analysed 2,212 Gamma sequences to obtain minimum estimates of Gamma’s local transmission in each location (Supplementary Text, page 20). The observed COVID-19 attributable hospital admissions in the 11 age groups are then decomposed by variant according to the estimated replacement dynamics. We model age-specific in-hospital fatality rates through a regression equation that captures non-parametric location effects, fixed effects associated with the healthcare pressure indices, and non-parametric virus variant effects associated with Gamma’s replacement dynamics. Using the modelled COVID-19 fatality rates, we first calculate the expected deaths in patients attributed to hospitalisation by variant as shown in Figure 4. Then, we sum the expected deaths across Gamma and non-Gamma variants, and fit these for each age group to the weekly, underreporting-adjusted deaths in hospitals (Supplementary Text, page 42). In the model, the non-parametric location effects do not vary in time and account for a variety of constant social, economic or healthcare related factors that differentiate one city from another, such as the prevalence of co-morbidities that might modify disease severity or pre-existing inequities in the both the quality and quantity of available healthcare resources [15, 32]. In contrast, the health-care pressure effects vary in time, and correspond independently in each location to a linear combination of the measured pandemic healthcare pressure indices.

**Figure 4:**
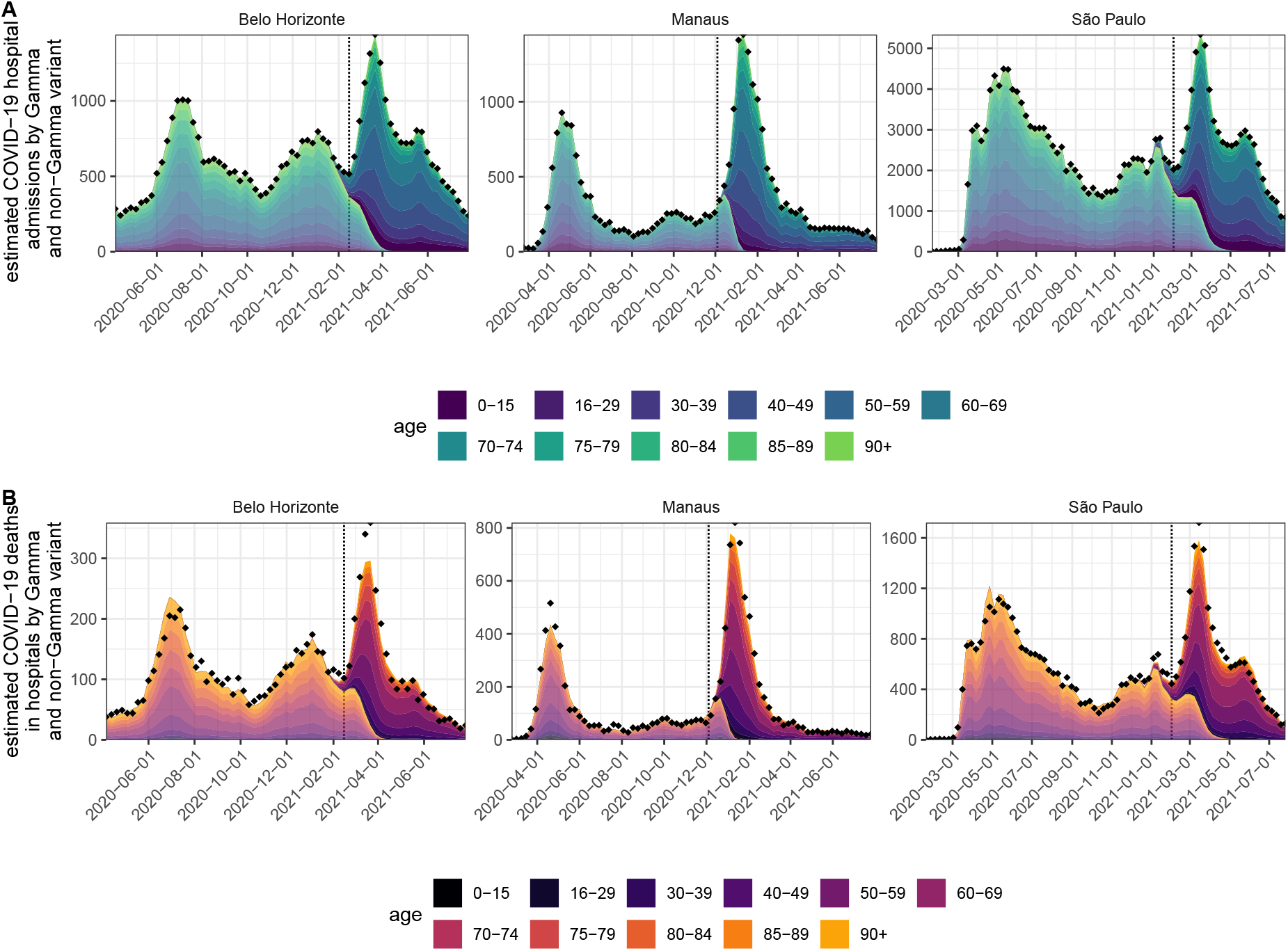
Inferred COVID-19 attributable hospital admissions and deaths among hospitalisations by SARS-CoV-2 variant. (A) Using SARS-CoV-2 variant frequency data, COVID-19 attributable hospital admissions in each age band and state capital were decomposed by Gamma and non-Gamma variant. Posterior median estimates for each age band are shown in fill colours, while estimates attributed to non-Gamma lineages are shown in lighter shades, and estimates attributed to the Gamma lineage in darker shades. The weekly totals of observed hospital admissions are shown as diamonds, and the date of Gamma’s first detection is shown as a dotted vertical line. (B) Deaths among variant-specific hospital admissions were jointly estimated under the multi-strain fatality model. Colours and shades are as in subfigure A, while weekly totals of observed deaths among hospital admissions are shown as diamonds. Admissions and deaths closely follow infection waves of different SARS-CoV-2 lineages and variants, for example the initial variant, P.2, and then Gamma in Belo Horizonte.

Brazil’s rapid vaccination roll-out overlapped with Gamma’s temporal expansion, as well as changes in Brazil’s age-specific population structure [33], and so changes in the age composition of hospital admissions could reflect any of these factors. To account for this, we used deaths records from Brazil’s Civil Registry [34] and vaccine administration records from the Brazilian Ministry of Health [35] to adjust downwards the population at risk of hospitalisation per location and age group in the model (Supplementary Text, page 32). Importantly, we observed substantial variation in the timing of vaccine roll-out, time to second dose, and vaccine type administered (Supplementary Figure S7). After accounting for this variation, the model also provided good fits to age-specific shifts in hospital admissions and deaths, as well as weekly age-specific COVID-19 hospital admissions, deaths, in-hospital fatality rates, and Gamma’s variant frequencies (Figure 4 and Supplementary Figures S1 and S8-S14).

In Figure 1C, the solid black line indicates for Manaus the estimated in-hospital fatality rates over Gamma and non-Gamma variants, and the black dotted line shows the inferred in-hospital fatality rates without Gamma’s estimated contribution to in-hospital disease severity. In Manaus, we find that the marked increase in COVID-19 in-hospital fatality rates is better explained by changes in healthcare pressures rather than a direct effect of Gamma on fatality rates in hospitalized patients. This is inferred from the fact that the empirical fatality rates (shown in dots in Figure 1C) fluctuate in the pre-Gamma period, decrease after the initial shock post-Gamma detection, and are strongly associated with the pandemic healthcare pressure indices. As of 26 July 2021, in all other state capitals, the epidemic waves post Gamma’s emergence have also subsided, and in-hospital fatality rates are declining concomitantly. However, in Curitiba, Natal, Porto Alegre and São Paulo, the empirical weekly fatality rates have to date remained above levels seen between epidemic waves for some age groups, and there, Gamma’s estimated contribution on in-hospital fatality rates is non-negligible, though not dominant (Supplementary Figures S12). We also find hospital admissions were more frequent in young adults following Gamma’s temporal expansion (Supplementary Figures S15), which is expected if Gamma’s disease severity is shifted towards younger adults [36].

## Location inequities and healthcare pressures drive Brazil’s in-hospital fatality rates

Figure 5A compares the fitted age-standardised COVID-19 in-hospital fatality rates across Brazil’s macroregions, the North, Northeast, Central-West, Southeast and South, revealing considerable geographical heterogeneity. Before Gamma’s first detection in each location, the fitted age-standardised in-hospital fatality rate was lowest in Belo Horizonte and highest in Rio de Janeiro (shown as dotted horizontal lines in Figure 5A). The high estimates for Rio de Janeiro prompted us to compare excess deaths derived from Brazilian’s Civil Registry to the COVID-19 attributable in hospital deaths, which suggested that a smaller than expected proportion of hospitalised patients with unknown clinical outcomes may have died. Yet, Rio de Janeiro’s age-standardised COVID-19 fatality rates remained the highest even when we assume that all patients with unknown clinical outcomes were successfully treated and survived COVID-19 (Supplementary Text, page 69). We then calculated the ratio of the lowest achieved rates of all other state capitals relative to that seen in Belo Horizonte, and interpret these ratios as the contribution of location to in-hospital fatality rates, which measure non-specific baseline differences in fatality rates across the state capitals that are not captured in our healthcare pressure indices. Further, we extract from the model, for each location, the multipliers to the minimum fatality rates that are specifically associated with the pandemic healthcare pressure indices, and interpret this multiplier as the contribution of healthcare pressure to in-hospital fatality rates. Finally, we interpret the ratio of fatality rates with and without Gamma’s non-parametric effect as Gamma’s overall contribution on in-hospital infection severity. We note that the measured Gamma effect merely reflects associations with fatality rates that are better explained with changes in population-level variant frequencies than with the observed healthcare pressure indices, and so is not capturing a causal relationship.

**Figure 5:**
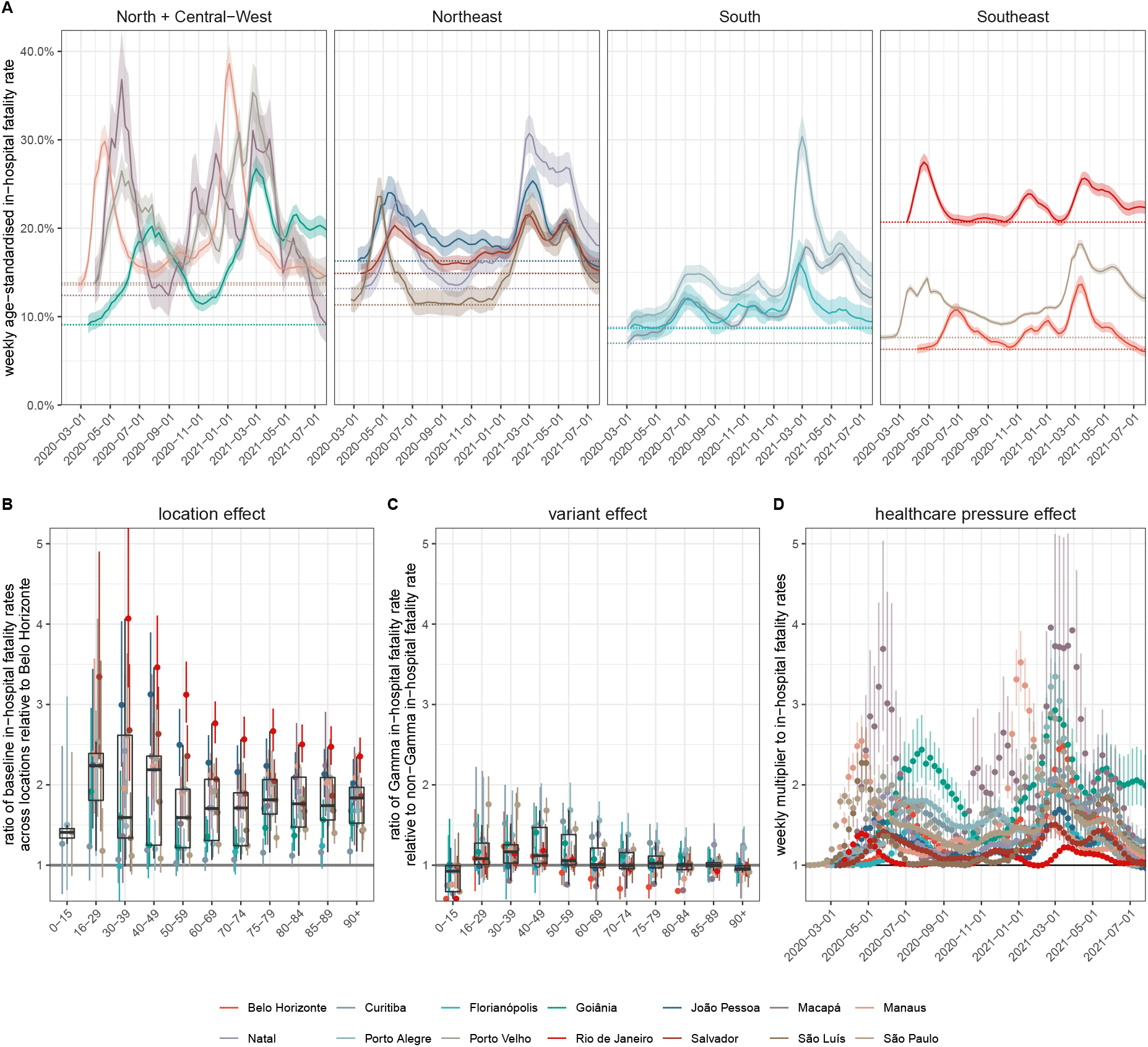
Estimated contribution of location inequity, Gamma’s infection severity, and pandemic healthcare pressure on COVID-19 in-hospital fatality rates. (A) Estimated, weekly age-standardised COVID-19 in-hospital fatality rates, averaged across SARS-CoV-2 variants. Posterior median estimates (line) are shown with 95%CrIs (ribbon), and the lowest estimated fatality rates during the observation period in each state capital (dotted horizontal line). (B) Estimated ratio in lowest in-hospital fatality rates in each location relative that seen in Belo Horizonte. (C) Estimated ratio in in-hospital fatality rates for Gamma versus non-Gamma lineages. (D) Estimated multiplier to the lowest age-standardised fatality rates shown in subplot A that is associated with the pandemic healthcare pressure indices. In each plot, posterior median estimates are shown as dots, 95%CrIs as linerange, and summaries across locations as boxplots.

Figures 5B-D compare the estimated location, Gamma and healthcare pressure contributions. Relative to Belo Horizonte, the lowest fatality rates were an estimated 1.94 (1.08-3.34) fold higher in all other state capitals, a location effect that was consistent across all age groups. Whilst we do not explore the exact drivers of this “location” effect, it is likely shaped by a combination of socio-economic factors specific to each city, in conjunction with city-specific, pre-existing inequities in the availability, quality and accessibility of healthcare [37, 38]. At peak times, pandemic healthcare pressures as measured by our indices were associated with a 2.29 (1.37-4.24) fold multiplicative effect across the 14 cities, and this contribution was stronger in places that already had high in-hospital fatality rates at baseline. Gamma was not associated with a statistically significant effect on in-hospital fatality rates after controlling for healthcare pressures (1.14 (0.51-1.79) fold increase among patients aged 40-49, and declining in younger and older ages). Importantly, this suggests that the COVID-19 fatality rate shocks seen in hospitals can be explained with substantively increased demand on limited healthcare resources that follow infection waves, and that these shocks are strongest in places where healthcare resources are lowest. Indeed, we observed fatality rate shocks were recurring prior to the emergence of Gamma and prior to widespread acquired immunity due to vaccination. Following Gamma’s emergence, are likely the result of Gamma’s increased transmissibility and ability to evade prexisting natural immunity from previously circulating SARS-CoV-2 lineages - thereby increasing the availability of susceptible individuals, and leading to a new epidemic wave. We are only able to measure Gamma’s effect following hospitalisation, and so it is possible that healthcare demand is further amplified through increased risk of hospitalisation following infection and thus overall higher infection severity, as has been shown for SARS-CoV-2 Alpha [39, 40].

## Projected avoidable deaths in the absence of resource limitations

To quantify the impact that the observed fluctuations in COVID-19 attributable in-hospital rates had on the death toll in the 14 state capitals, we considered counterfactual simulations where infection waves and resource limitations did not result in surging in-hospital fatality rates (Supplementary Text, page 56). We consider this counterfactual conservative as it assumes for each location achievable in-hospital fatality rates that are based on the lowest values observed prior to Gamma’s first detection in each location, and does not account for improvements in clinical management, patient triage, treatments (e.g. Dexamethasone). Across the 14 state capitals, we find deaths could have been, in the absence of pandemic resource limitations, reduced by an estimated 28.8% (27.7%-29.8%) (Table 1), which are highest in patients aged 50-74 due to Brazil’s age pyramid (Supplementary Figures S16-S17). More than one in three deaths could have been averted in Curitiba, Goiânia, Macapá, Manaus, Natal, Porto Alegre, Porto Velho, São Luís, São Paulo. Taking the percent reduction across the 14 state capitals as indicative and generalisable to all of Brazil, we estimate that as of 26 July 2021, 167,194 (160,809-173,000) of Brazil’s observed 580,538 COVID-19 attributable deaths in hospitals could have been avoided in the absence of pandemic resource limitations.

We further evaluated in counterfactual simulations how many in-hospital deaths could have been avoided in the absence of location inequities and pandemic resource limitations, i. e. if all 14 state capitals would have experienced the lowest observed in-hospital fatality rates per age group throughout the pandemic, which were observed in Belo Horizonte. We consider these projections less conservative, since the observed, low in-hospital fatality rates in Belo Horizonte may reflect a large range of factors that may not be translatable to other locations in Brazil. We find in-hospital deaths could have been reduced by an estimated 56.55% (53.95%-59.22%) in the 14 state capitals (Table 1 and Supplementary Figures S16-S17). Furthermore, extrapolating Belo Horizonte’s lowest observed in-hospital fatality rates to all of Brazil, we estimate that as of 26 July 2021, 328,294 (313,200-343,794) of Brazil’s COVID-19 attributable in-hospital deaths could have been avoided.

## Limitations

The findings of this study should be considered in the context of the following limitations. First, Brazil’s line list in-hospital patient data is limited in that co-morbidity factors, vaccination status or SARS-CoV-2 genotype data are either frequently missing or not available at the individual level [41], and so it is challenging to comprehensively control for individual-level factors that can modulate fatality rates [39, 42]. Yet, Brazil’s freely accessible line list data constitutes one of the world’s largest databases to characterise the pandemic impact of COVID-19 in a middle-income country, and across areas that differ substantially in baseline development and available health-care resources. In sensitivity analyses we considered SARS-CoV-2 sequence data from alternative sources as well as alternative patient inclusion criteria, which suggest that our estimates of location and healthcare pressure effect sizes are robust (Supplementary Text, page 58). Second, we here delineate fluctuations in COVID-19 in-hospital fatality rates at city level to focus on the extreme spatio-temporal heterogeneity in fatality rates across Brazil. We recognise this broader geographic focus masks important differences within cities, and thus we cannot identify the exact factors determining the substantial location effect that we measure. However, these are likely related to differences in catchment populations as vulnerable populations with poor healthcare access are highly clustered across Brazil’s largest cities [30, 43], underfunding of the public healthcare system and emerging discrepancies in healthcare resources compared to private hospitals [17, 37, 43, 44], or inequities in the quality and capabilities of healthcare systems which, for example, have been documented pre-pandemic in sepsis survival rates (with differences particularly marked between private and public hospitals [45]). Additional relevant factors could include differences in demand reflecting variation in epidemic magnitude [37], or the timing and extent of measures aimed at controlling spread and preventing infection in vulnerable groups [19, 38, 28], suggesting that the location effects could also reflect healthcare pressures that are present already at the lowest incidences of COVID-19 hospital admissions in each location. Importantly, we observe substantial fluctuations in in-hospital fatality rates even in large private hospitals of São Paulo city (Supplementary Text, page 66), suggesting that large effects of healthcare pressure on in-hospital fatality rates are common, and present even in high-income countries [46]. Third, the pandemic healthcare pressure indices that we derive are based on data reported to CNES, which does not capture all resource limitations such as the acute shortages in oxygen supply that were experienced in January 2021 in Manaus [47], and is prone to potential reporting biases [20]. In our view, the inferred associations between healthcare pressure indices and fatality rates demonstrate that where resources are limited, real-time monitoring of available resources is especially important to identify critical resource limitations and avoid the catastrophic shocks in death rates that we describe for many state capitals across Brazil. Finally, our analyses start with hospitalisation, which is a limitation because in-hospital fatality rates also depend on who, and under what circumstances, severely ill patients are admitted to hospitals. It is possible that Gamma is associated with increased probability of requiring hospitalisation as other SARS-CoV-2 variants [39, 40]. We also find evidence that in several locations, out-of-hospital deaths have surged at times of peak demand (Supplementary Figure S3), and that in hospitalised patients with a fatal outcome, the time to death following admission tended to be shorter during peak demand (Supplementary Text, page 70). These observations indicate that increased healthcare pressure acts to shape in-hospital fatality rates through distinct mechanisms, likely through a combination of both the reduced ability to provide adequate care and an increase in the average severity of admitted patients (with only the most severely ill admitted during periods of highest pressure). In this context, we expect that the projected proportion of avoidable COVID-19 in-hospital deaths in the absence of healthcare pressure effects would be lower if less severe COVID-19 cases could also have been admitted. At the same time, the projected numbers are likely an underestimate of COVID-19 deaths that could have been avoided in the absence of healthcare pressure effects because we did not account for deaths in severely ill individuals that were not cared for in hospitals.

## Summary

This study highlights important geographic and temporal variation in COVID-19 in-hospital fatality rates since the beginning of the epidemic in 14 state capitals across Brazil. This variation is driven primarily by shortages in health-care capacity, which in turn emerges from a combination of pre-pandemic inequities at city-level and increased healthcare pressure brought about by epidemic waves of SARS-CoV-2 transmission. The extent and degree of this mismatch between supply (of staff and equipment) and demand (patients requiring hospitalisation for COVID-19) is highly dynamic, varying significantly over the course of an epidemic. The implications of these location inequities and healthcare shortages are dramatic. As of 26 July 2021, we project that approximately one quarter of Brazil’s COVID-19 attributable deaths in hospitals could have been avoided if healthcare pressure had not exacerbated baseline fatality rates, and we project approximately half of Brazil’s COVID-19 attributable deaths in hospitals could have been avoided if in addition all hospitals would have had the same baseline COVID-19 fatality rates as those observed in Belo Horizonte. Our findings are particularly important in calibrating the risk posed by new SARS-CoV-2 variants of concern. We find that the impact of Gamma in Brazil’s hospitals has predominantly been indirect and mediated through pre-existing geographic inequities, transient infection waves and concomitant shocks in healthcare demand. In conclusion, our results suggest that investments in healthcare resources, health-care optimization, and pandemic preparedness are critical to minimize population wide mortality and morbidity caused by highly transmissible and deadly pathogens such as SARS-CoV-2, especially in low- and middle-income countries.

## Data Availability

All data produced are available online at:
https://github.com/CADDE-CENTRE/covid19_brazil_hfr

https://opendatasus.saude.gov.br/dataset/bd-srag-2021

https://transparencia.registrocivil.org.br/

https://github.com/capyvara/brazil-civil-registry-data

https://opendatasus.saude.gov.br/dataset/covid-19-vacinacao/resource/ef3bd0b8-b605-474b-9ae5-c97390c197a8

https://www.ibge.gov.br/estatisticas/sociais/populacao/9171-pesquisa-nacional-por-amostra-de-domicilios-continua-mensal.html?=&t=o-que-e

https://github.com/CADDE-CENTRE/covid19_brazil_hfr

## Acknowledgments

We thank all contributors to GISAID for making SARS-CoV-2 sequence data information publicly available as listed in the Supplementary Text; all contributors to Rede Genomica Fiocruz for making SARS-CoV-2 variant frequency data publicly available; all members of the CADDE network for their comments throughout the project and earlier versions of the manuscript; Oliver G. Pybus, Andrew Rambaut and JT. McCrone for their insightful comments on SARS-CoV-2 phylogenetic analyses; and the Imperial College Research Computing Service, DOI: 10.14469/hpc/2232, for providing the computational resources to perform this study.

## Funding

This study was supported by the Medical Research Council-São Paulo Research Foundation (FAPESP) CADDE partnership award (MR/S0195/1 and FAPESP18/14389-0) (https://caddecentre.org), by the EPSRC through the EPSRC Centre for Doctoral Training in Modern Statistics and Statistical Machine Learning at Imperial and Oxford. RSA from the Rede Coronaômica BR MCTI/FINEP affiliated to RedeVírus/MCTI (FINEP 01.20.0029.000462/20, CNPq 404096/2020-4), from CNPq (312688/2017-2 and 439119/2018-9), MEC/CAPES (14/2020 - 23072.211119/2020-10), FINEP (0494/20 01.20.0026.00); LSB acknowledges support from Inova Fiocruz (48401485034116); DSC is funded by the Clarendon Fund, University of Oxford Department of Zoology and Merton College; NF from Well-come Trust and Royal Society (N.R.F.: Sir Henry Dale Fellowship. 204311/Z/16/Z) and from the Bill & Melinda Gates Foundation (INV-034540); CAP was supported by FAPESP (2019/21858-0), Fundação Faculdade de Medicina and Coordenação de Aperfeiçoamento de Pessoal de Nível Superior – Brasil (CAPES) – Finance Code 001 OTR from the Instituto de Salud Carlos III (Sara Borrell fellowship, CD19/00110); OR from the Bill & Melinda Gates Foundation (OPP1175094); ES from Bill & Melinda Gates Foundation (INV-034652); RPS from the Rede Coronaômica BR MCTI/FINEP affiliated to RedeVírus/MCTI (FINEP 01.20.0029.000462/20, CNPq 404096/2020-4), from CNPq (310627/2018-4), MEC/CAPES (14/2020 - 23072.211119/2020-10), FINEP (0494/20 01.20.0026.00), FAPEMIG (APQ-00475-20); WMS from FAPESP (2017/13981-0, 2019/24251-9) and the NIH (AI12094); CW acknowledges an MRC Doctoral Training Partnership Studentship.

## Author contributions

OR conceived the study. ECS, NRF, SB, OR oversaw the study. ABr, LMSS, IH, CAP, WMS, RSA, HJA, LJTA, JLFA, RMS, PLCF, VEVG, DMB, DCQ, RS oversaw and performed data collection. ABr, CW, LMSS, IH, CAP, WMS lead the analysis. ABl, LFB, DC, MCC, SSC, JC, ASS, CD, SF, BG, PL, ASL, JTMcC, TM, XM, SM, MM, RGM, FRRM, BN, RHMP, OR, RPS, ES, RS, RPS, XX contributed to the analysis. All authors discussed the results and contributed to the revision of the final manuscript.

## Competing interests

No competing interests to declare.

## List of Supplementary Texts, Tables and Figures

**Table S1**. Reported and under reporting-adjusted COVID-19 attributable in-hospital deaths.

**Table S2**: Healthcare resources, March 2020 to July 2021.

**Table S3**: Longest observed duration of COVID-19 in-hospital fatality rates above 50%.

**Figure S1**: Spatiotemporal expansion of the SARS-CoV-2 Gamma variant across Brazil. SARS-CoV-2 genome sequences were obtained from the GISAID repository along with confirmed lineage assignments.

**Figure S2**: COVID-19 attributable hospital admissions among residents without evidence of vaccination.

**Figure S3**: Under reporting-adjusted COVID-19 attributable deaths.

**Figure S4**: Time trends in age-specific COVID-19 in-hospital fatality rates.

**Figure S5**: Healthcare resources in hospital settings, per 100,000.

**Figure S6**: Time evolution of pandemic healthcare pressure indices.

**Figure S7**: COVID-19 vaccine coverage.

**Figure S8**: Model fit of expected weekly COVID-19 attributable hospital admissions.

**Figure S9**: Model fit of expected weekly COVID-19 attributable deaths.

**Figure S10**: Estimated COVID-19 attributable hospital admissions by SARS-Cov-2 variant.

**Figure S11**: Estimated COVID-19 attributable deaths in hospitals by SARS-Cov-2 variant.

**Figure S12**: Model fits to age-specific COVID-19 in-hospital fatality rates.

**Figure S13**: Model fits of the expected age composition of COVID-19 attributable hospital admissions.

**Figure S14**: Model fits of the expected age composition of COVID-19 attributable deaths following hospital admission.

**Figure S15**: Estimated ratio in the share of age groups in Gamma versus non-Gamma residents’ hospital admissions.

**Figure S16**: Projected proportion of avoidable COVID-19 attributable deaths in hospitals with-out pandemic resource limitations.

**Figure S17**: Projected proportion of avoidable COVID-19 attributable deaths in hospitals with-out location inequities and without pandemic resource limitations.

**Supplementary Text:** Supplementary Text to Factors driving extensive spatial and temporal fluctuations in COVID-19 fatality rates in Brazilian hospitals.

**Supplementary References:** (48-119)

## Notes

### Competing Interest Statement

The authors have declared no competing interest.

### Funding Statement

This study was supported by the Medical Research Council-Sao Paulo Research Foundation (FAPESP) CADDE partnership award (MR/S0195/1 and FAPESP18/14389-0)
(https://caddecentre.org),by the EPSRC through the EPSRC Centre for Doctoral Training in Modern Statistics and Statistical Machine Learning at Imperial and Oxford. RSA from the Rede Coronaomica BR MCTI/FINEP affiliated to RedeVirus/MCTI
(FINEP 01.20.0029.000462/20, CNPq 404096/2020-4), from CNPq (312688/2017-2 and 439119/2018-9),
MEC/CAPES (14/2020 - 23072.211119/2020-10), FINEP (0494/2001.20.0026.00); LSB acknowledges support from Inova Fiocruz (48401485034116); DSC is funded by the Clarendon Fund, University of Oxford Department of Zoology and Merton College; NF from Wellcome Trust and Royal Society (N.R.F.: Sir Henry Dale Fellowship. 204311/Z/16/Z) and from the Bill & Melinda Gates Foundation (INV-034540); CAP was supported by FAPESP (2019/21858-0), Fundacao Faculdade de Medicina and Coordenacao de Aperfeicoamento de Pessoal de Nivel Superior
Brasil (CAPES) Finance Code 001 OTR from the Instituto de Salud Carlos III (Sara Borrell fellowship, CD19/00110); OR from the Bill & Melinda Gates Foundation (OPP1175094); ES from Bill & Melinda Gates Foundation (INV-034652); RPS from the Rede Coronaomica BR MCTI/FINEP affiliated to RedeVirus/MCTI (FINEP 01.20.0029.000462/20, CNPq 404096/2020-4),from CNPq (310627/2018-4), MEC/CAPES (14/2020 - 23072.211119/2020-10), FINEP
(0494/20 01.20.0026.00), FAPEMIG (APQ-00475-20); WMS from FAPESP (2017/13981-0, 2019/24251-9) and the NIH (AI12094); CW acknowledges an MRC Doctoral Training Partnership Studentship

### Author Declarations

This study involves only openly available human data, which can be obtained from: - the SIVEP-Gripe platform (https://opendatasus.saude.gov.br/dataset/bd-srag-2021) - the Brazilian Civil Registry (https://transparencia.registrocivil.org.br/) - the Brazilian Ministry of Health (https://opendatasus.saude.gov.br/dataset/covid-19-vacinacao/resource/ef3bd0b8-b605-474b-9ae5-c97390c197a8)

