## Supplementary Figures and Tables for "Report 46: Factors driving extensive spatial and temporal fluctuations in COVID-19 fatality rates in Brazilian hospitals"

Affiliations

DOI: https://doi.org/10.25561/91875 

*Contributed equally, **joint senior corresponding authors.

This PDF file includes:
Supplementary Tables S1-S3
Supplementary Figures S1-S17

| Location | Reported COVID-19 attributable in-hospital deaths† | | | | Underreporting-adjusted COVID-19 attributable in-hospital deaths‡ | | | |
|---|---|---|---|---|---|---|---|---|
| | Total | Ages | | | Total | Ages | | |
| | | 0-49 | 50-74 | 75+ | | 0-49 | 50-74 | 75+ |
| Belo Horizonte | 7388 | 772 | 3576 | 3040 | 8142 | 891 | 3942 | 3309 |
| Curitiba | 7412 | 1103 | 3856 | 2453 | 7719 | 1151 | 4009 | 2559 |
| Florianópolis | 903 | 86 | 443 | 374 | 938 | 92 | 458 | 388 |
| Goiânia | 6112 | 996 | 3117 | 1999 | 6811 | 1158 | 3469 | 2184 |
| João Pessoa | 2792 | 503 | 1316 | 973 | 4068 | 769 | 1911 | 1388 |
| Macapá | 996 | 241 | 510 | 245 | 1069 | 253 | 552 | 264 |
| Manaus | 9812 | 1847 | 5295 | 2670 | 10545 | 1982 | 5691 | 2872 |
| Natal | 3073 | 457 | 1455 | 1161 | 3938 | 618 | 1890 | 1430 |
| Porto Alegre | 5143 | 514 | 2631 | 1998 | 5446 | 562 | 2785 | 2099 |
| Porto Velho | 2153 | 471 | 1198 | 484 | 2667 | 611 | 1460 | 596 |
| Rio de Janeiro | 23464 | 3092 | 11597 | 8775 | 33917 | 4764 | 16943 | 12210 |
| Salvador | 7584 | 1157 | 3700 | 2727 | 9060 | 1483 | 4456 | 3121 |
| São Luís | 2109 | 366 | 1011 | 732 | 3071 | 526 | 1503 | 1042 |
| São Paulo | 40454 | 5390 | 20158 | 14906 | 46876 | 6440 | 23337 | 17099 |

† COVID-19 attributable in-hospital deaths were defined as deaths in hospitalised unvaccinated residents with PCR confirmed or clinically diagnosed COVID-19 infection, or severe respiratory infection with no other confirmed cause. Data capture admissions and deaths until 26 July 2021.

‡ To reported counts were added expected fatal outcomes in COVID-19 attributable hospital admissions in unvaccinated residents with unreported outcomes (Supplementary Text, page 24).

**Table S1:** Reported and underreporting-adjusted COVID-19 attributable in-hospital deaths.

DOI: https://doi.org/10.25561/91875

| | Date | Healthcare resources per 100,000 population[†] | | | | | | |
|---|---|---|---|---|---|---|---|---|
| | | Belo Horizonte | Curitiba | Florianó-polis | Goiânia | João Pessoa | Macapá | Manaus |
| Intensive Care Specialists | March 2020 | 2 | 3 | 3 | 2 | 7 | 0 | 2 |
| | July 2021 | 2(0%) | 3(0%) | 4(33%) | 2(0%) | 6(-14%) | 0 | 2(0%) |
| Nurses | March 2020 | 249 | 185 | 251 | 151 | 212 | 129 | 114 |
| | July 2021 | 306(23%) | 228(23%) | 299(19%) | 180(19%) | 238(12%) | 164(27%) | 152(33%) |
| Physicians | March 2020 | 631 | 482 | 615 | 368 | 309 | 124 | 155 |
| | July 2021 | 674(7%) | 520(8%) | 686(12%) | 394(7%) | 345(12%) | 139(12%) | 166(7%) |
| Physiotherapists | March 2020 | 74 | 65 | 86 | 50 | 76 | 47 | 19 |
| | July 2021 | 92(24%) | 84(29%) | 117(36%) | 59(18%) | 90(18%) | 56(19%) | 29(53%) |
| Nurse assistants | March 2020 | 584 | 356 | 479 | 371 | 309 | 318 | 290 |
| | July 2021 | 723(24%) | 491(38%) | 601(25%) | 427(15%) | 367(19%) | 350(10%) | 328(13%) |
| CC Beds | March 2020 | 41 | 41 | 35 | 51 | 27 | 10 | 11 |
| | July 2021 | 56(37%) | 63(54%) | 54(54%) | 86(69%) | 59(119%) | 28(180%) | 28(155%) |
| CC Beds with ventilator | March 2020 | 40 | 41 | 30 | 36 | 23 | 9 | 11 |
| | July 2021 | 55(38%) | 62(51%) | 51(70%) | 70(94%) | 54(135%) | 25(178%) | 26(136%) |
| ICU Beds | March 2020 | 34 | 33 | 24 | 32 | 22 | 6 | 9 |
| | July 2021 | 48(41%) | 56(70%) | 46(92%) | 66(106%) | 51(132%) | 22(267%) | 24(167%) |
| Ventilators | March 2020 | 83 | 79 | 86 | 77 | 57 | 21 | 38 |
| | July 2021 | 101(22%) | 112(42%) | 108(26%) | 113(47%) | 87(53%) | 41(95%) | 58(53%) |
| | | Natal | Porto Alegre | Porto Velho | Rio de Janeiro | Salvador | São Luís | São Paulo |
| Intensive Care Specialists | March 2020 | 4 | 6 | 2 | 5 | 6 | 1 | 3 |
| | July 2021 | 4(0%) | 6(0%) | 2(0%) | 6(20%) | 7(17%) | 2(100%) | 4(33%) |
| Nurses | March 2020 | 162 | 290 | 168 | 185 | 229 | 223 | 183 |
| | July 2021 | 201(24%) | 334(15%) | 250(49%) | 228(23%) | 265(16%) | 258(16%) | 219(20%) |
| Physicians | March 2020 | 313 | 626 | 237 | 299 | 329 | 207 | 324 |
| | July 2021 | 348(11%) | 654(4%) | 293(24%) | 326(9%) | 344(5%) | 229(11%) | 358(10%) |
| Physiotherapists | March 2020 | 43 | 67 | 44 | 35 | 73 | 53 | 41 |
| | July 2021 | 55(28%) | 79(18%) | 76(73%) | 41(17%) | 80(10%) | 71(34%) | 55(34%) |
| Nurse assistants | March 2020 | 378 | 624 | 461 | 291 | 374 | 497 | 214 |
| | July 2021 | 451(19%) | 868(39%) | 608(32%) | 422(45%) | 432(16%) | 569(14%) | 271(27%) |
| CC Beds | March 2020 | 41 | 55 | 37 | 44 | 55 | 37 | 37 |
| | July 2021 | 68(66%) | 88(60%) | 86(132%) | 56(27%) | 67(22%) | 58(57%) | 52(41%) |
| CC Beds with ventilator | March 2020 | 36 | 53 | 35 | 38 | 46 | 33 | 30 |
| | July 2021 | 63(75%) | 86(62%) | 82(134%) | 50(32%) | 63(37%) | 48(45%) | 48(60%) |
| ICU Beds | March 2020 | 27 | 45 | 31 | 22 | 42 | 24 | 18 |
| | July 2021 | 53(96%) | 82(82%) | 78(152%) | 31(41%) | 60(43%) | 37(54%) | 37(106%) |
| Ventilators | March 2020 | 75 | 102 | 68 | 72 | 83 | 65 | 66 |
| | July 2021 | 123(64%) | 156(53%) | 147(116%) | 94(31%) | 109(31%) | 87(34%) | 86(30%) |

[†] Numbers summarise healthcare facility level microdata on personnel (nurses, nurse assistants, physiotherapists, physicians and critical care specialists, i. e. intensive care physicians), and reported equipment (critical care beds, ICU beds, ventilators) from Brazil's National Register of Health Facilities (Cadastro Nacional de Estabelecimentos de Saúde - CNES); see Supplementary Text, page 14. Values in brackets give the percent increase from March 2020 to July 2021. CC stands for critical care.

**Table S2:** Healthcare resources, March 2020 to July 2021.

| Location | Longest observed duration of COVID-19 in-hospital fatality rates above 50% (in weeks, by age group shown in years) | | | | | | | | | | |
|---|---|---|---|---|---|---|---|---|---|---|---|
| | 0-15 | 16-29 | 30-39 | 40-49 | 50-59 | 60-69 | 70-74 | 75-79 | 80-84 | 85-89 | 90+ |
| Belo Horizonte | 0 | 0 | 0 | 0 | 0 | 0 | 0 | 0 | 0 | 0 | 0 |
| Curitiba | 0 | 0 | 0 | 0 | 0 | 0 | 3 | 2 | 2 | 4 | 9 |
| Florianópolis | 0 | 0 | 0 | 0 | 0 | 0 | 1 | 7 | 7 | 7 | 21 |
| Goiânia | 0 | 0 | 0 | 0 | 0 | 0 | 7 | 12 | 11 | 22 | 23 |
| João Pessoa | 0 | 0 | 0 | 0 | 0 | 3 | 8 | 19 | 39 | 25 | 19 |
| Macapá | 0 | 7 | 0 | 8 | 9 | 11 | 15 | 20 | 23 | 53 | 31 |
| Manaus | 0 | 0 | 0 | 1 | 4 | 9 | 9 | 13 | 51 | 26 | 50 |
| Natal | 0 | 0 | 0 | 0 | 0 | 23 | 15 | 25 | 30 | 48 | 45 |
| Porto Alegre | 0 | 0 | 0 | 0 | 0 | 6 | 9 | 10 | 13 | 56 | 47 |
| Porto Velho | 0 | 0 | 0 | 0 | 0 | 8 | 16 | 30 | 25 | 48 | 64 |
| Rio de Janeiro | 0 | 0 | 0 | 0 | 0 | 0 | 23 | 72 | 72 | 72 | 72 |
| Salvador | 0 | 0 | 0 | 0 | 3 | 0 | 10 | 11 | 15 | 38 | 29 |
| São Luís | 1 | 2 | 3 | 0 | 0 | 2 | 2 | 13 | 11 | 16 | 14 |
| São Paulo | 0 | 1 | 0 | 0 | 0 | 4 | 7 | 2 | 8 | 20 | 19 |

**Table S3:** Longest observed duration of COVID-19 in-hospital fatality rates above 50%.

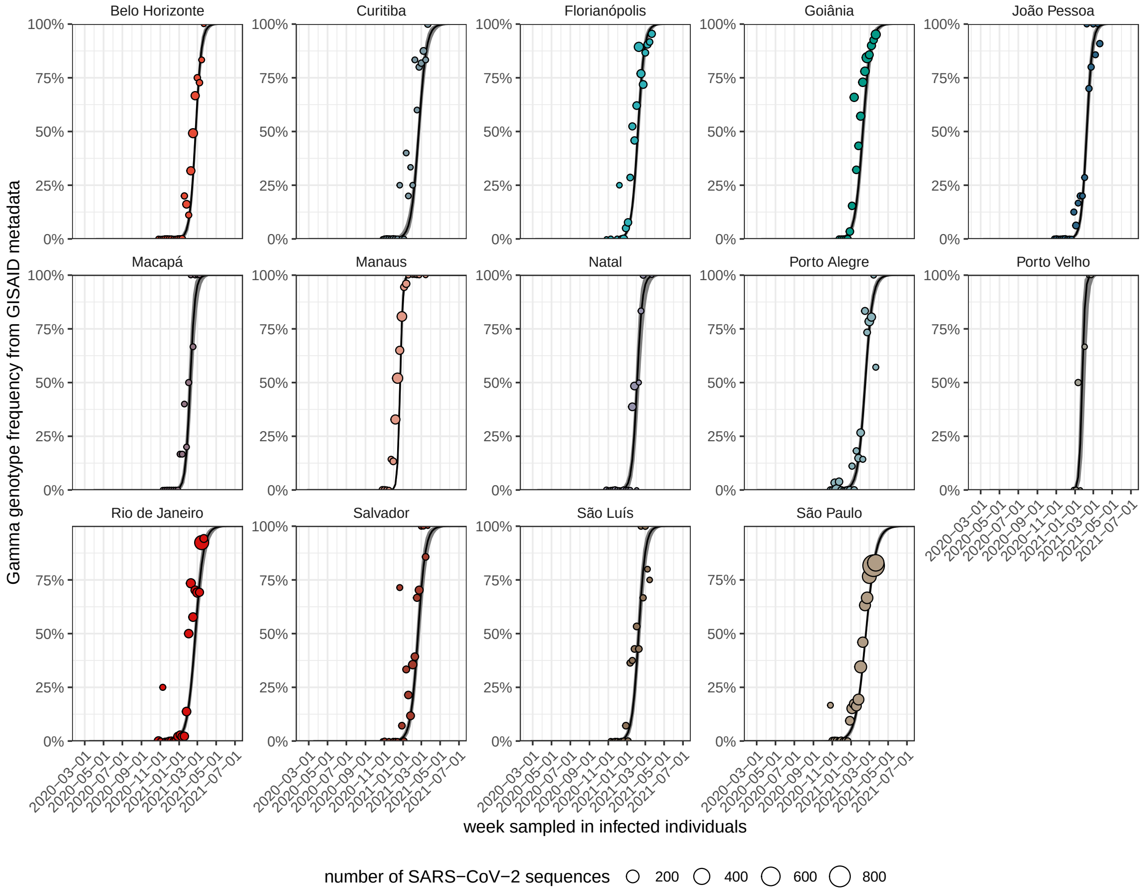


**Figure S1:** Spatiotemporal expansion of the SARS-CoV-2 Gamma variant across Brazil. SARS-CoV-2 genome sequences were obtained from the GISAID repository [1] along with confirmed lineage assignments. The frequency of the Gamma variant (dots) in weekly SARS-CoV-2 genome sequence counts (size of dots) is shown along with posterior median estimates of Gamma's variant frequencies (black line) under the Bayesian multi-strain fatality model (Supplementary Text page 44) and 95% credible intervals (grey ribbon).

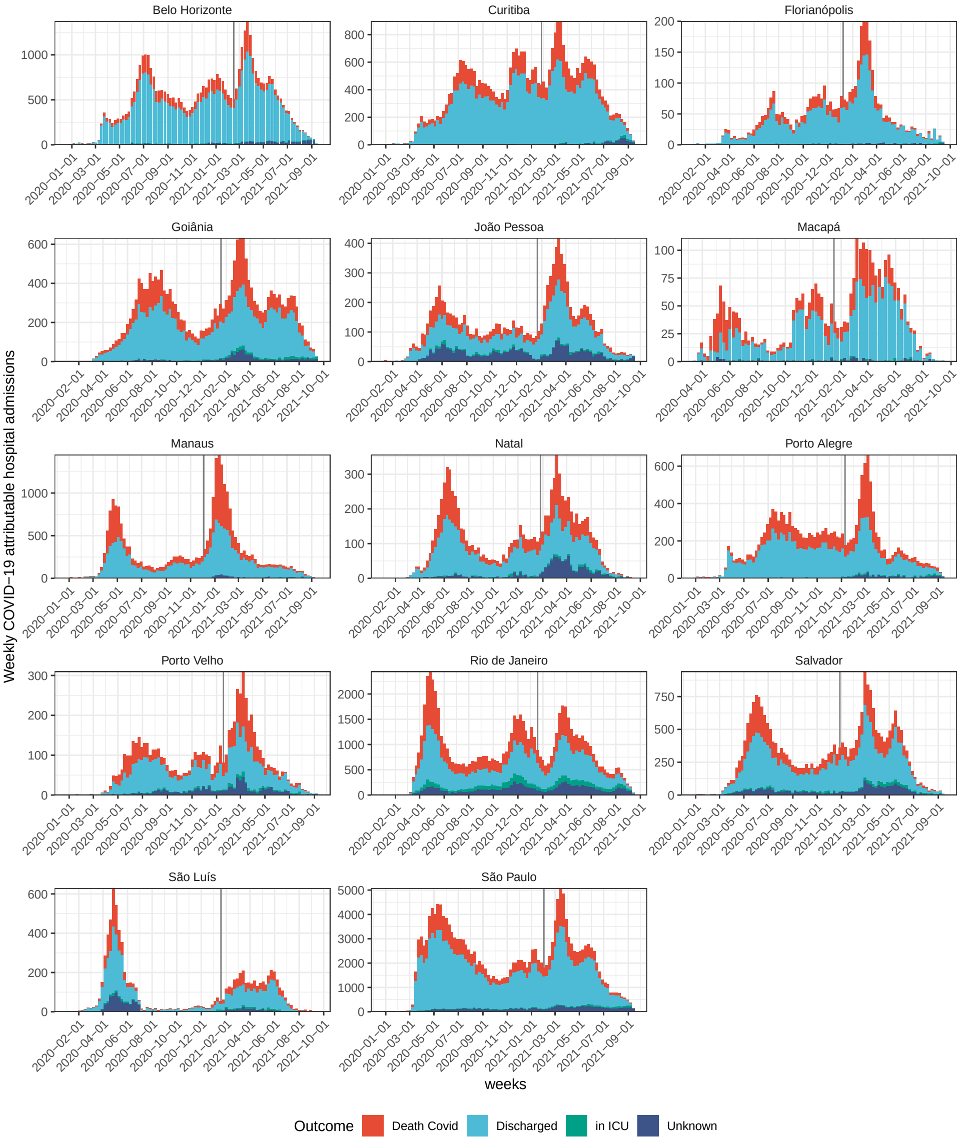


**Figure S2:** COVID-19 attributable hospital admissions among residents without evidence of vaccination. Data are from the SIVEP-Gripe platform as of 20 September 2021, and are shown in colours according to reported clinical outcomes. The date of Gamma's first detection in each city is shown as a vertical line.

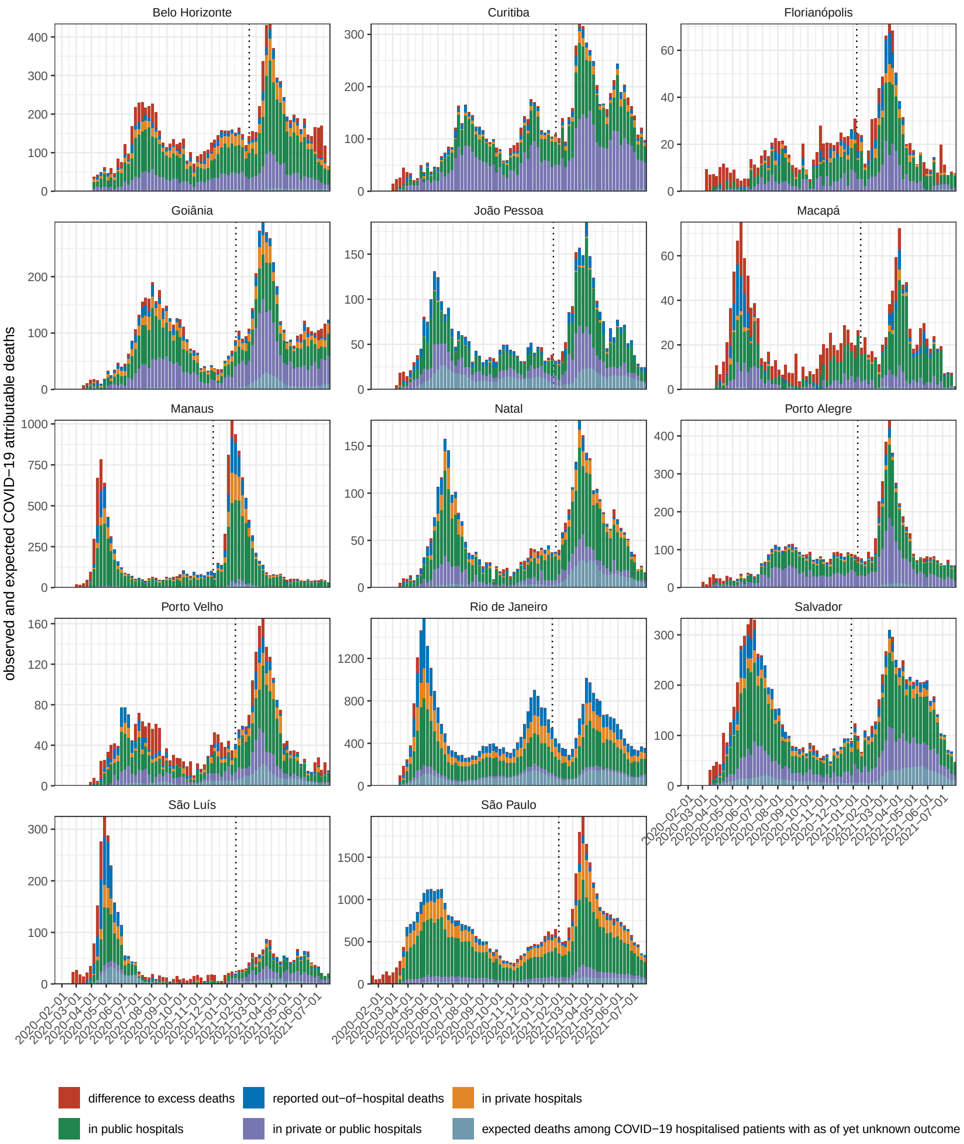


**Figure S3:** Underreporting-adjusted COVID-19 attributable deaths. Reported COVID-19 attributable deaths in the SIVEP-Gripe platform were adjusted for in-hospital underreporting, by counting a proportion of hospitalised patients with as of yet unknown outcome as fatal, and for likely out-of-hospital under-reporting, by comparison against population excess deaths derived from all-cause mortality data of the Brazilian Civil Registry (Supplementary Text, page 24). The date of Gamma's first detection in each city is shown as a vertical dotted line.

**A**

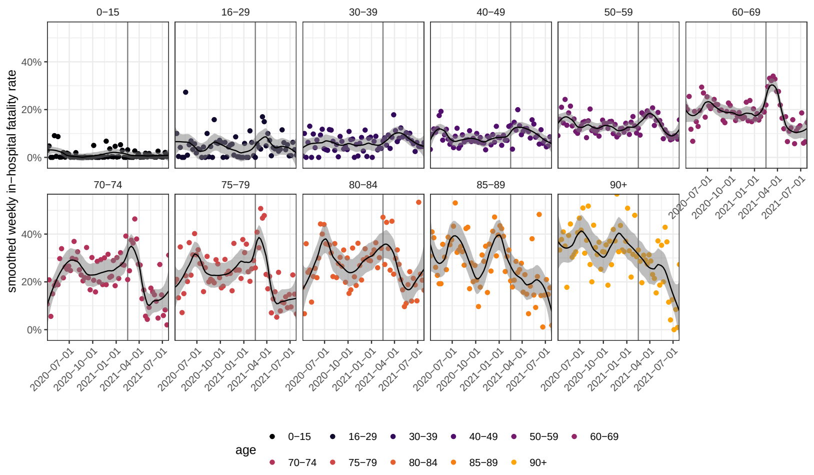


**B**

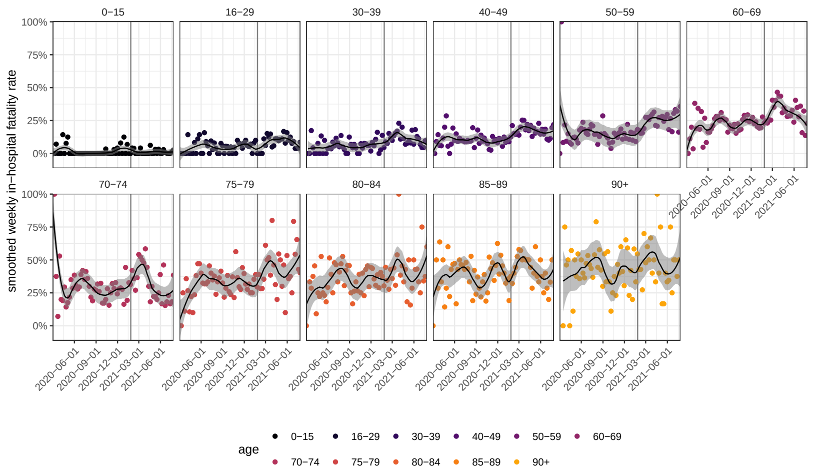


**Figure S4:** Time trends in age-specific COVID-19 in-hospital fatality rates. Weekly, age-specific COVID-19 in-hospital fatality rates are shown as dots, and non-parametric loess estimates of time trends are shown as block solid line along with 95% confidence intervals as grey ribbon. The date of Gamma's first detection is indicated as a vertical dotted black line. (A) For Belo Horizonte. (B) For Curitiba.

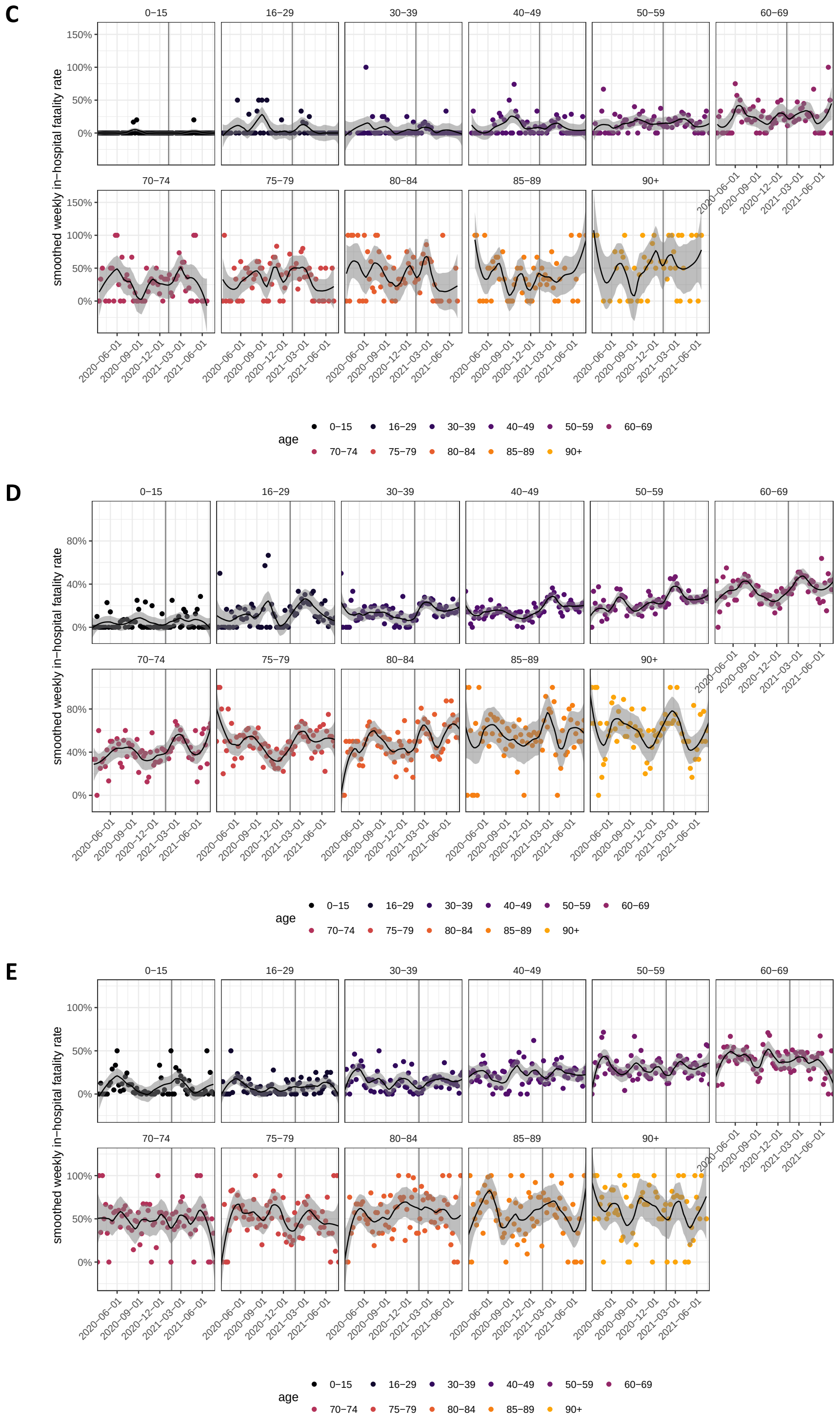


**Figure S4:** (continued) Time trends in age-specific COVID-19 in-hospital fatality rates. (C) For Florianópolis. (D) For Goiânia. (E) For João Pessoa.

**F**

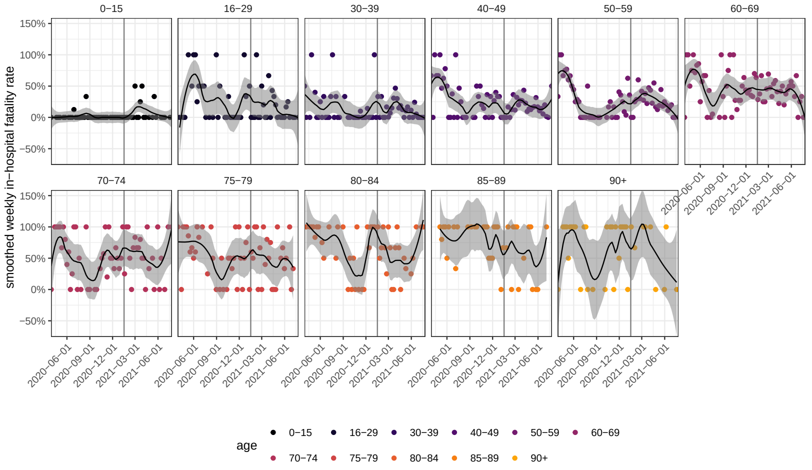


**G**

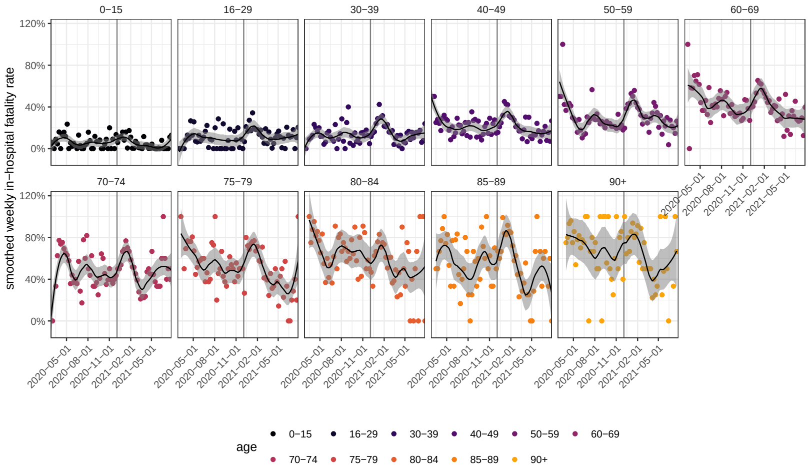


**H**

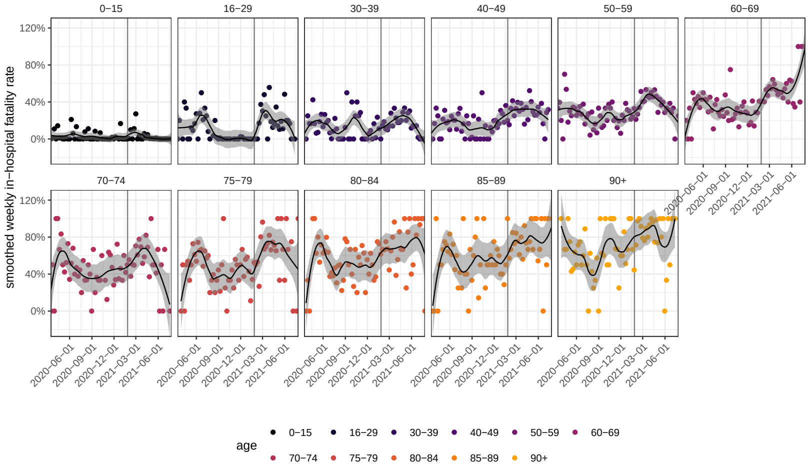


**Figure S4:** (continued) Time trends in age-specific COVID-19 in-hospital fatality rates. (F) For Macapá. (G) For Manaus. (H) For Natal.

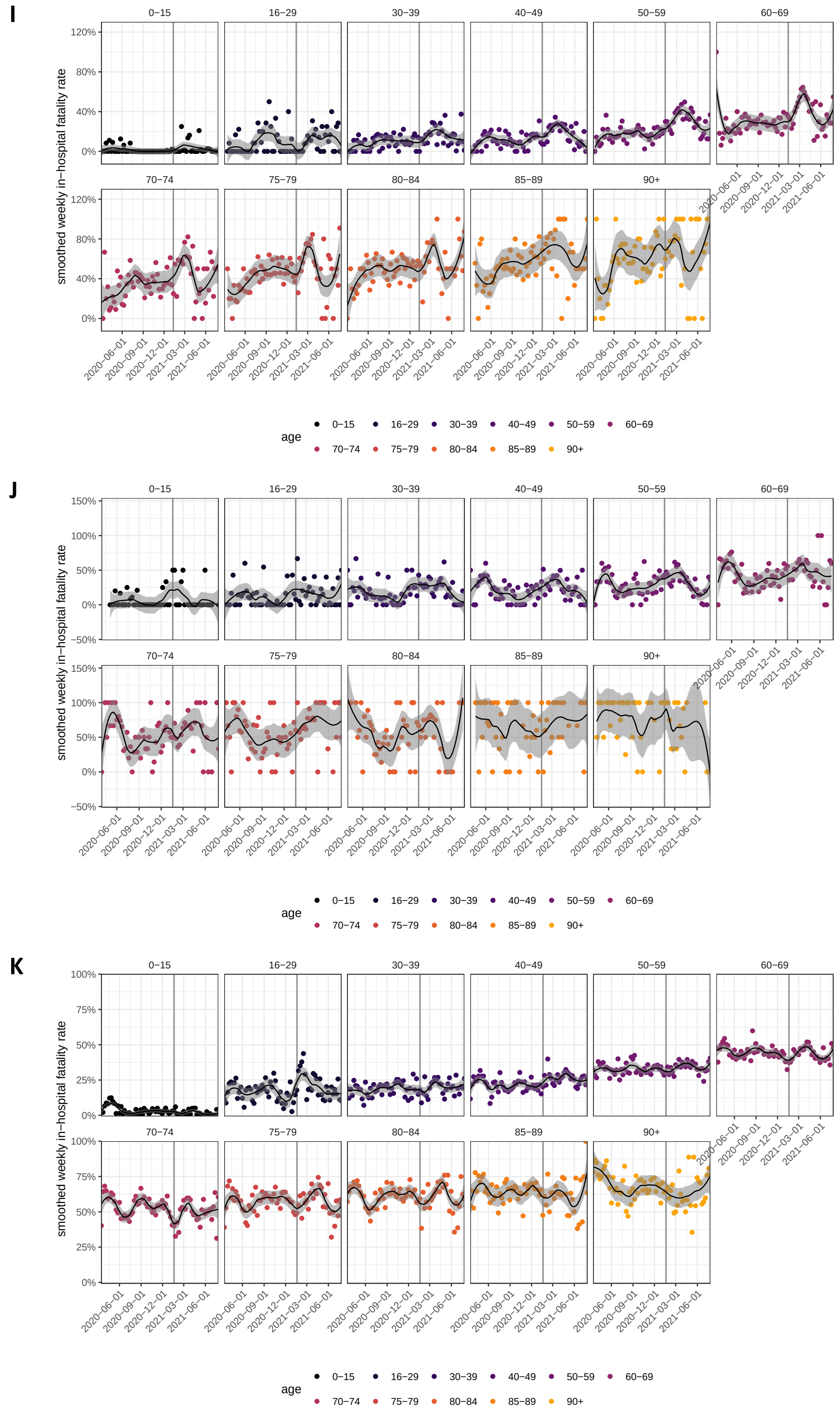


**Figure S4:** (continued) Time trends in age-specific COVID-19 in-hospital fatality rates. (I) For Porto Alegre. (J) For Porto Velho. (K) For Rio De Janeiro.

L

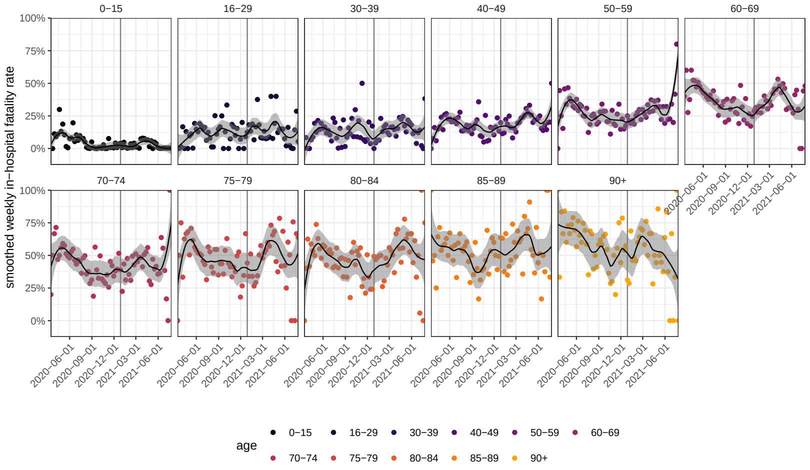


M

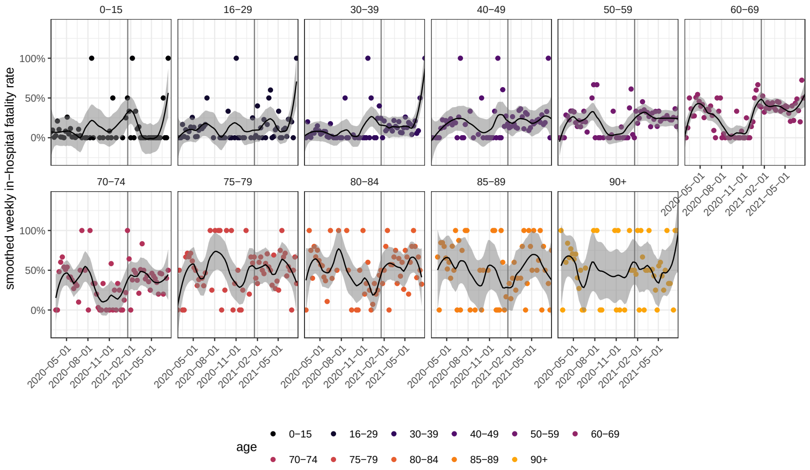


N

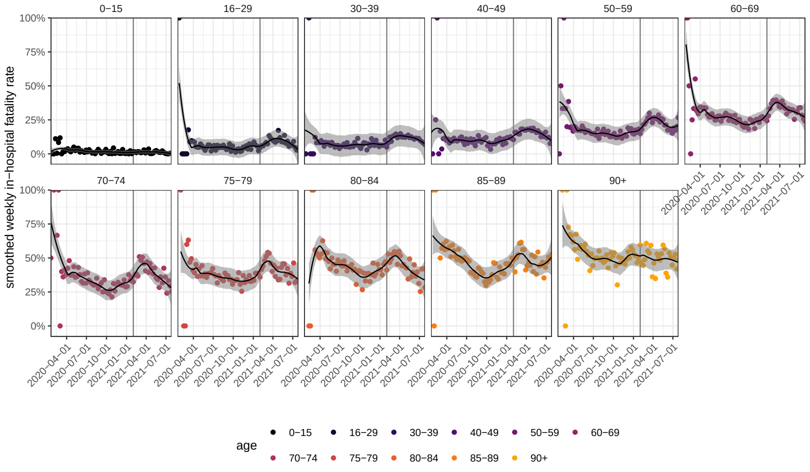


**Figure S4:** (continued) Time trends in age-specific COVID-19 in-hospital fatality rates. (L) For Salvador. (M) For São Luís. (N) For São Paulo city.

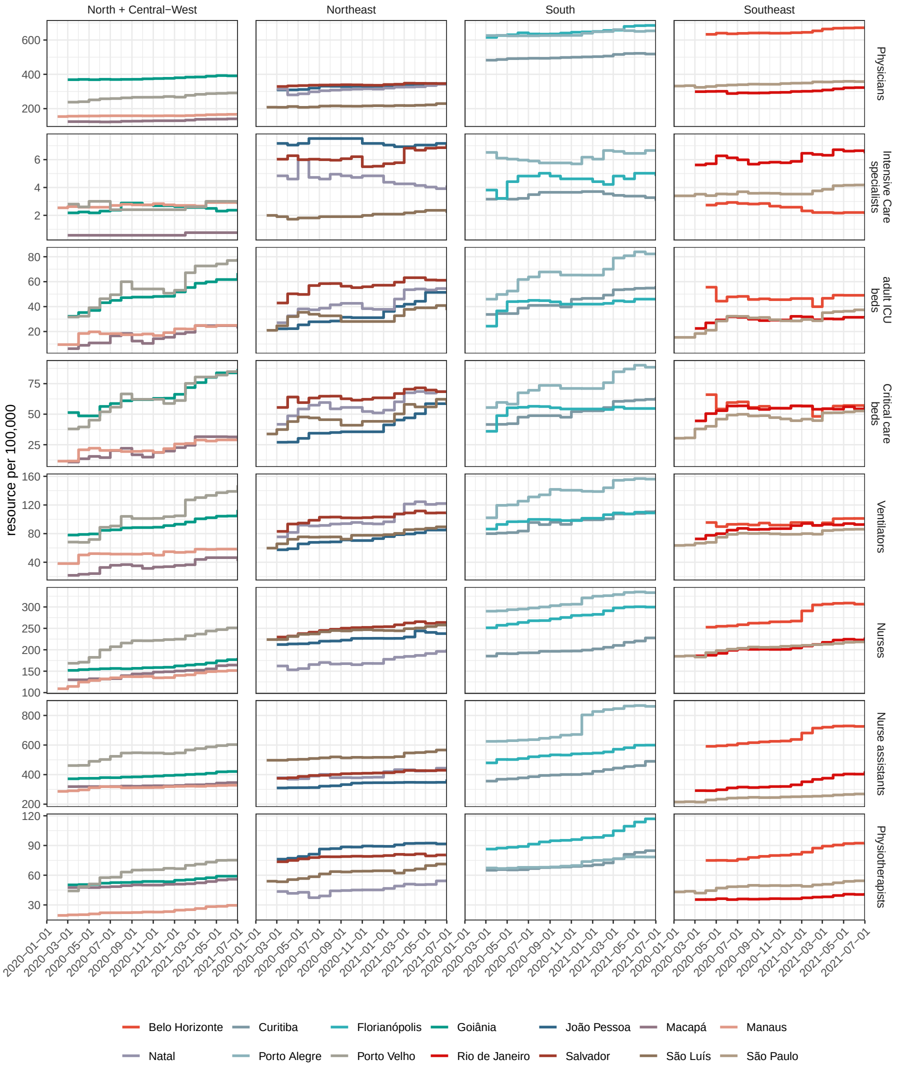


**Figure S5:** Healthcare resources in hospital settings, per 100,000. Monthly data on health care resources in each location were obtained from the National Register of Health Facilities (Cadastro Nacional de Estabelecimentos de Saúde - CNES), and were aggregated from facility-level microdata to city-level as described in Supplementary Text, page 14.

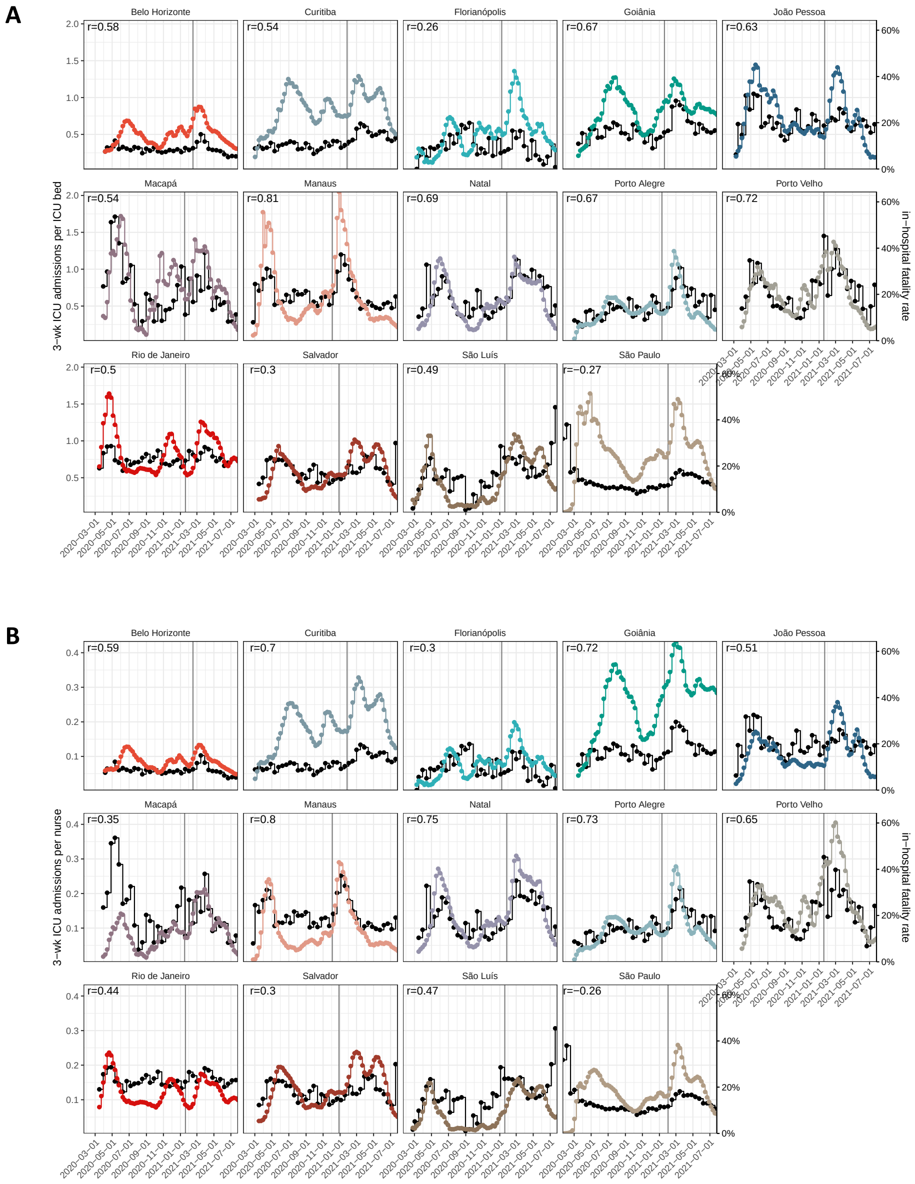


**Figure S6:** Time evolution of pandemic healthcare pressure indices. (A) ICU admissions in this and the following two weeks per ICU bed with ventilator are shown in colour, with y-axis on the left. The biweekly, empirical age-standardised in-hospital fatality rates are shown in black, with y-axis on the right. Pearson correlation coefficients ($r$) are shown in the upper left corner, and dates of Gamma's first detection as vertical dotted lines. (B) ICU admissions in this and the following two weeks per nurse.

**C**

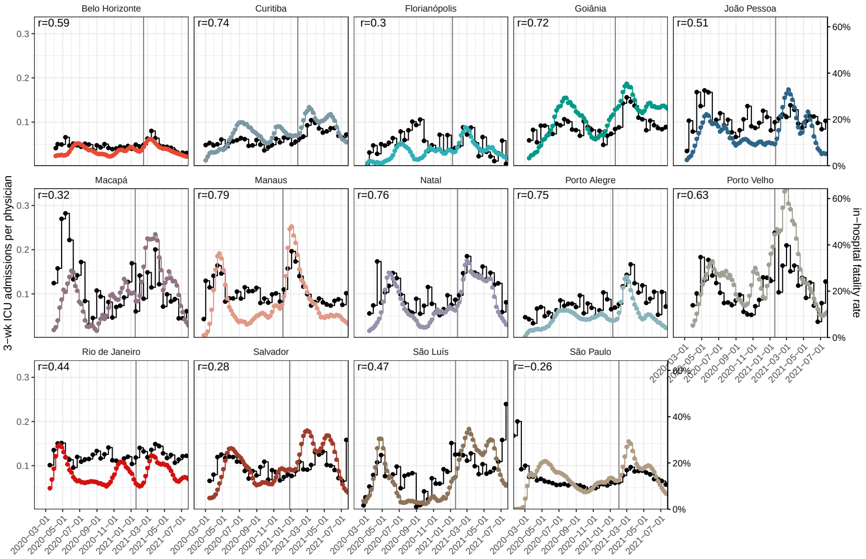


**D**

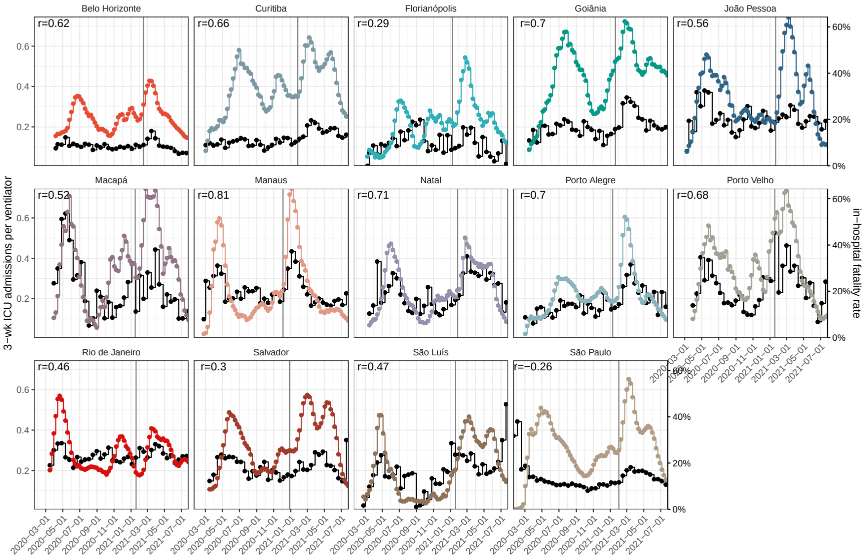


**Figure S6:** (continued) (C) ICU admissions in this and the following two weeks per physician. (D) ICU admissions in this and the following two weeks per ventilator.

**E**

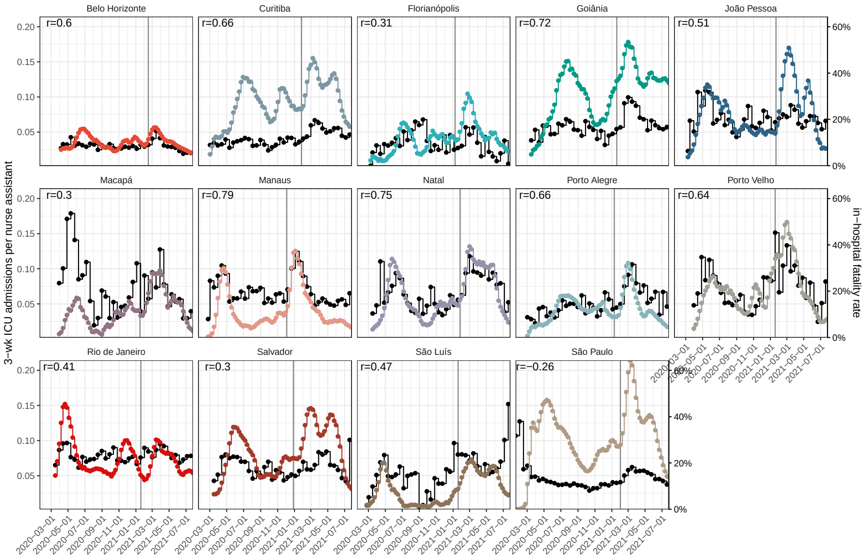


**F**

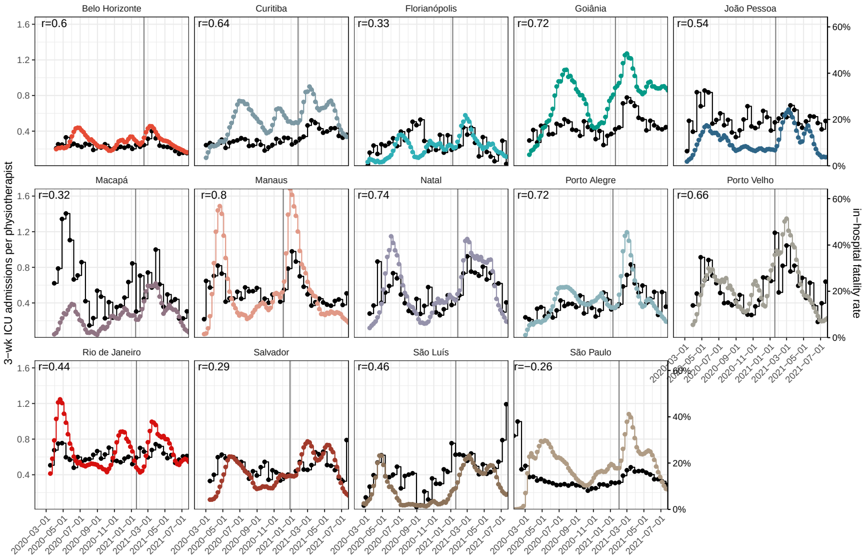


**Figure S6:** (continued) (E) ICU admissions in this and the following two weeks per nurse assistant. (F) ICU admissions in this and the following two weeks per physiotherapist.

**G**

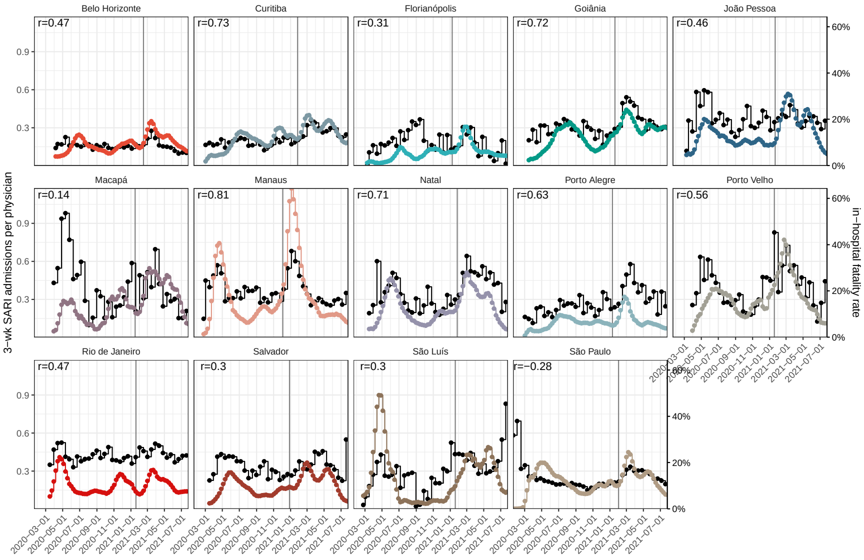


**H**

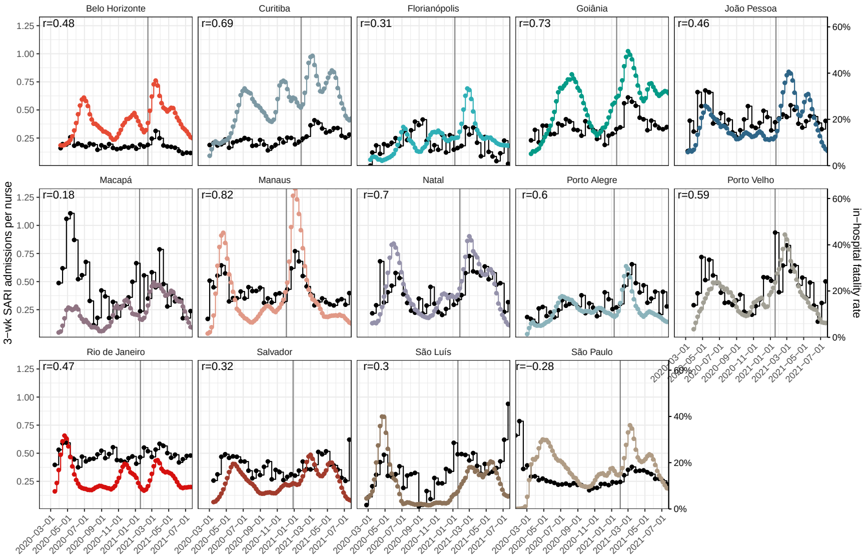


**Figure S6:** (continued) (G) SARI admissions in this and the following two weeks per physician. (H) SARI admissions in this and the following two weeks per nurse.

I

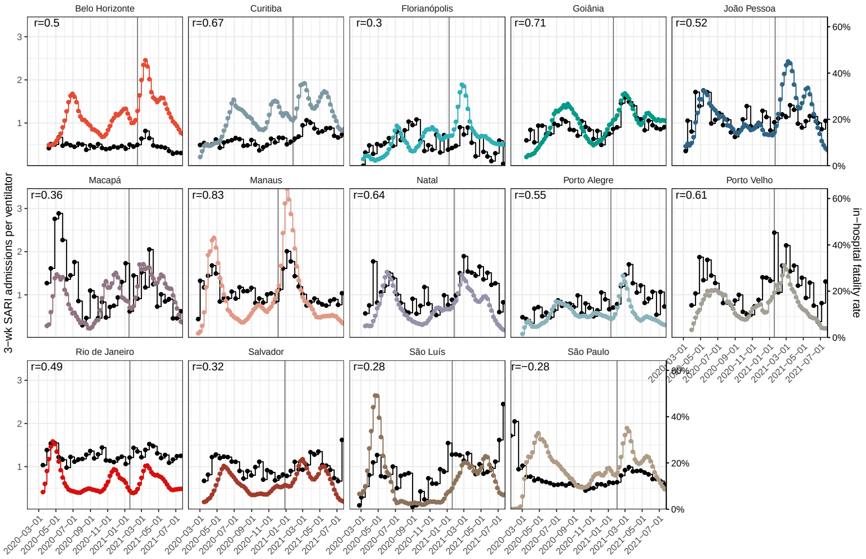


J

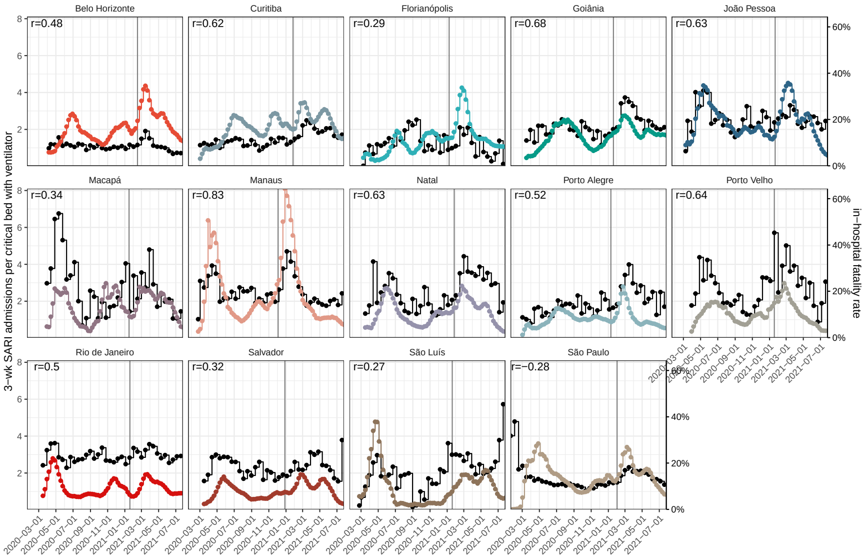


**Figure S6:** (continued) (I) SARI admissions in this and the following two weeks per ventilator. (J) SARI admissions in this and the following two weeks per ICU and intermediate beds with ventilator.

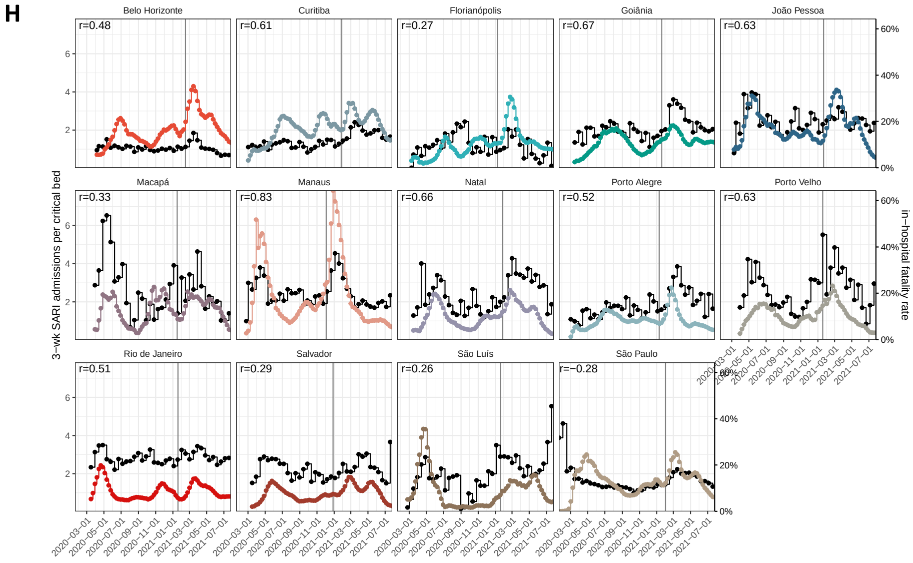


**Figure S6:** (continued) (I) SARI admissions in this and the following two weeks per ventilator. (J) SARI admissions in this and the following two weeks per ICU and intermediate beds with or without ventilator.

**A**

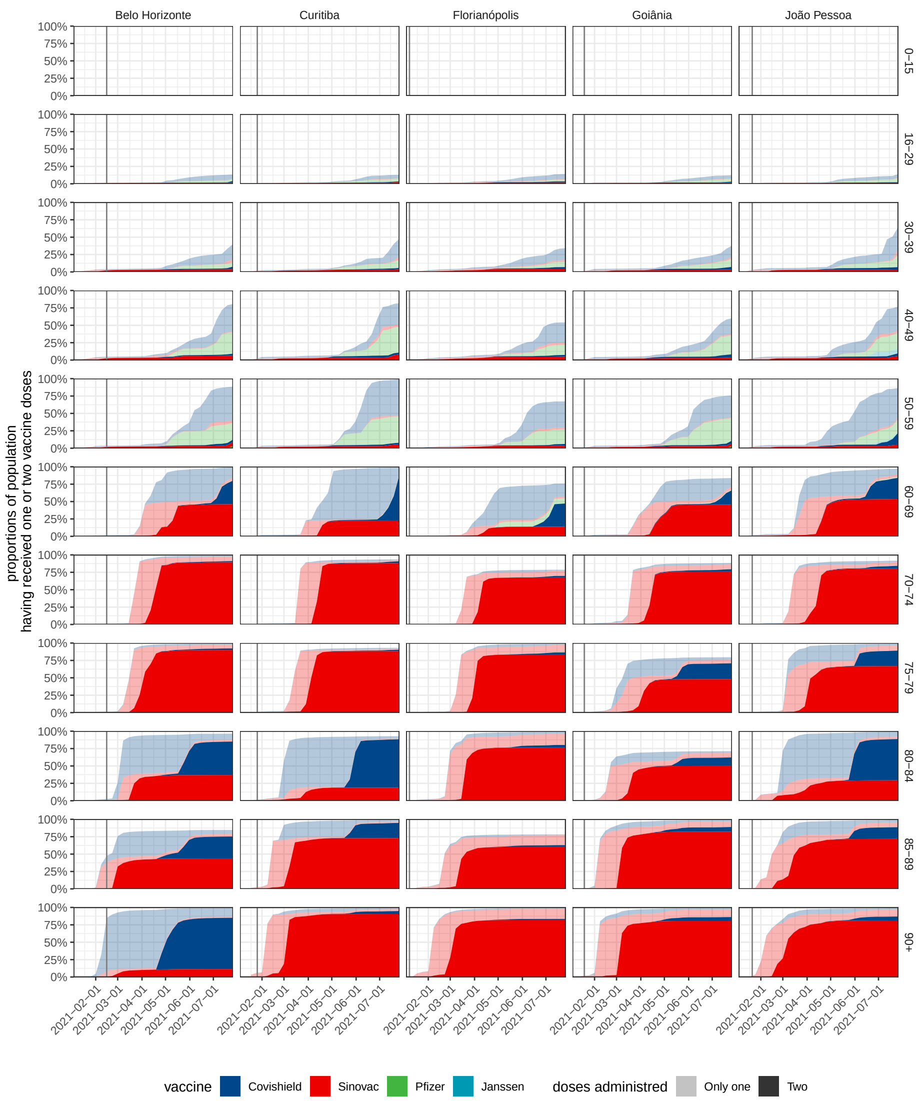


**Figure S7:** COVID-19 vaccine coverage. Individual-level data on administered vaccine doses from the Brazilian Ministry of Health database [2] were retrieved on 05 August 21, and preprocessed as described in the Supplementary Text, page 19 and 32. Estimated vaccine coverage is shown by vaccine (colour) and number of doses administered (colour intensity). The date of Gamma's detection is added as a grey vertical line. (A) Data for Belo Horizonte, Curitiba, Florianópolis, Goiânia, João Pessoa.

**B**

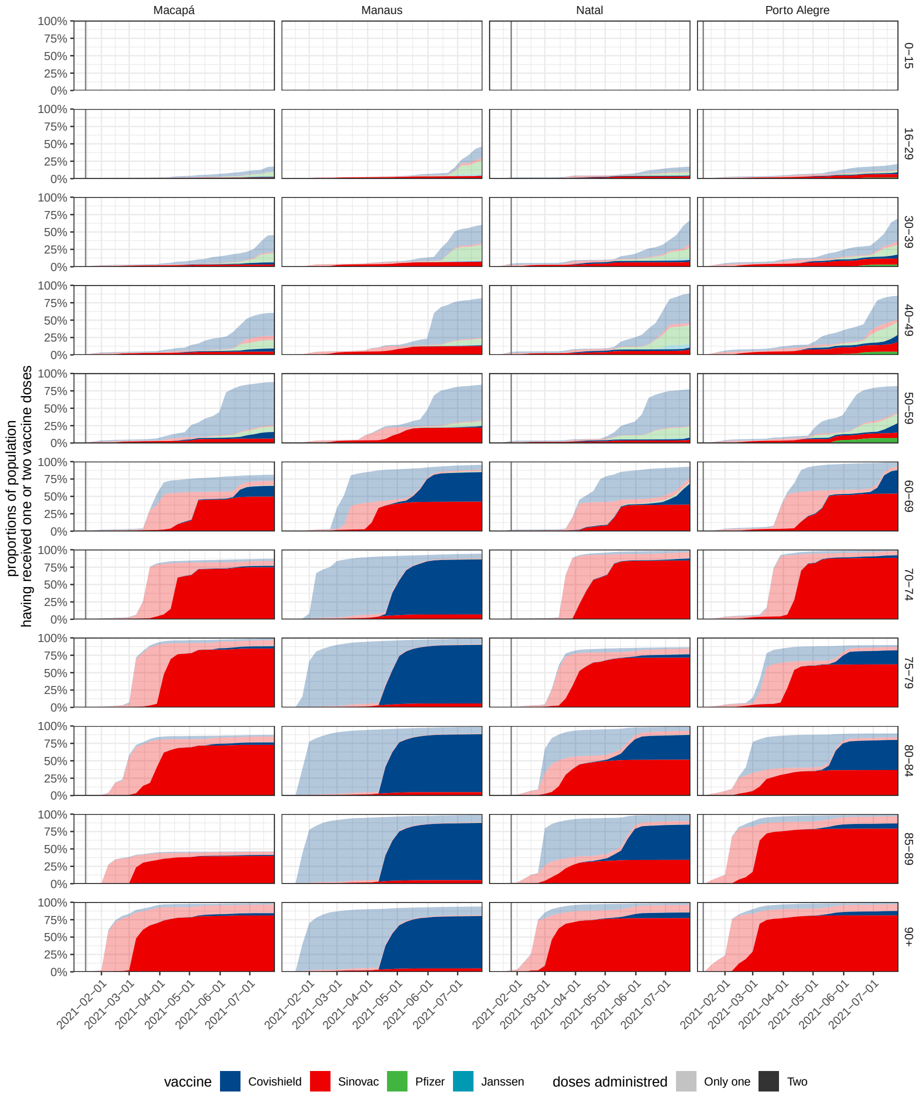


**Figure S7:** (continued) (B) Estimated COVID-19 vaccine coverage in Macapá, Manaus, Natal, Porto Alegre.

C

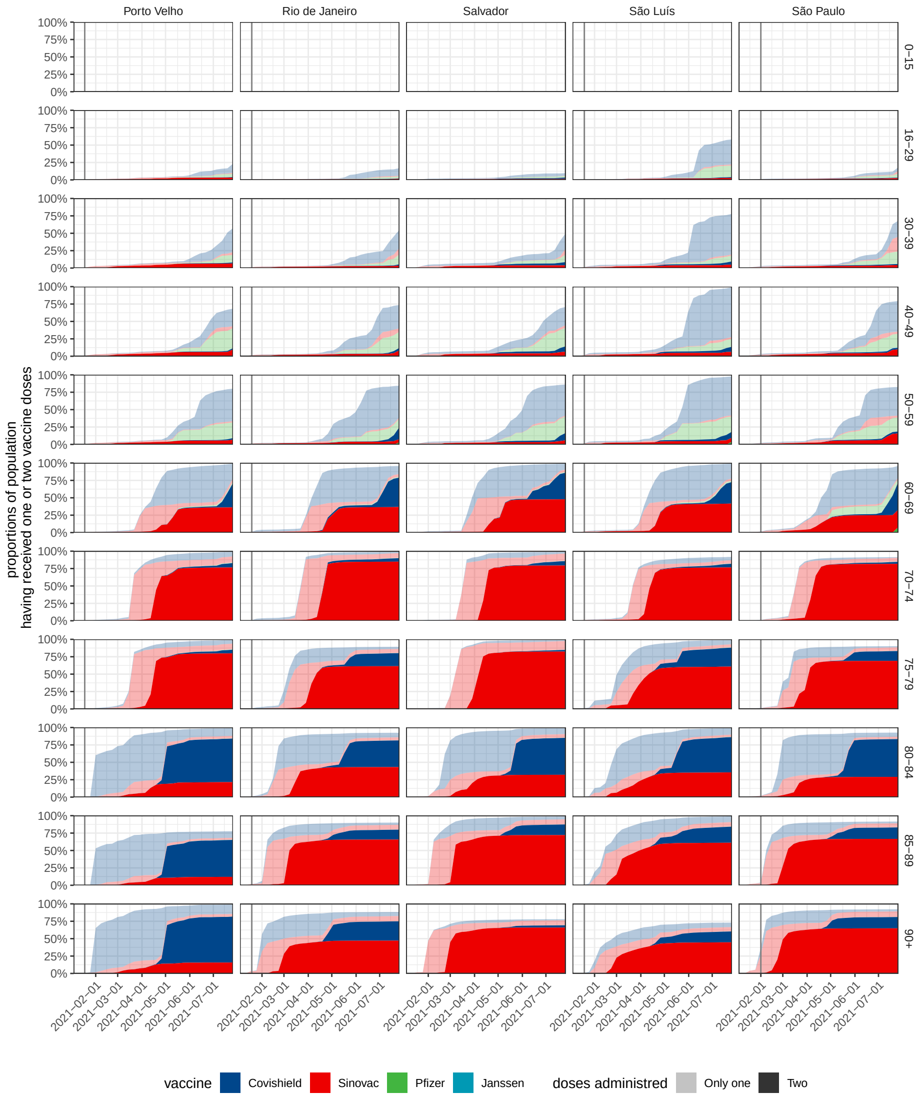


**Figure S7:** (continued) (C) Estimated COVID-19 vaccine coverage in Porto Velho, Rio De Janeiro, Salvador, São Luís, São Paulo city.

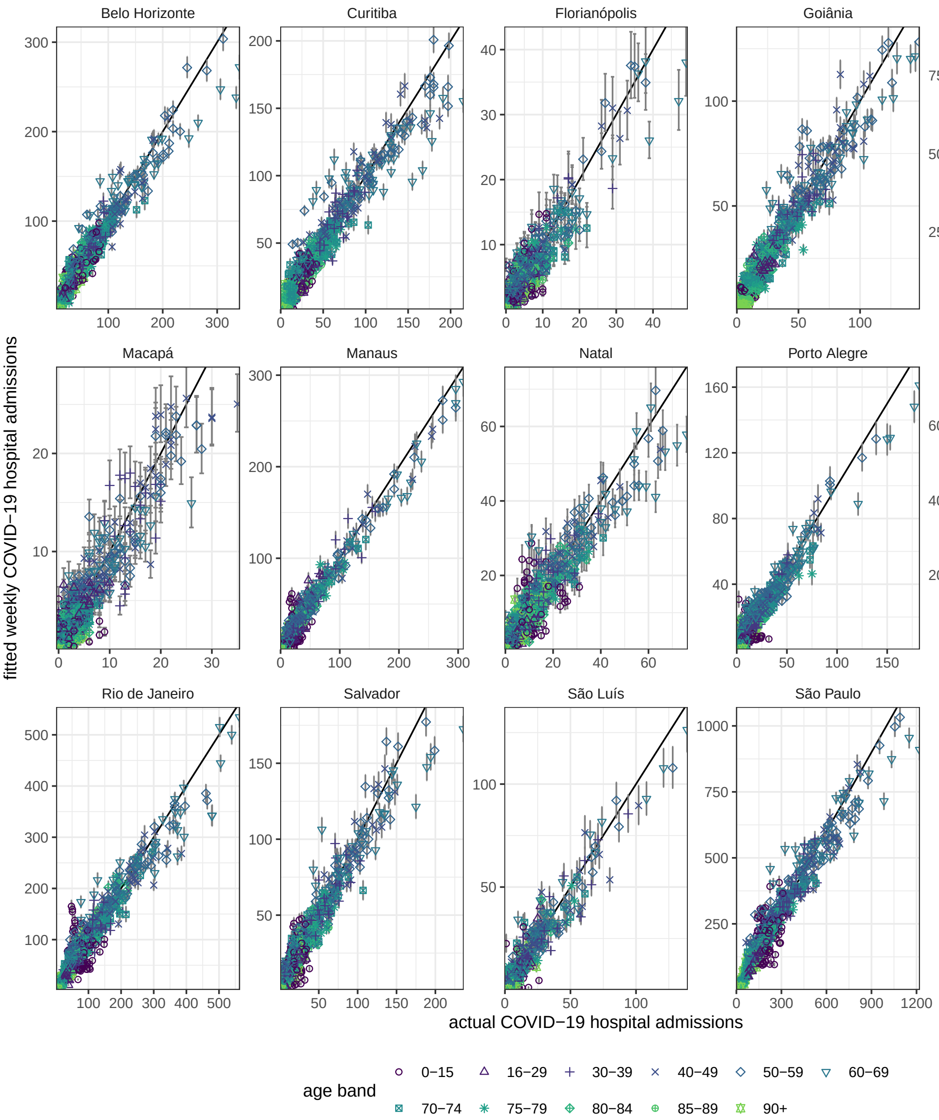


**Figure S8:** Model fit of expected weekly COVID-19 attributable hospital admissions. Posterior median estimates of the expected hospital admissions in residents in location $l$ and age band $a$ in week $w$ obtained with the Bayesian multi-strain fatality model, Supplementary Text Equation (S37), are shown on the y-axis along with 95% credible intervals against the observed, COVID-19 attributable hospital admissions among residents in location $l$ and age band $a$ in week $w$ defined in Supplementary Text, page 13. Locations are shown across facets, and age bands are shown using colours and plot symbols.

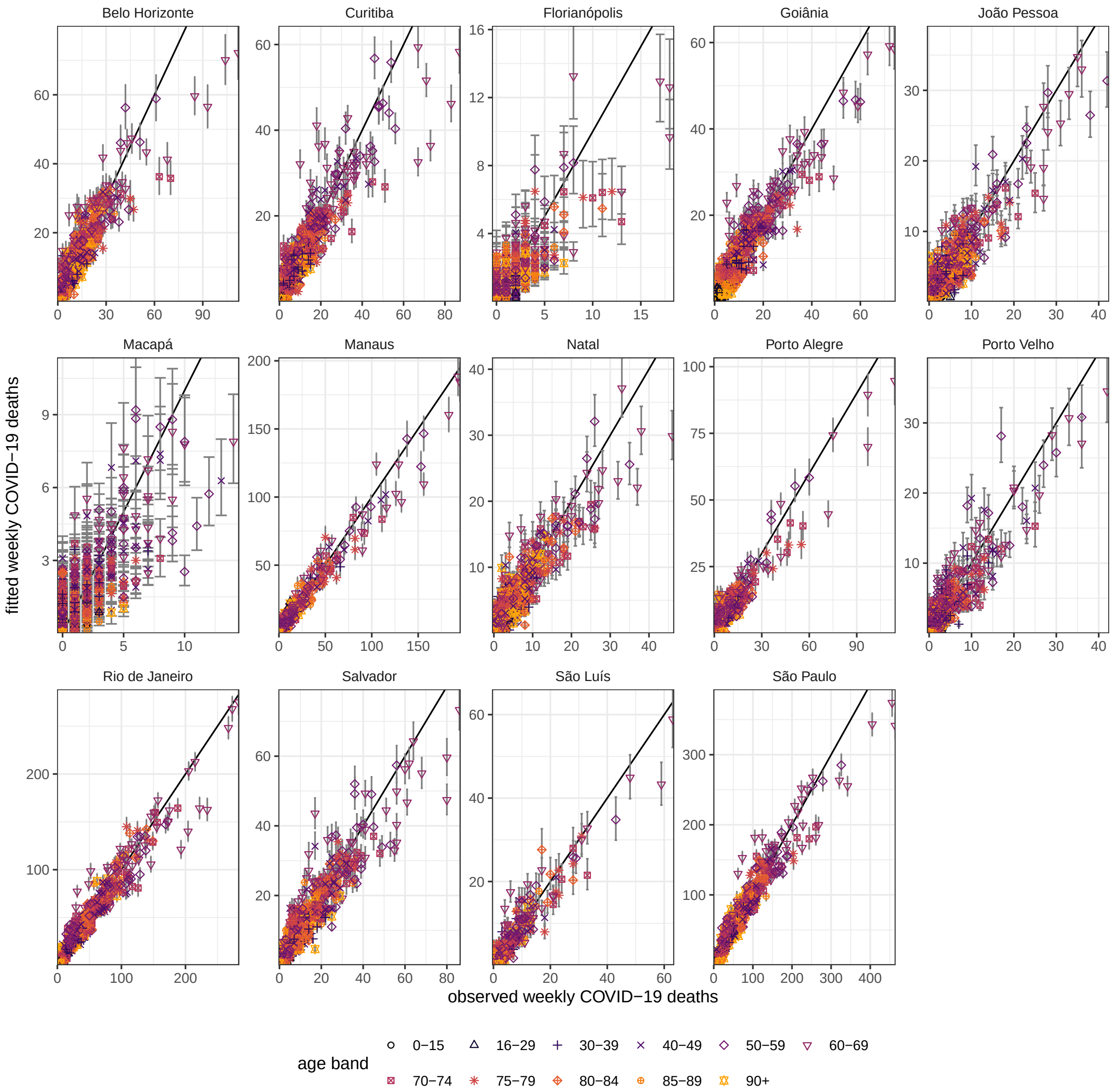


**Figure S9:** Model fit of expected weekly COVID-19 attributable deaths. Posterior median estimates of the expected deaths in location $l$ and age band $a$ following hospital admission of residents in week $w$ obtained with the Bayesian multi-strain fatality model, Supplementary Text Equation (S40), are shown on the y-axis along with 95% credible intervals against the underreporting-adjusted COVID-19 attributable deaths following hospital admission, Supplementary Text Equation (S11), on the x-axis. Locations are shown across facets, and age bands are shown using different colours and plot symbols.

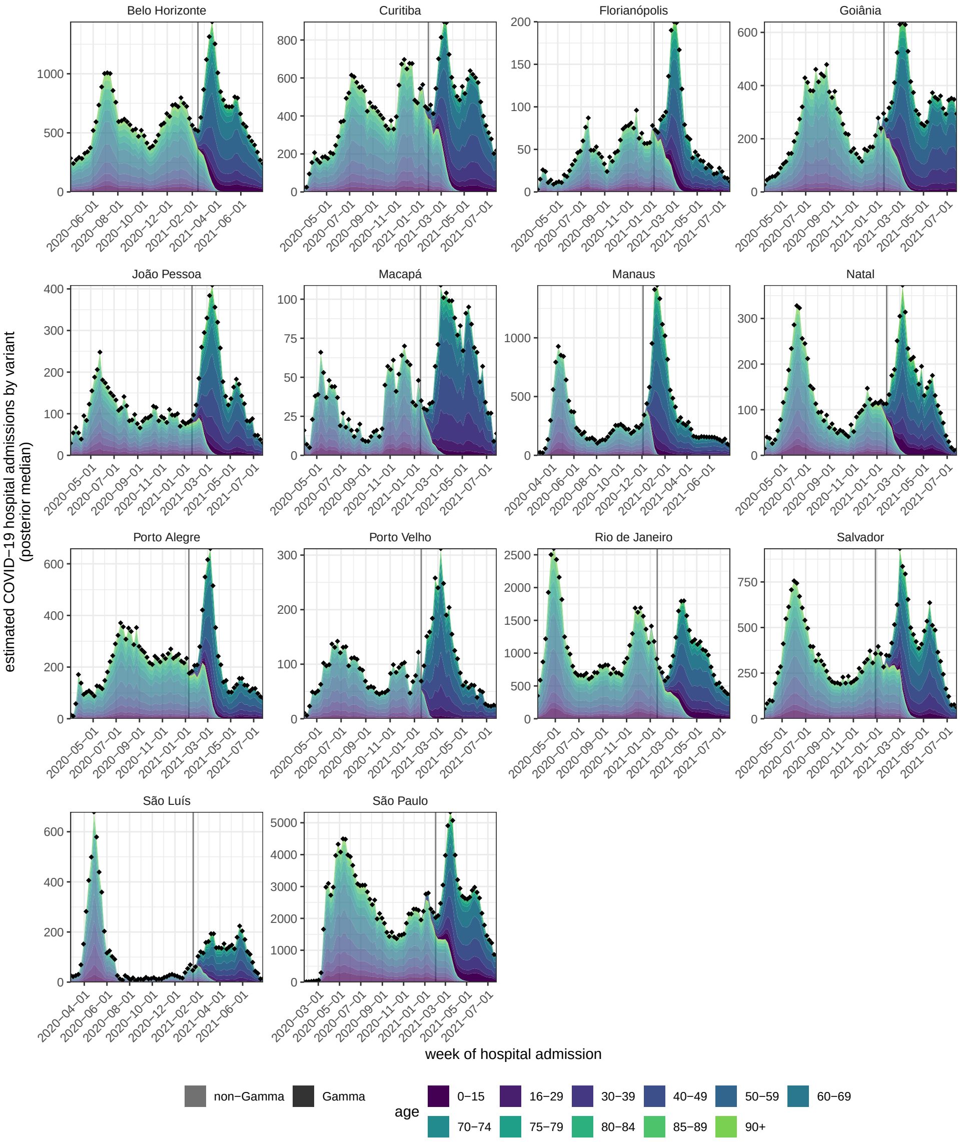


**Figure S10:** Estimated COVID-19 attributable hospital admissions by SARS-Cov-2 variant. Posterior median estimates of hospital admissions among residents in each location, that are attributed to non-Gamma variants are shown for each age band (color) in lighter shades, while those for the Gamma variant are shown in darker shades. Estimates are derived using the Bayesian multi-strain fatality model, Supplementary Text page 42. Locations are shown across facets. The date of Gamma's detection is shown as a vertical dotted line. Observed weekly hospital admissions in residents are shown as black dots.

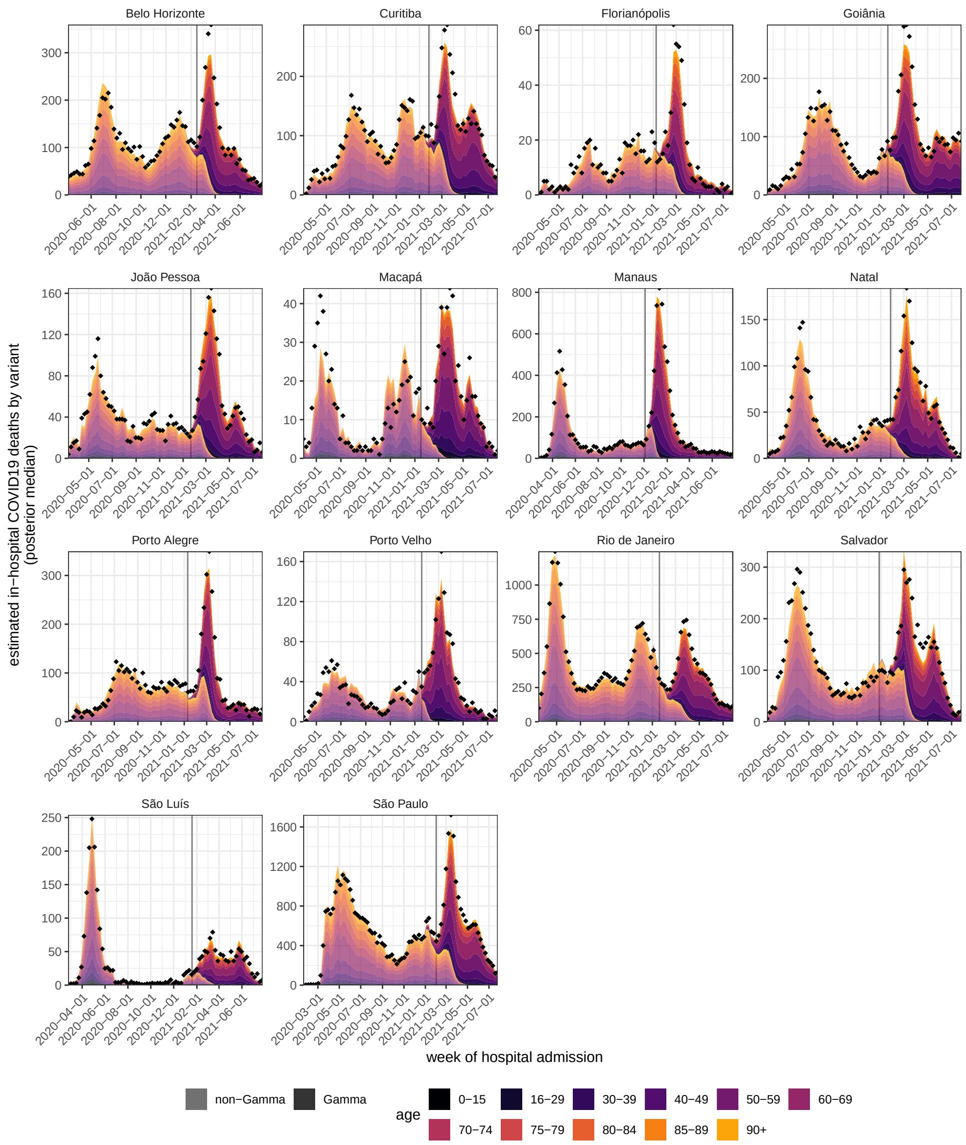


**Figure S11:** Estimated COVID-19 attributable deaths in hospitals by SARS-Cov-2 variant. Posterior median estimates of deaths following hospital admissions of residents in each location, that are attributed to non-Gamma variants are shown for each age band (color) in lighter shades, while those for the Gamma variant are shown in darker shades. Estimates are derived using the Bayesian multi-strain fatality model, Supplementary Text page 42. Locations are shown across facets. The date of Gamma's detection is shown as a vertical dotted line. Observed weekly deaths following hospital admissions in residents are shown as black dots.

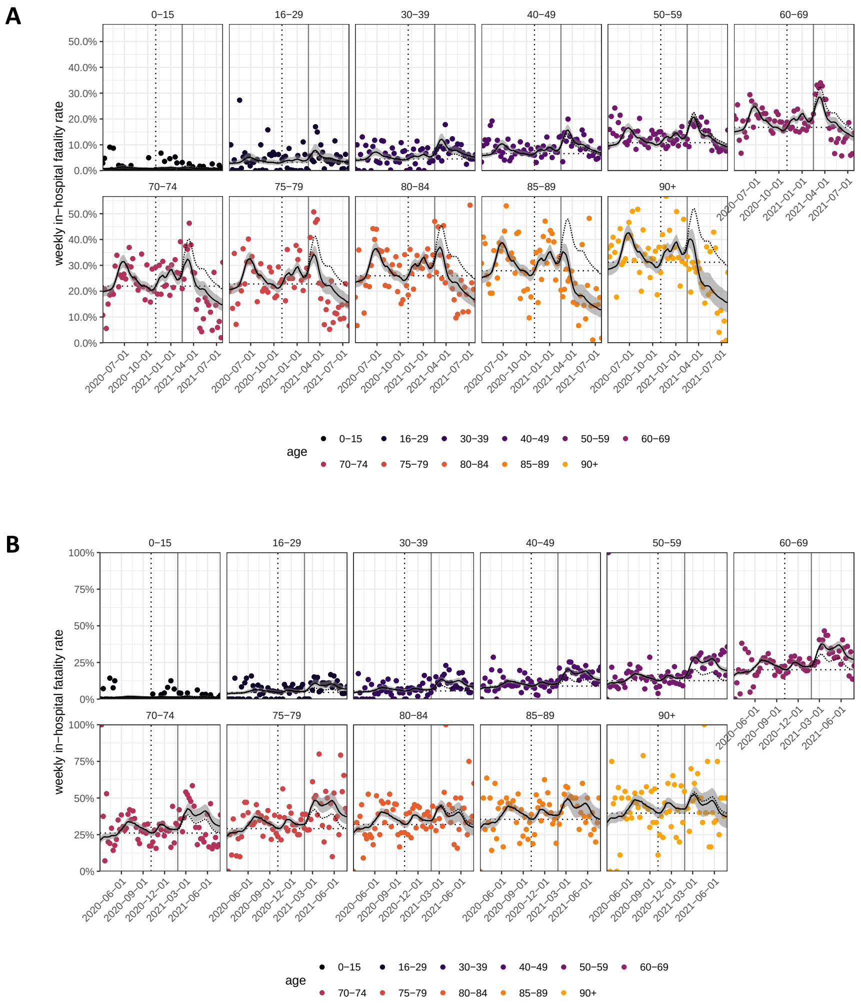


**Figure S12:** Model fits to age-specific COVID-19 in-hospital fatality rates. Weekly, age-specific in-hospital fatality rates are shown as dots. Posterior median estimates of the expected in-hospital fatality rates across variants from the Bayesian multi-strain fatality model, Supplementary Text Equation (S34), are shown on the y-axis (black line) along with 95% credible intervals (grey ribbon). The expected in-hospital fatality rates of non-Gamma variants, Supplementary Text Equation (S33a), are shown as dotted line. The date of Gamma's first detection is indicated as a vertical dotted black line. (A) For Belo Horizonte. (B) For Curitiba.

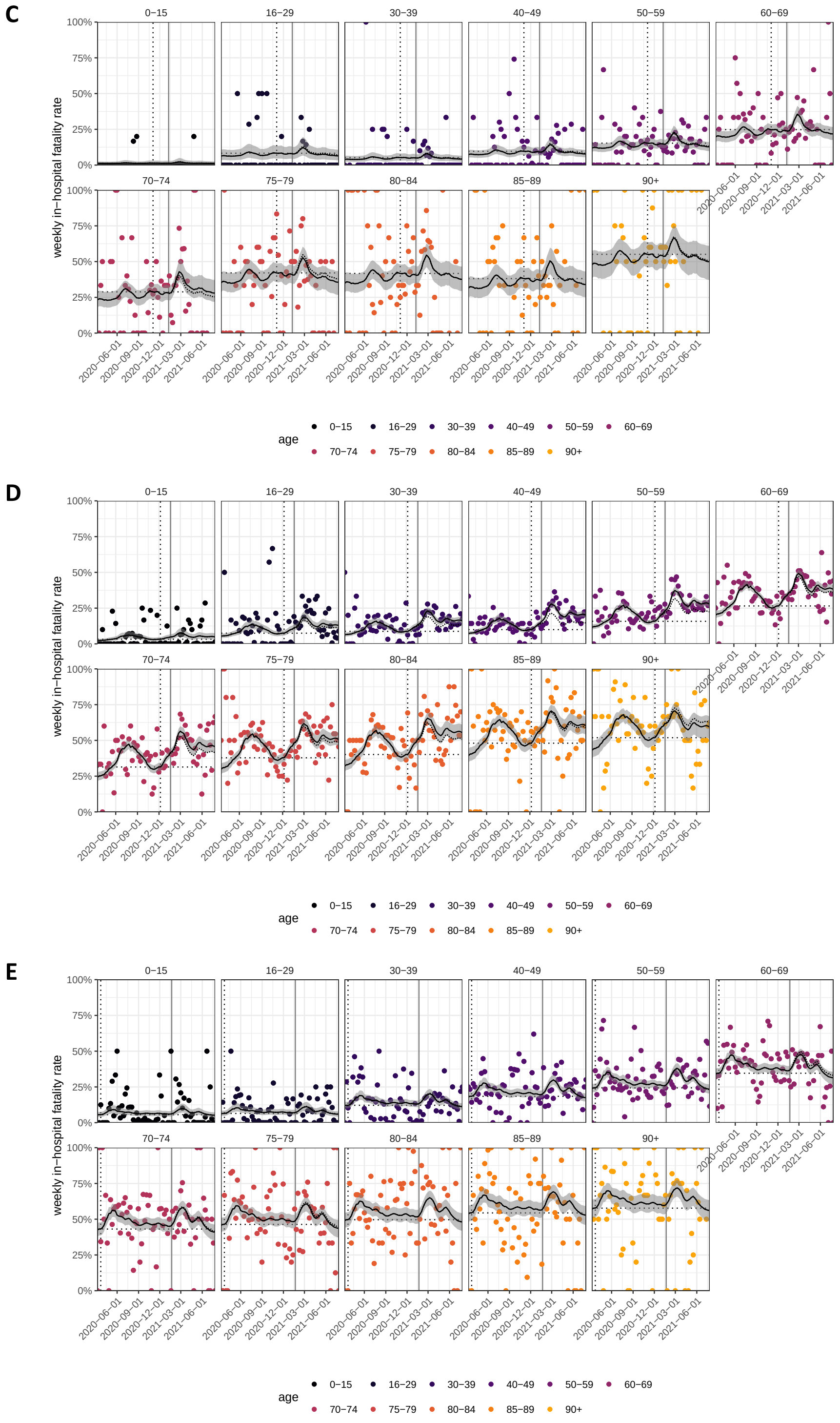


**Figure S12:** (continued) Model fits to age-specific COVID-19 in-hospital fatality rates. (C) For Florianópolis. (D) For Goiânia. (E) For João Pessoa.

DOI: `https://doi.org/10.25561/91875` 

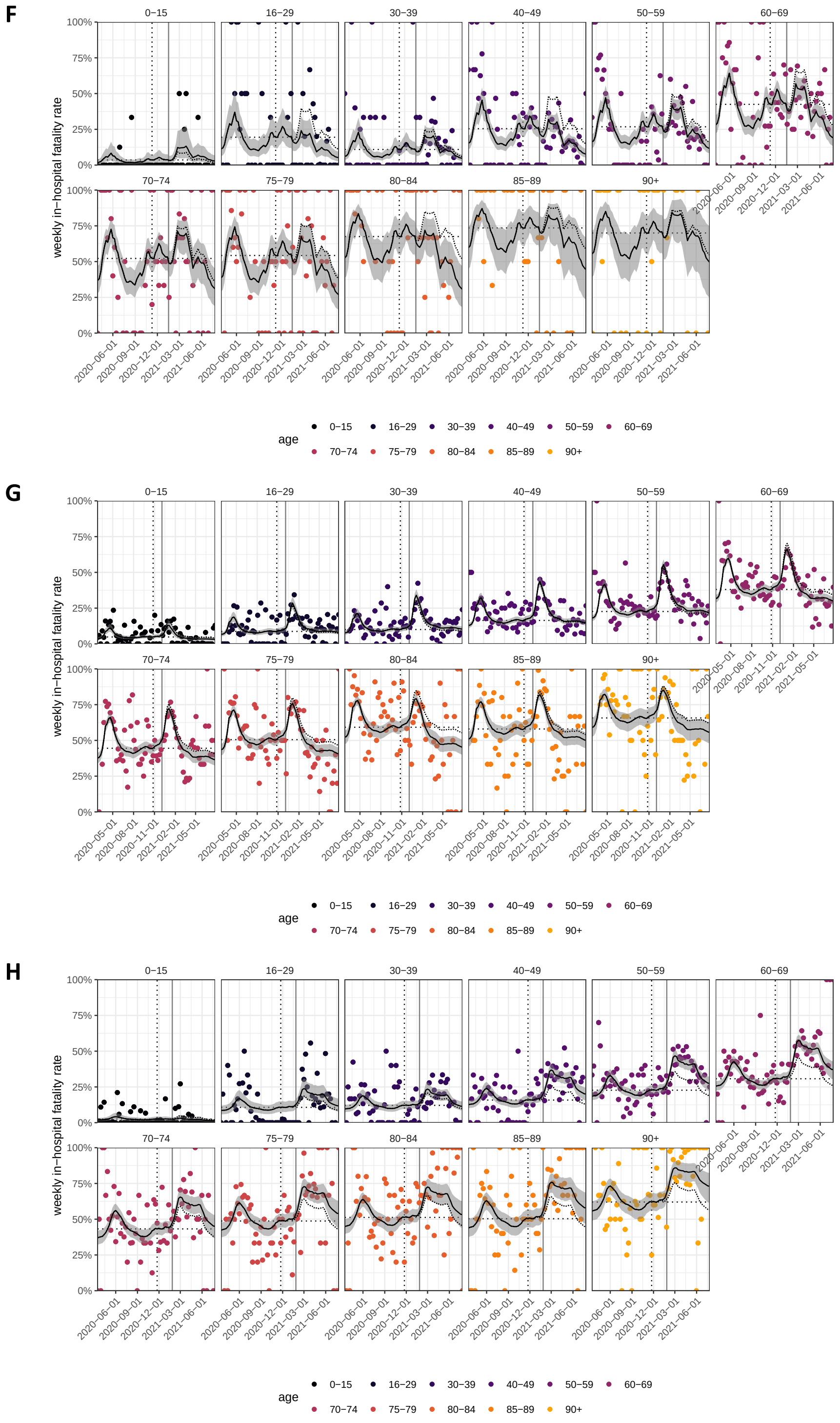


**Figure S12:** (continued) Model fits to age-specific COVID-19 in-hospital fatality rates. (F) For Macapá. (G) For Manaus. (H) For Natal.

I

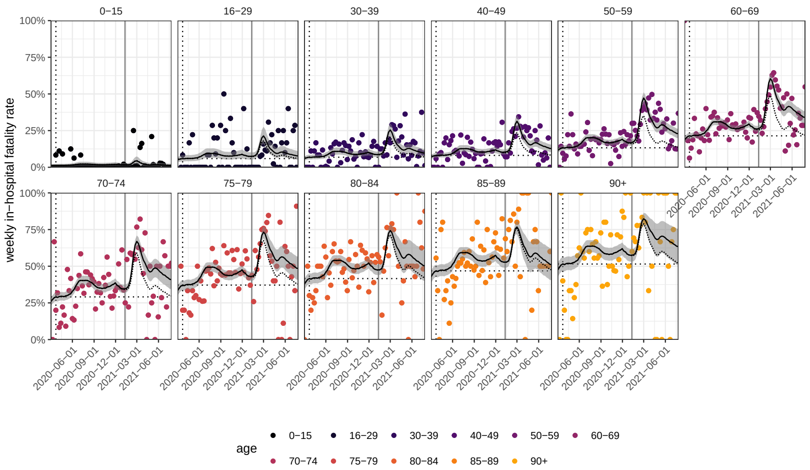


J

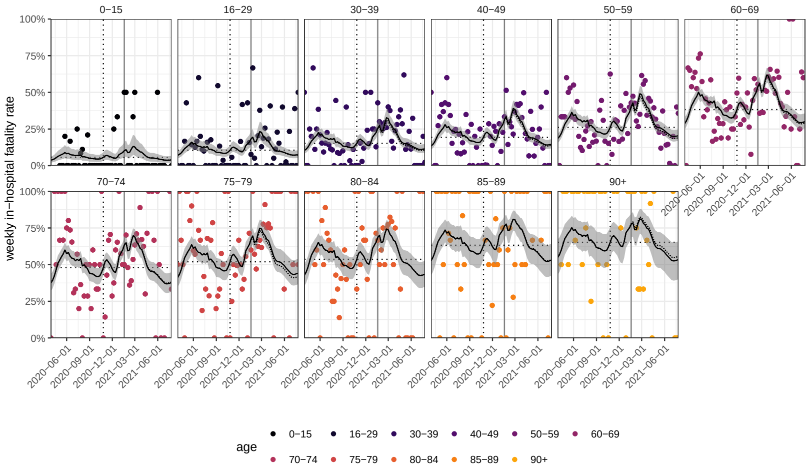


K

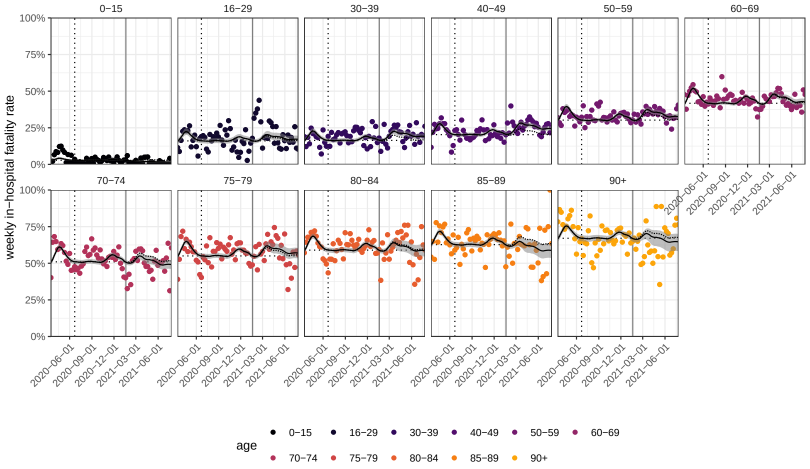


**Figure S12:** (continued) Model fits to age-specific COVID-19 in-hospital fatality rates. (I) For Porto Alegre. (J) For Porto Velho. (K) For Rio De Janeiro.

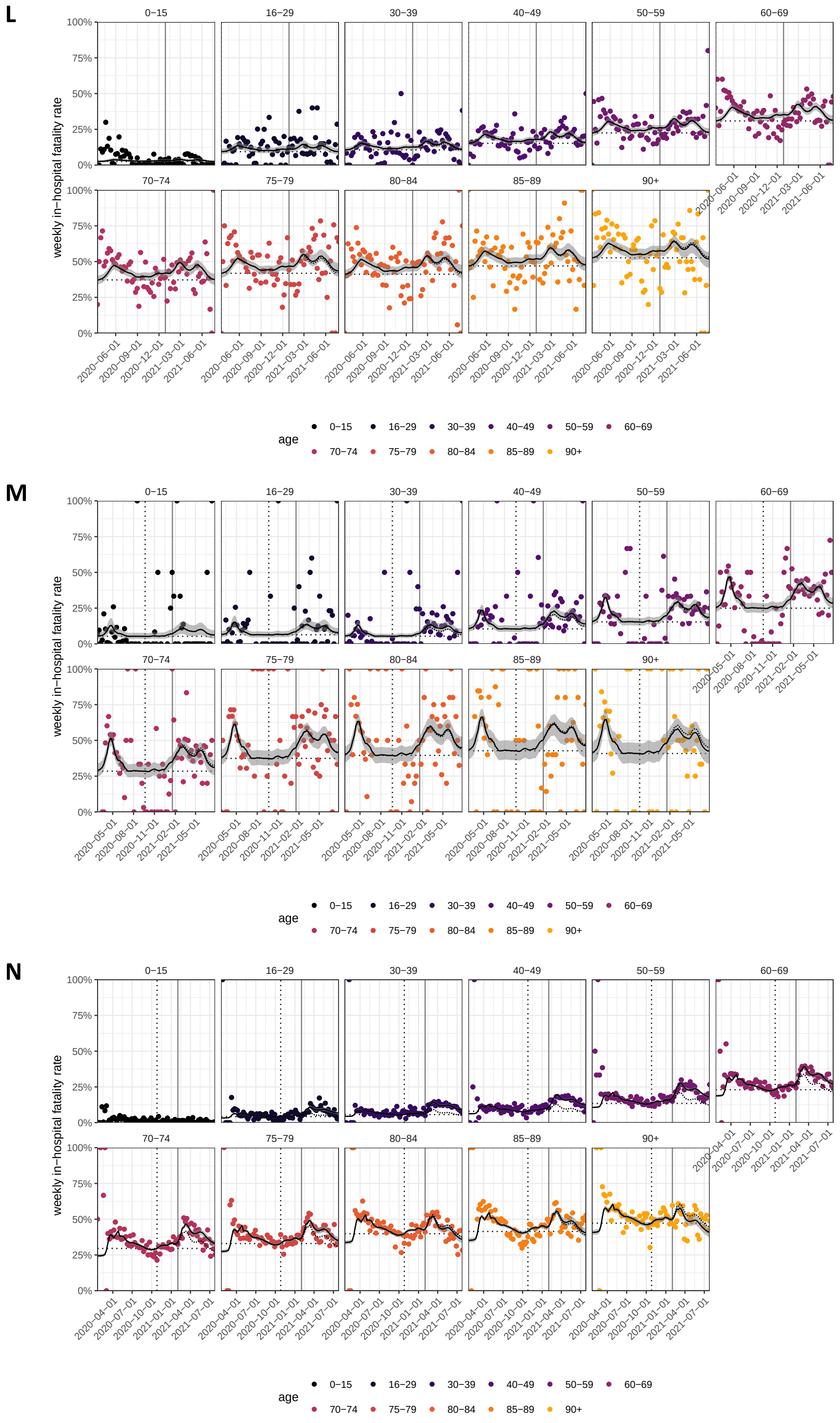


**Figure S12:** (continued) Model fits to age-specific COVID-19 in-hospital fatality rates. (L) For Salvador. (M) For São Luís. (N) For São Paulo city.

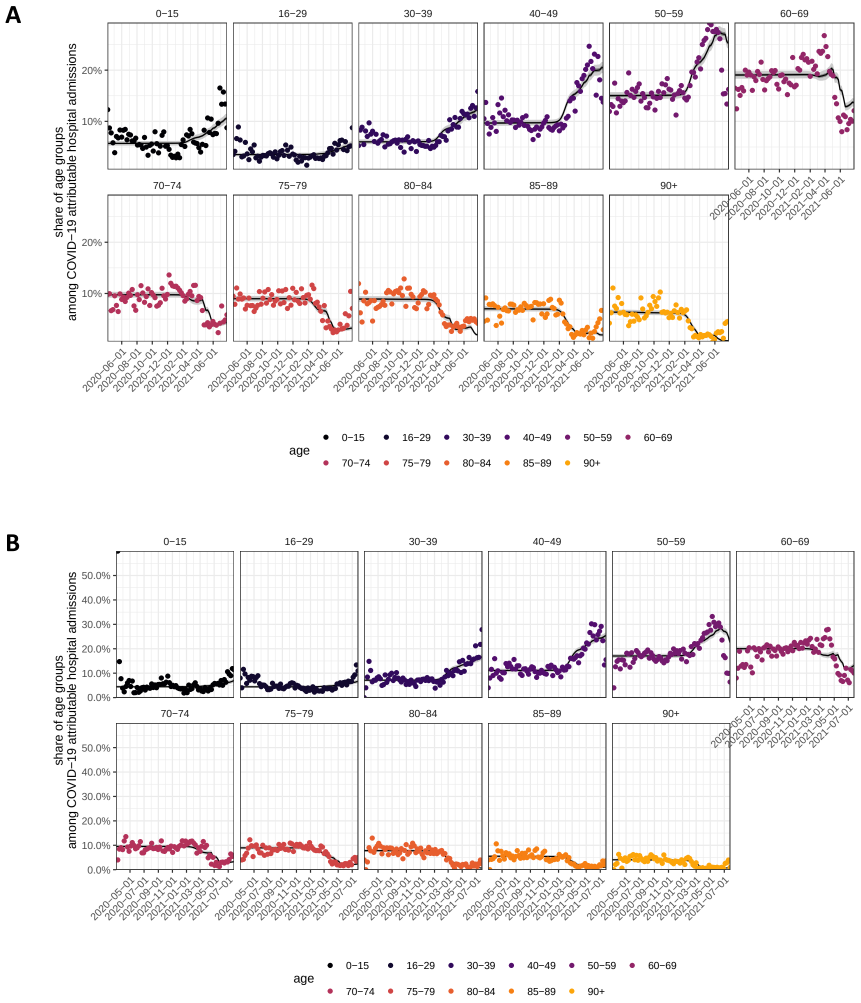


**Figure S13:** Model fits of the expected age composition of COVID-19 attributable hospital admissions. Posterior median estimates of the expected age composition of hospital admissions in each location, age band, and week, obtained with the Bayesian multi-strain fatality model, Supplementary Text Equation (S31b), are shown on the y-axis as a black line along with 95% credible intervals. Time trends are shown by week of hospital admission (x-axis). The empirical proportions are shown as dots. (A) For Belo Horizonte. (B) For Curitiba.

C

D

E

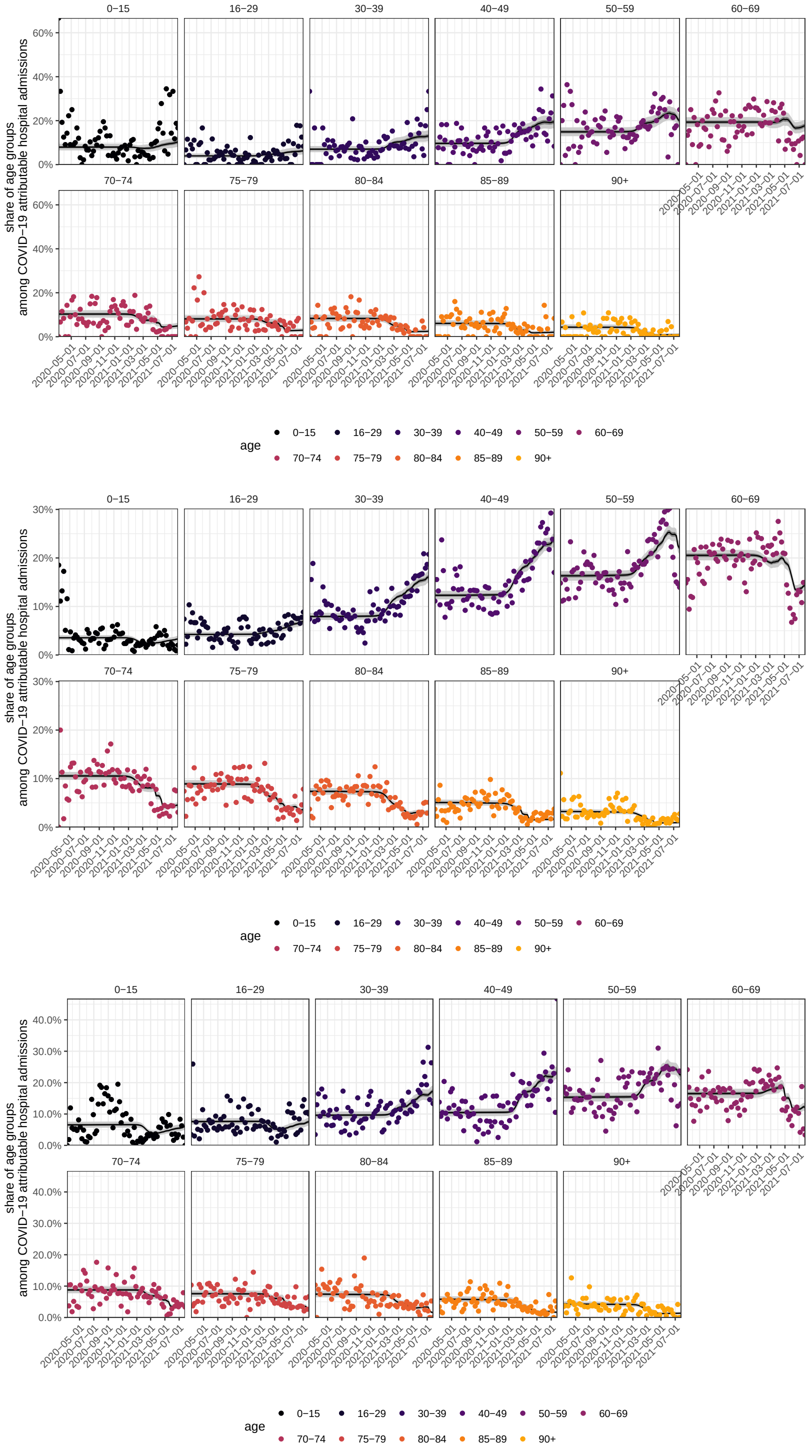


**Figure S13:** (continued) Model fits of the expected age composition of COVID-19 attributable hospital admissions. (C) For Florianópolis. (D) For Goiânia. (E) For João Pessoa.

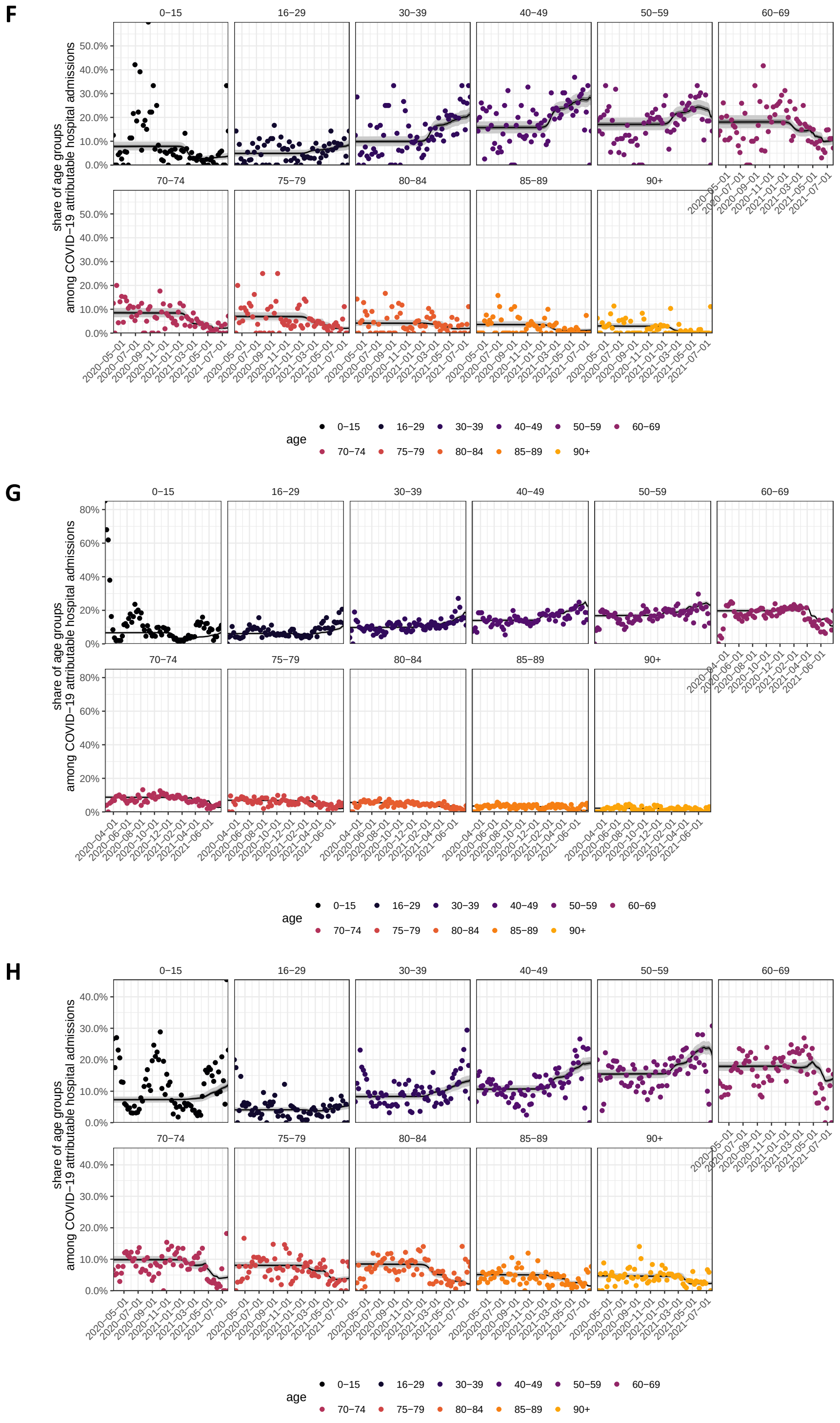


**Figure S13:** (continued) Model fits of the expected age composition of COVID-19 attributable hospital admissions. (F) For Macapá. (G) For Manaus. (H) For Natal.

**I**

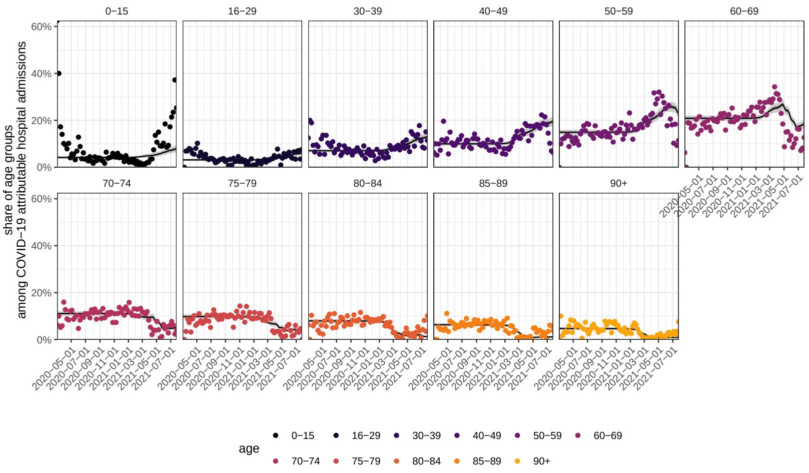


**J**

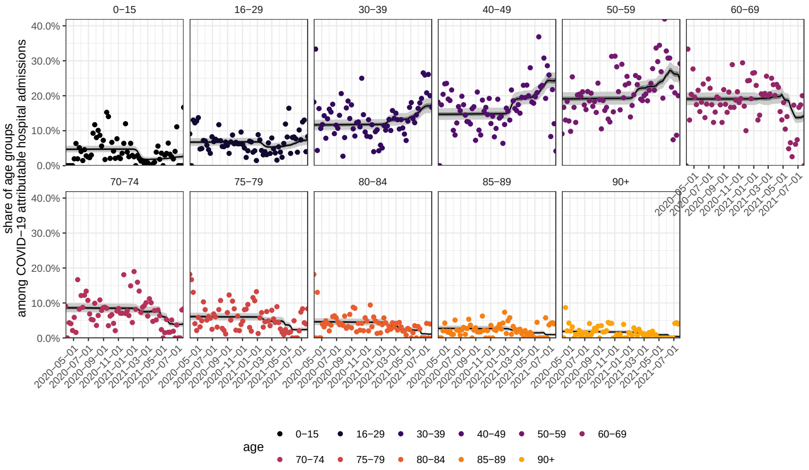


**K**

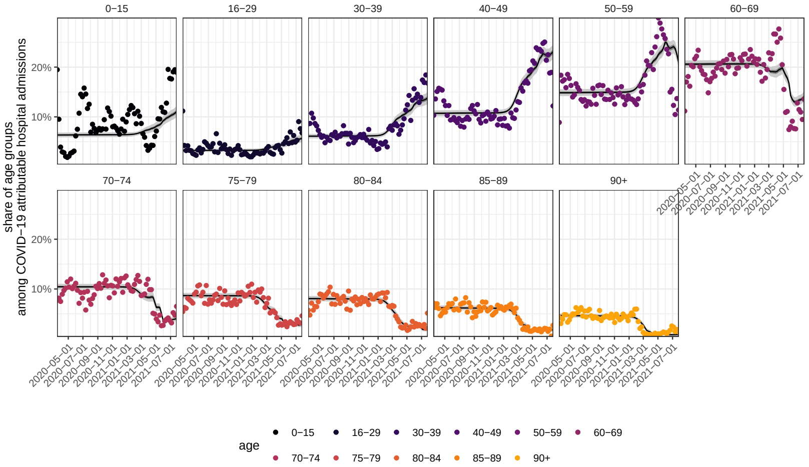


**Figure S13:** (continued) Model fits of the expected age composition of COVID-19 attributable hospital admissions. (I) For Porto Alegre. (J) For Porto Velho. (K) For Rio De Janeiro.

**L**

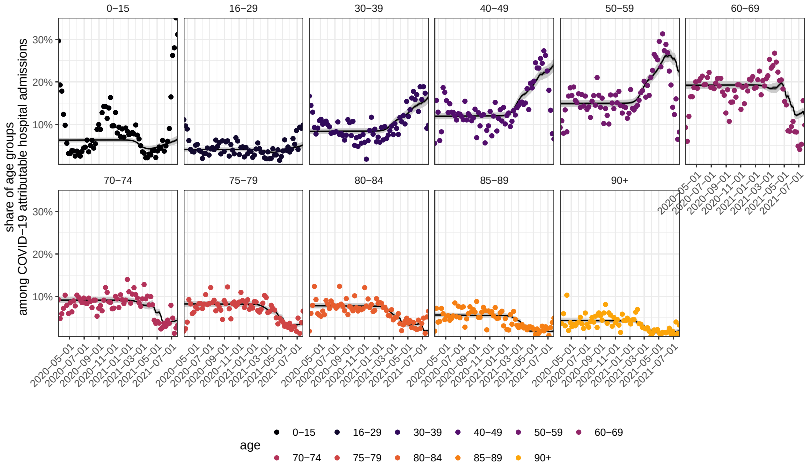


**M**

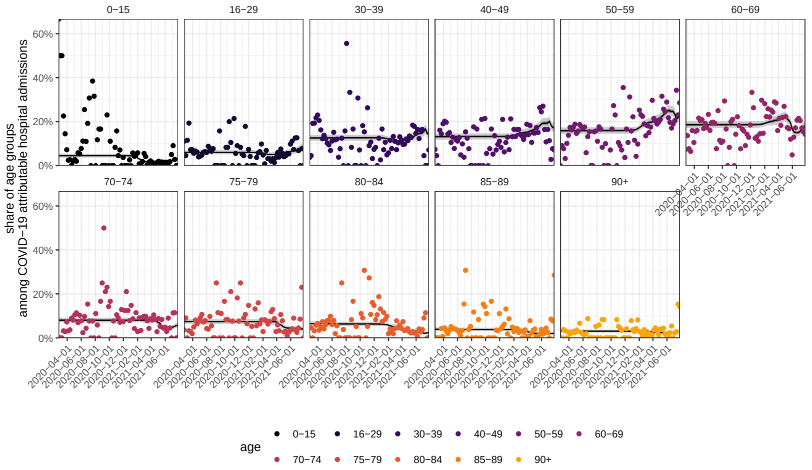


**N**

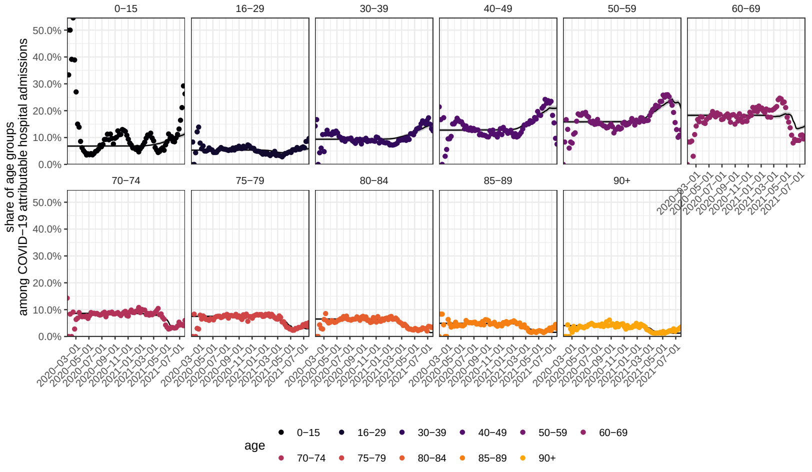


**Figure S13:** (continued) Model fits of the expected age composition of COVID-19 attributable hospital admissions. (L) For Salvador. (M) For São Luís. (N) For São Paulo city.

A

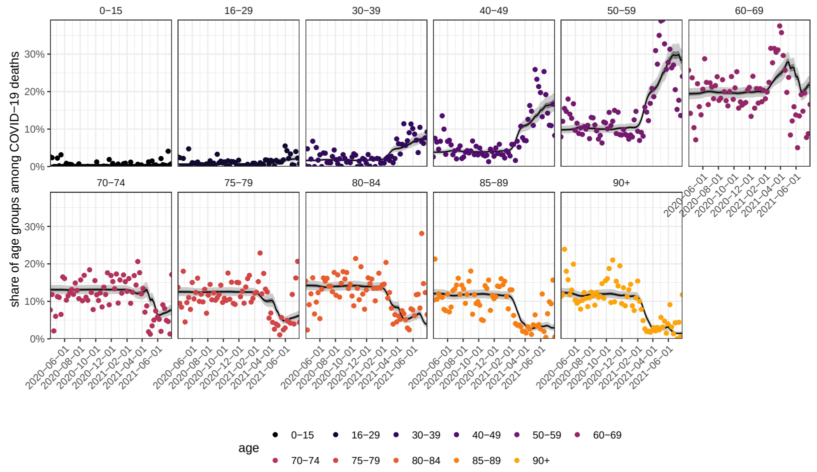


B

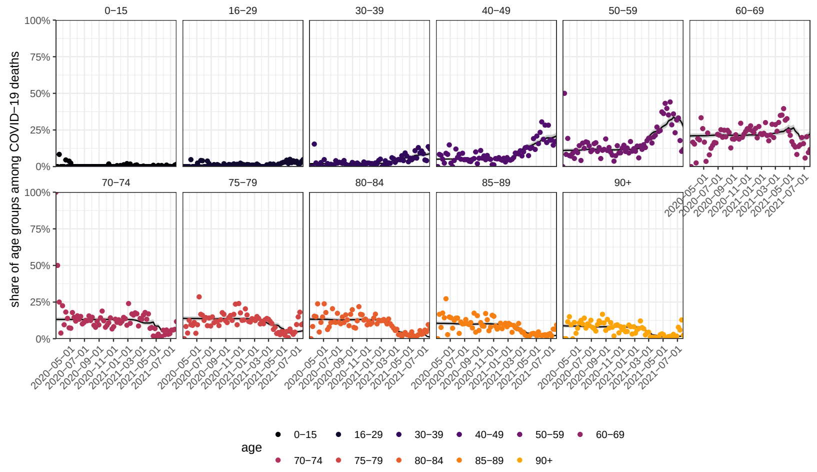


**Figure S14:** Model fits of the expected age composition of COVID-19 attributable deaths following hospital admission. Posterior median estimates of the expected age composition of deaths in each location, age band, and week, obtained with the Bayesian multi-strain fatality model, Supplementary Text Equation (S42), are shown on the y-axis as a black line along with 95% credible intervals. Time trends are shown by week of hospital admission (x-axis). The empirical proportions are shown as dots. (A) For Belo Horizonte. (B) For Curitiba.

**C**

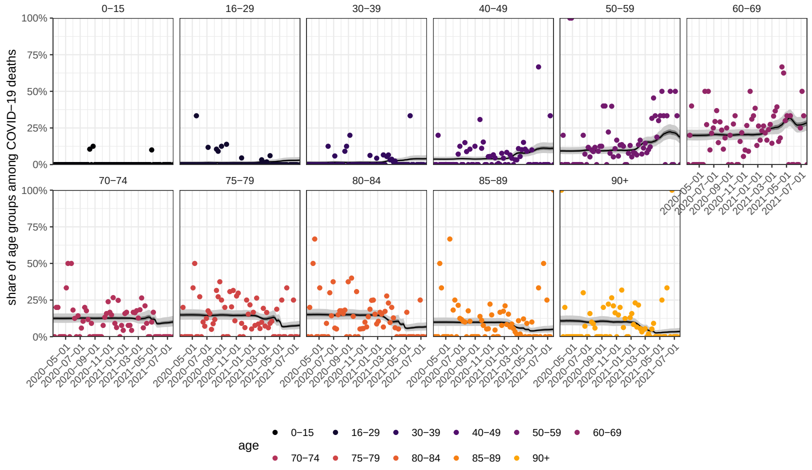


**D**

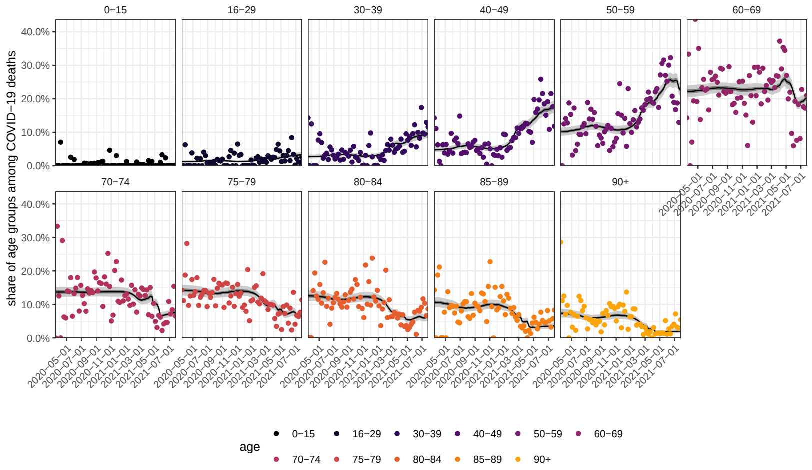


**E**

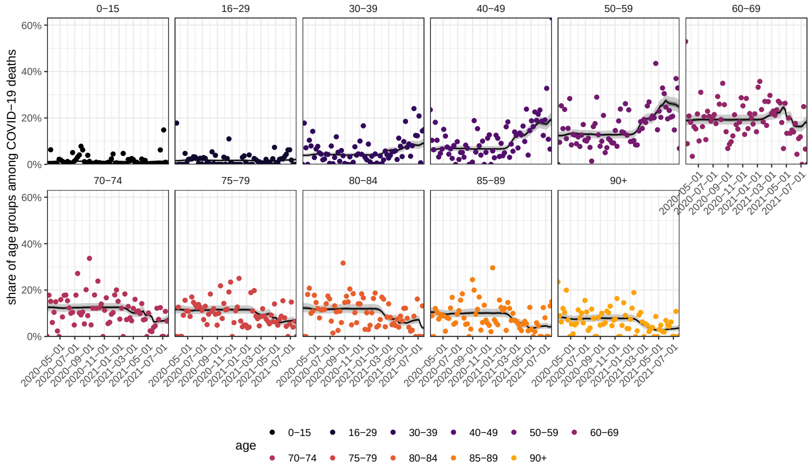


**Figure S14:** (continued) Model fits of the expected age composition of COVID-19 attributable deaths following hospital admission. (C) For Florianópolis. (D) For Goiânia. (E) For João Pessoa.

F

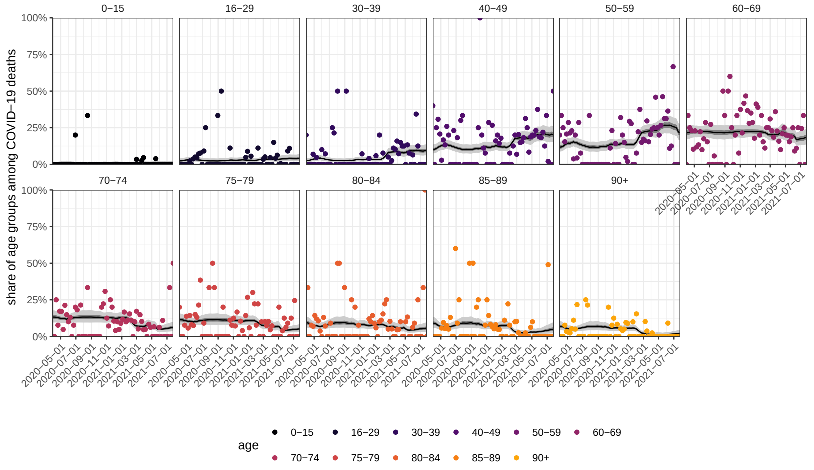


G

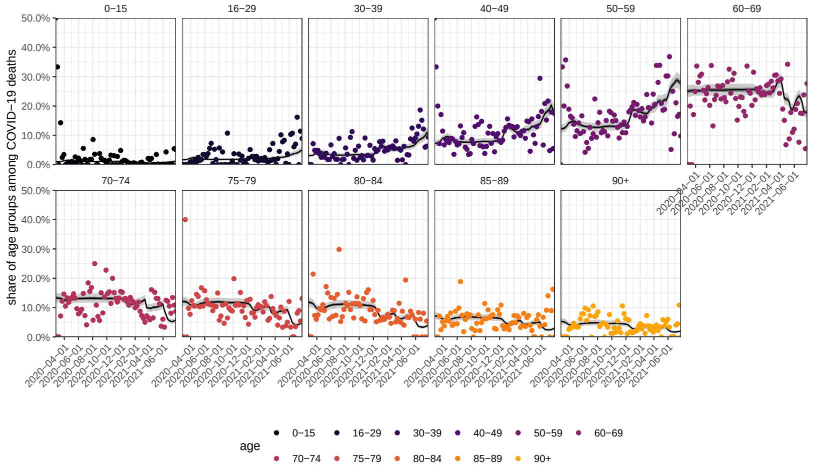


H

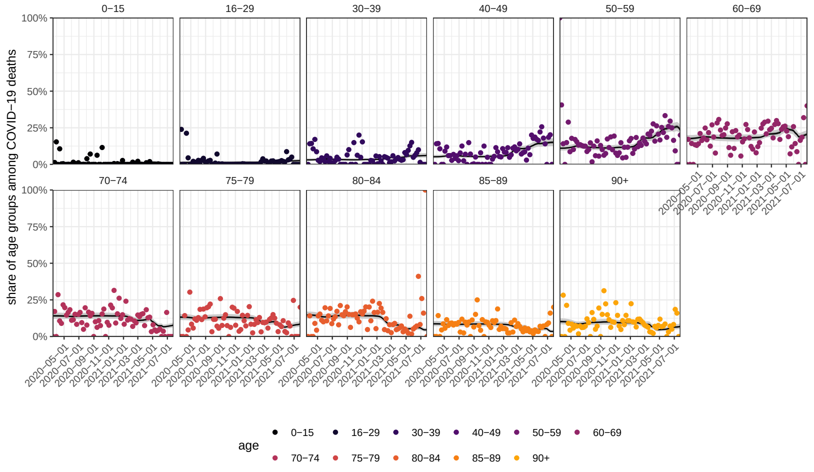


**Figure S14:** (continued) Model fits of the expected age composition of COVID-19 attributable deaths following hospital admission. (F) For Macapá. (G) For Manaus. (H) For Natal.

**I**

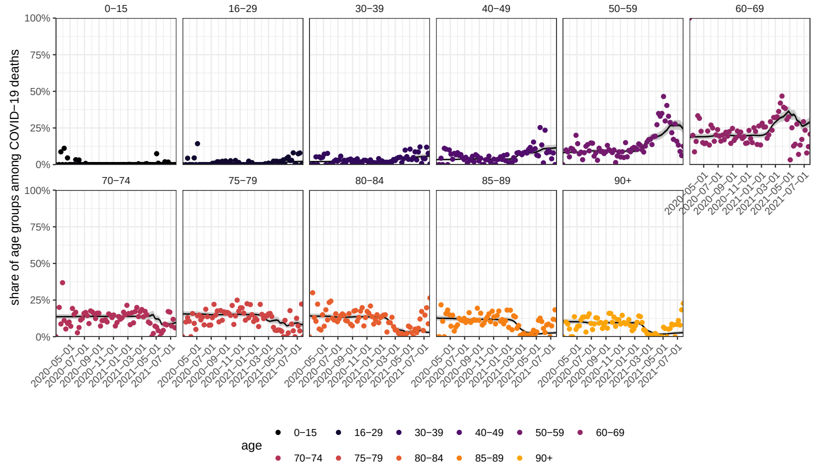


**J**

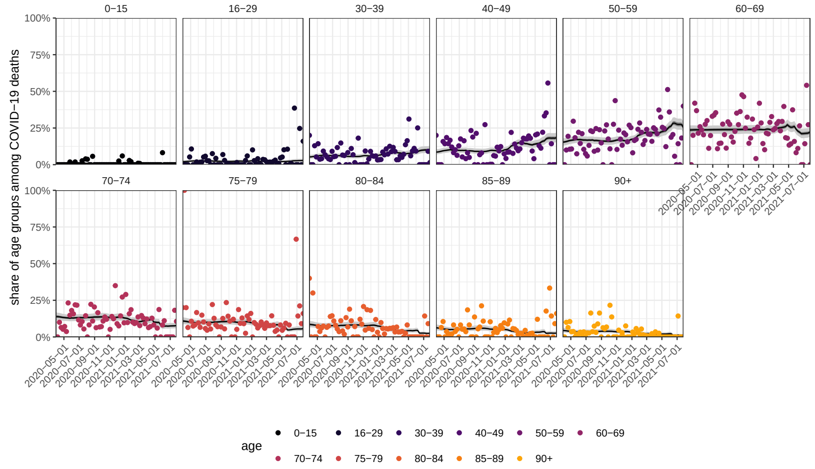


**K**

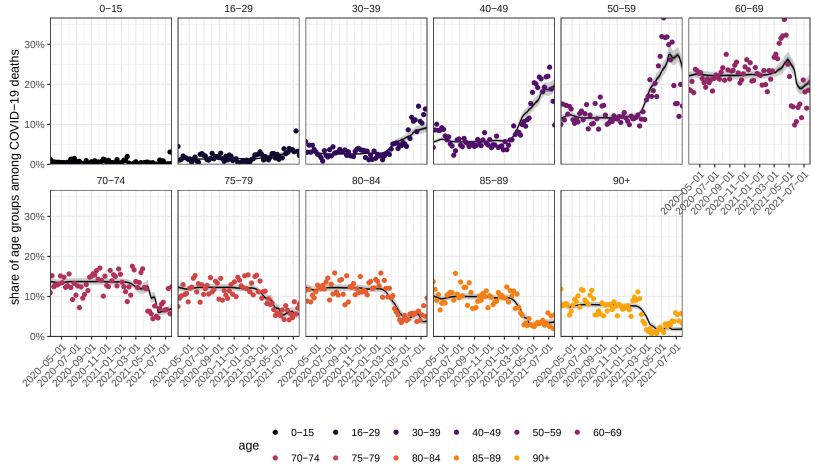


**Figure S14:** (continued) Model fits of the expected age composition of COVID-19 attributable deaths following hospital admission. (I) For Porto Alegre. (J) For Porto Velho. (K) For Rio De Janeiro.

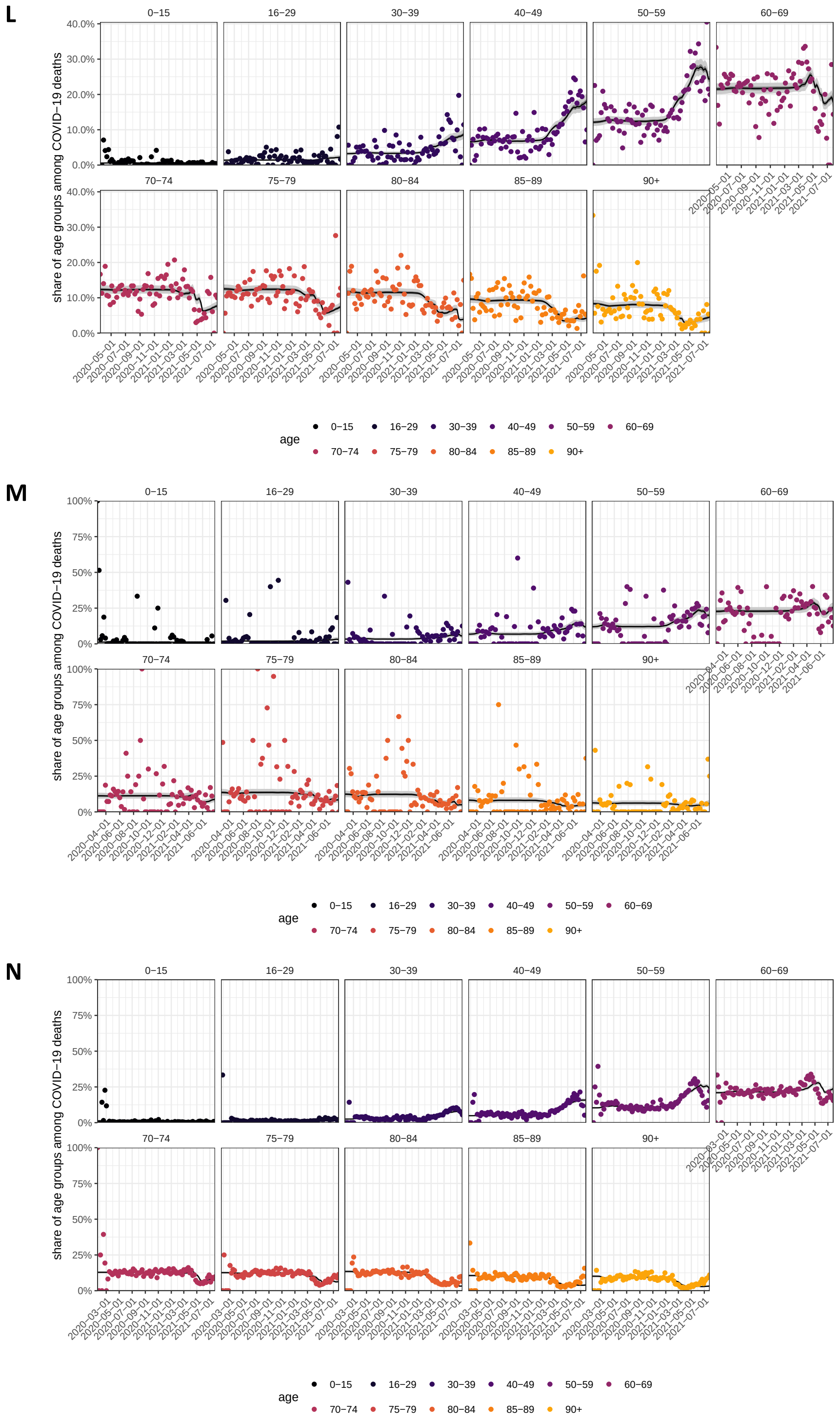


**Figure S14:** (continued) Model fits of the expected age composition of COVID-19 attributable deaths following hospital admission. (L) For Salvador. (M) For São Luís. (N) For São Paulo city.

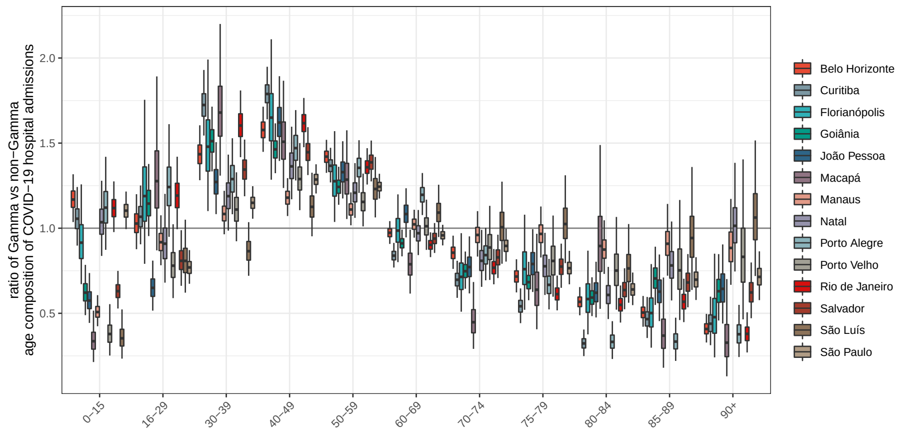


**Figure S15:** Estimated ratio in the share of age groups in Gamma versus non-Gamma residents' hospital admissions. Posterior median estimates are shown as black horizontal bar, 50% interquantile ranges as box and 95% credible intervals as whiskers. Estimates were derived independently for each location (colour) and control for changes in the population at risk of a fatal outcome over time, see Supplementary Text, page 51. Estimates for which credible interval width was larger than 3 were removed from the figure.

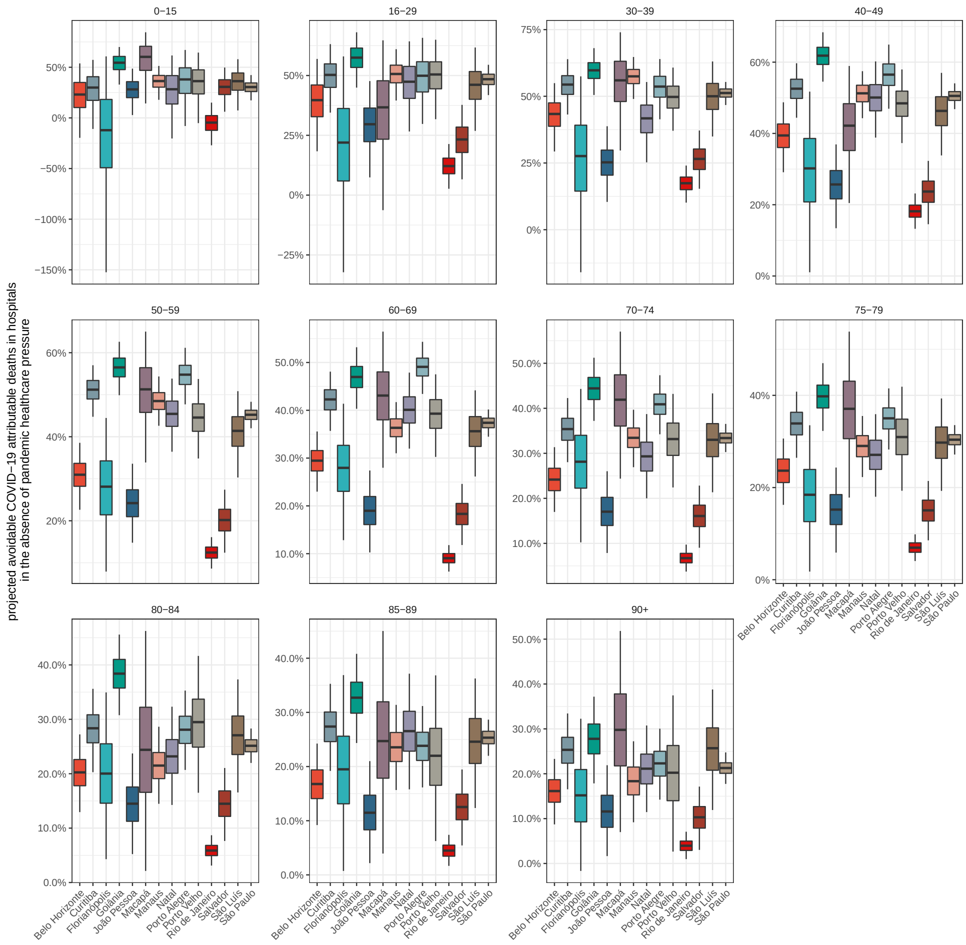


**Figure S16:** Projected proportion of avoidable COVID-19 attributable deaths in hospitals without pandemic resource limitations. The projections are based on counterfactual simulations assuming the lowest COVID-19 in-hospital fatality rates that were achieved prior to Gamma's detection in the corresponding location. Posterior median estimates of the proportion of avoidable COVID-19 attributable in-hospital deaths are shown as black horizontal bar, 50% interquantile ranges as box, and 95% credible intervals as whiskers.

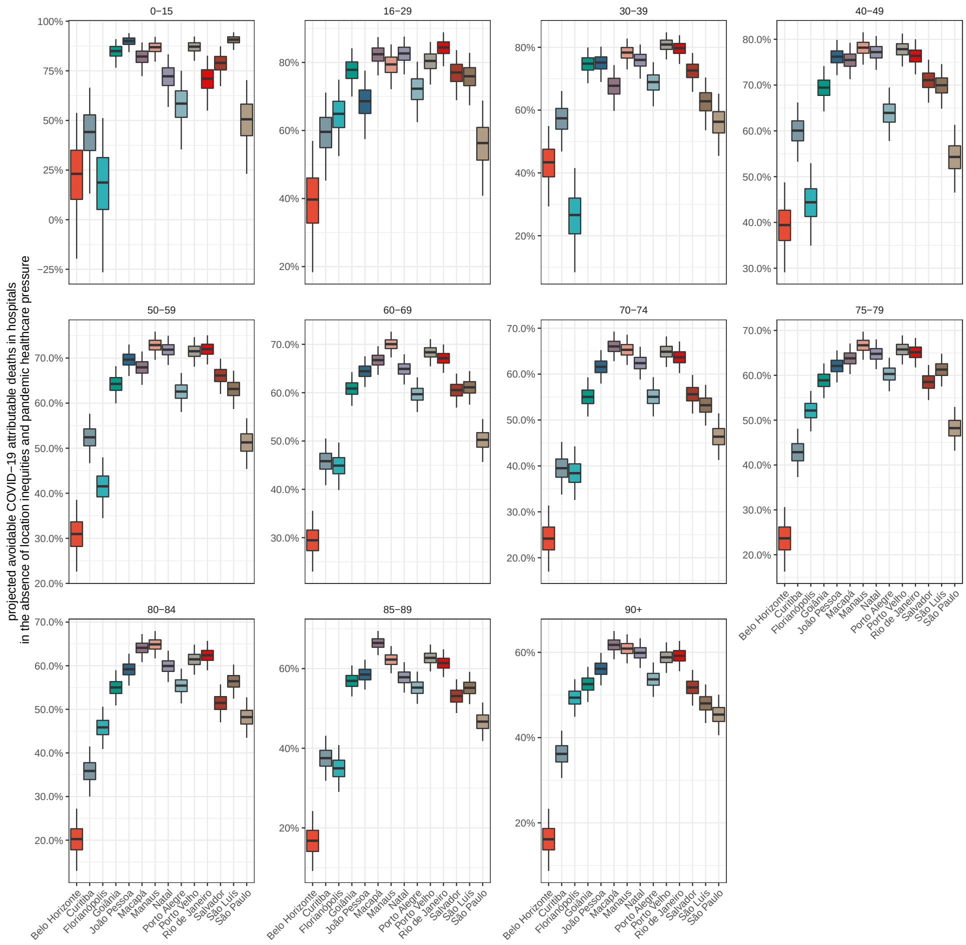


**Figure S17:** Projected proportion of avoidable COVID-19 attributable deaths in hospitals without location inequities and without pandemic resource limitations. The projections are based on counterfactual simulations assuming the lowest COVID-19 in-hospital fatality rates that were achieved prior to Gamma's detection across all locations. Posterior median estimates of the proportion of avoidable COVID-19 attributable in-hospital deaths are shown as black horizontal bar, 50% interquantile ranges as box, and 95% credible intervals as whiskers.
