## Supplementary Text for "Report 46: Factors driving extensive spatial and temporal fluctuations in COVID-19 fatality rates in Brazilian hospitals"

Affiliations

This PDF file includes:
Supplementary Text, with sections S1-S9
References

**S3 Controlling for hospitalised patients with unknown outcomes** **18**

**S4 Smoothed age-standardised COVID-19 in-hospital fatality rates** **20**

**S5 Healthcare pressure indices** **22**

**S6 Controlling for pandemic associated mortality and vaccine roll-out** **24**

### S1 Data sets

This section provides technical details on all data sets used in this study. Section S1.1 presents details on the sampling frame of the study. We focus our analysis on 14 state capitals, Belo Horizonte, Curitiba, Florianópolis, Goiânia, João Pessoa, Macapá, Manaus, Natal, Porto Alegre, Porto Velho, Rio de Janeiro, Salvador, São Luís and São Paulo, for which SARS-CoV-2 sequence data were publicly available to reconstruct Gamma's temporal expansion at city-level. Availability of these data are key to decompose the factors associated with fluctuations in in-hospital fatality rates, and so determined the locations that could be included in the study. Reflecting the geographical expansion of COVID-19 through Brazil over time, our observation periods varied across cities. Section S1.2 presents the SARS-CoV-2 sequence data used in this study. Section S1.3 describes the population sizes that we use in each state capital as population denominators. Sections S1.4-S1.7 describe data sets extracted from the SIVEP-Gripe data, which is produced by the Brazilian Ministry of Health [1, 2] and provides detailed, patient-level information on all individuals hospitalised with severe acute respiratory illness. We characterise COVID-19 attributable hospital admissions, deaths, and in-hospital fatality rates based on these data. Section S1.8 presents data from Brazil's National Register of Health Facilities (Cadastro Nacional de Estabelecimentos de Saúde (CNES)) [3] on reported healthcare resources in each location. These data underlie the healthcare pressure indices. Section S1.9 introduces all-cause death data from Brazil's Civil Registry [4], and Section S1.10 introduces individual-level data on administered vaccine doses, obtained from the Brazilian Ministry of Health website. These data were used in the Bayesian multi-strain fatality model to adjust population denominators to the estimated populations that remain at risk of fatal COVID-19 outcomes in each location. All data are freely available at the github repository https://github.com/CADDE-CENTRE/covid19_brazil_hfr.

#### S1.1 Choice of study locations and observation period

Individual patient records in Brazil's SIVEP-Gripe data do not contain linked SARS-CoV-2 sequence data. To characterise Gamma's effect on in-hospital fatality rates, we estimated Gamma's temporal expansion at population level in geographically well-defined locations from publicly available SARS-CoV-2 sequence data. We searched GISAID (https://www.gisaid.org) on June 14, 2021, for sequence data associated with SARS-CoV-2 Gamma virus genome sequences in the 27 federal units (26 Brazilian states and the Federal District). Gamma genome data was available for 16 federal units, Amapá, Amazonas, Bahia, Goiás, Maranhão, Mato Grosso do Sul, Minas Gerais, Paraíba, Paraná, Rio de Janeiro, Rio Grande do Norte, Rio Grande do Sul, Rondônia, Santa Catarina, São Paulo, and Tocantins. We focused our analysis on the state capitals in these federal units because key variables such as population size, vaccination coverage and healthcare indicators were more directly available at the level of state capitals. We assumed that the frequency of Gamma in state capitals is similar to the frequency of Gamma across that state, measured by data available in GISAID and metadata available from Rede Genômica FioCruz that provides more representative sampling [5]. Two state capitals, Palmas and Campo Grande, were excluded from further analysis due to limited, weekly age-specific COVID-19 hospital admission and death counts among residents reported in SIVEP-Gripe. The cities in our sampling frame were thus Belo Horizonte, Curitiba, Florianópolis, Goiânia, João Pessoa, Macapá, Manaus, Natal, Porto Alegre, Porto Velho, Rio de Janeiro, Salvador, São Luís and São Paulo.

Reflecting the geographical expansion of COVID-19 through Brazil over time, our observation periods varied across

| State capital | State | Observation period | | SARS-CoV-2 Gamma variant | |
|---|---|---|---|---|---|
| | | Start | End | Estimated emergence (posterior median) | First detection (observed) |
| Belo Horizonte | Minas Gerais | 06/04/2020 | 26/07/2021 | 15/02/2021 | 15/12/2020 |
| Curitiba | Paraná | 02/03/2020 | 26/07/2021 | 26/01/2021 | 22/10/2020 |
| Florianópolis | Santa Catarina | 09/03/2020 | 26/07/2021 | 08/01/2021 | 11/12/2020 |
| Goiânia | Goiás | 16/03/2020 | 26/07/2021 | 19/01/2021 | 28/11/2020 |
| João Pessoa | Paraíba | 09/03/2020 | 26/07/2021 | 21/01/2021 | 20/12/2020 |
| Macapá | Amapá | 30/03/2020 | 26/07/2021 | 16/01/2021 | 14/12/2020 |
| Manaus | Amazonas | 24/02/2020 | 26/07/2021 | 04/12/2020 | 04/12/2020 |
| Natal | Rio Grande do Norte | 16/03/2020 | 26/07/2021 | 26/01/2021 | 06/12/2020 |
| Porto Alegre | Rio Grande do Sul | 02/03/2020 | 26/07/2021 | 10/01/2021 | 21/11/2020 |
| Porto Velho | Rondônia | 30/03/2020 | 26/07/2021 | 18/01/2021 | 25/12/2020 |
| Rio de Janeiro | Rio de Janeiro | 16/03/2020 | 26/07/2021 | 19/01/2021 | 29/11/2020 |
| Salvador | Bahia | 16/03/2020 | 26/07/2021 | 28/12/2020 | 02/12/2020 |
| São Luís | Maranhão | 24/02/2020 | 26/07/2021 | 19/01/2021 | 04/12/2020 |
| São Paulo city | São Paulo | 20/01/2020 | 26/07/2021 | 01/02/2021 | 01/11/2020 |

**Table S4:** State capitals included and observation periods, with estimated dates of Gamma's emergence and first detection based on GISAID data.

cities. The start date was defined as the Monday after the date on which at least 2.5 patients with suspected or confirmed COVID-19 per 100,000 population were hospitalized in each location. To minimise the impact of reporting delays to SIVEP-Gripe on our inferences, the end date was set to 26 July 2021. Figure 1 in the main text and Table S4 report the 14 cities investigated in this study, and the corresponding observation periods.

#### S1.2 GISAID metadata to characterise Gamma's temporal expansion

Collection dates of all Brazilian Gamma sequences available were obtained from GISAID on June 28, 2021, with collection dates between November 1, 2020 and March 31, 2021. Collection dates could be associated with 8,609 sequences, among which 94 entries were removed due to incomplete collection dates, and 14 were removed because they were duplicates. Out of the remaining 8,501 samples, we retained the 7,221 samples collected in the 14 Brazilian states, which are shown in Figure S18. Most of the sequence data were from São Paulo (1104 viral genomes), few from Rondônia (9), and on average states reported 158 genomes. Acknowledgment tables with GISAID IDs are available at acknowledgments_GISAID_Tables in the github repository.

#### S1.3 Population denominators

National population estimates by sex and age were retrieved from the 2020 National Household Sample Survey COVID-19, Pesquisa Nacional por Amostra de Domicílios COVID-19 (PNAD COVID-19) [6] by the Brazilian Institute of Geography and Statistics (IBGE). Between May and September 2020, over 1.888.560 interviews were conducted by IBGE in all state capitals and the Federal District of Brazil, from which population sizes were estimated for men and women by $1$-year age bands. The population size projections were adjusted so that at most 99% of residents

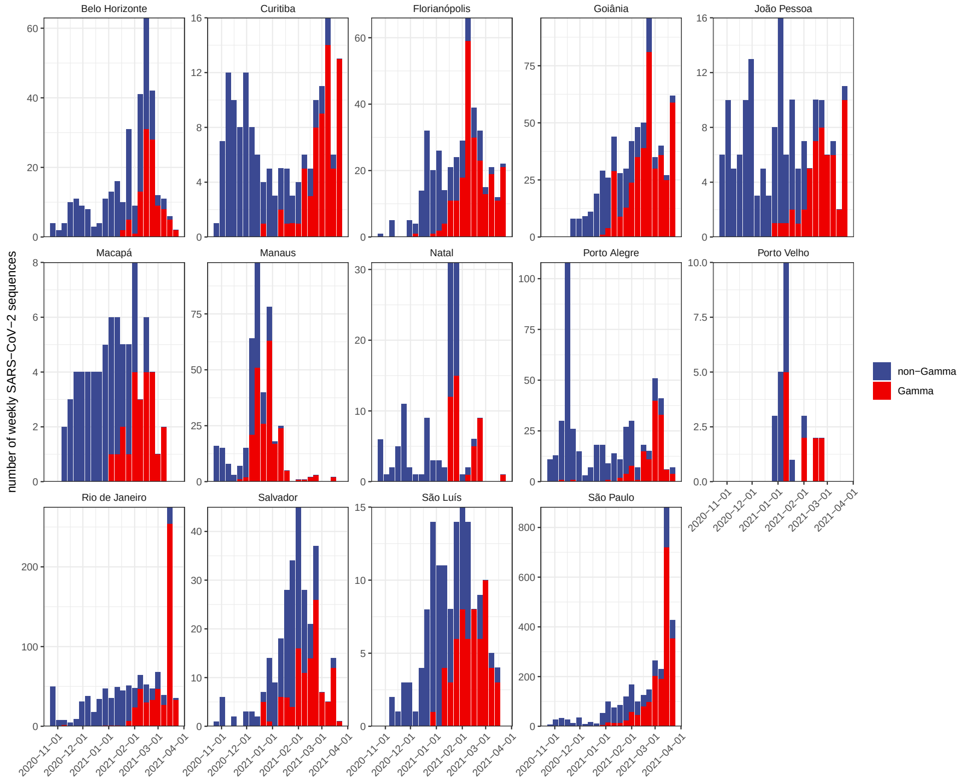

**Figure S18:** SARS-CoV-2 sequence data obtained from GISAID in the 14 states in which the 14 state capitals are located.

received at least one vaccine dose during the observation period as further described in Section S6. Figure S19 illustrates the PNDAc 2020 population size projections along with these adjustments.

The population sizes were then stratified into the following age bands

$$\mathcal{A} = \Big\{0-15, 16-29, 30-39, 40-49, 50-59, 60-69, \\ 70-74, 75-79, 80-84, 85-89, 90+\Big\}, \tag{S1}$$

and we denote by $n_{l,a}$ the (adjusted) projected 2020 population sizes in location $l$ and age band $a$. Children and younger adults were grouped by 15 year age bands, middle aged adults were grouped by 10 year age bands, and older adults were grouped by 5 year age bands to reflect exponentially increasing infection severity of SARS-CoV-2 [7]. The resulting population size estimates are available at inst/data/PNADc_vaxadj_population_210802.csv in the github repository.

#### S1.4 Hospital admissions with reported severe acute respiratory illness

The first data set that we extracted from SIVEP-Gripe describes for each location during our observation period the incidence of severe acute respiratory illness hospitalisations due to any cause, not restricted to COVID-19. This data set is used in Section S5 to calculate healthcare pressure indices, and with this aim in mind captures hospitalised patients in each location regardless of residency or vaccination status.

All patients presenting to Brazilian hospitals with severe acute respiratory illness (SARI) must be notified onto the SIVEP-Gripe platform, which records their clinical details as well as personal information such as city of residence, city of hospitalisation, and age [1, 2]. Following reporting directives issued since the start of the COVID-19 pandemic [8, 9], the SIVEP-Gripe data also capture COVID-19 attributable deaths that occurred out of hospital, and that were known to data reporters. To exclude out-of-hospital deaths reported to SIVEP-gripe, we retained reported cases who as of 19 September 2021 had a non-empty hospital admission date, or did not die on the same date. All SARI cases were stratified into the age strata described in equation (S1). Table S5 reports the total number of hospital admissions extracted from the SIVEP-Gripe data set.

#### S1.5 COVID-19 attributable hospital admissions in residents and non-residents without evidence of vaccination

The second data set describes COVID-19 attributable hospital admissions and associated metadata during the observation period among residents and non-residents in each location. This data set is (only) used to account for underreporting of deaths in hospitalised and unvaccinated patients with unknown clinical outcomes (Section S3).

During the course of hospital admission, the attending team, in conjunction with local health authorities, establish a final classification of SARI diagnosed patients, integrating diagnostic tests, epidemiological links to other cases and clinical information. The data and classifications are retrospectively updated in the latest data release. We defined as COVID-19 attributable hospital admissions all patients of class 4, 5 or missing classification, comprising PCR-confirmed COVID-19 infections, clinically diagnosed COVID-19 infections, and patients with severe acute respiratory infection and no evidence of infection with other respiratory pathogens. Table S5 shows the proportion of hospitalised patients in our observation periods of each location by SIVEP-Gripe classification.

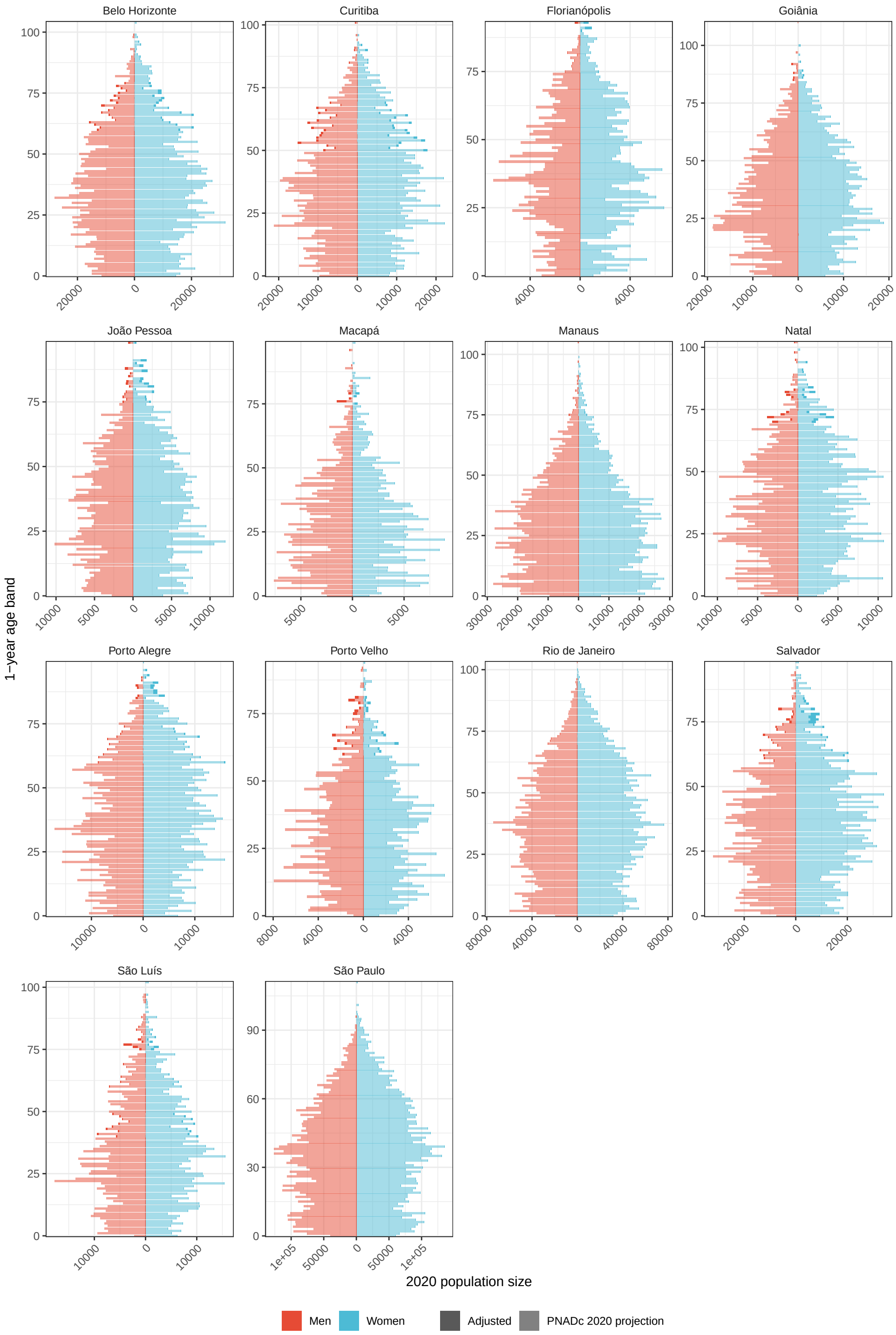

**Figure S19:** PNADc 2020 population size projections by sex and 1-year age bands across state capitals and the Federal District. For our purposes, size projections were adjusted so that at most 99% of residents received at least one vaccine dose during the observation period.

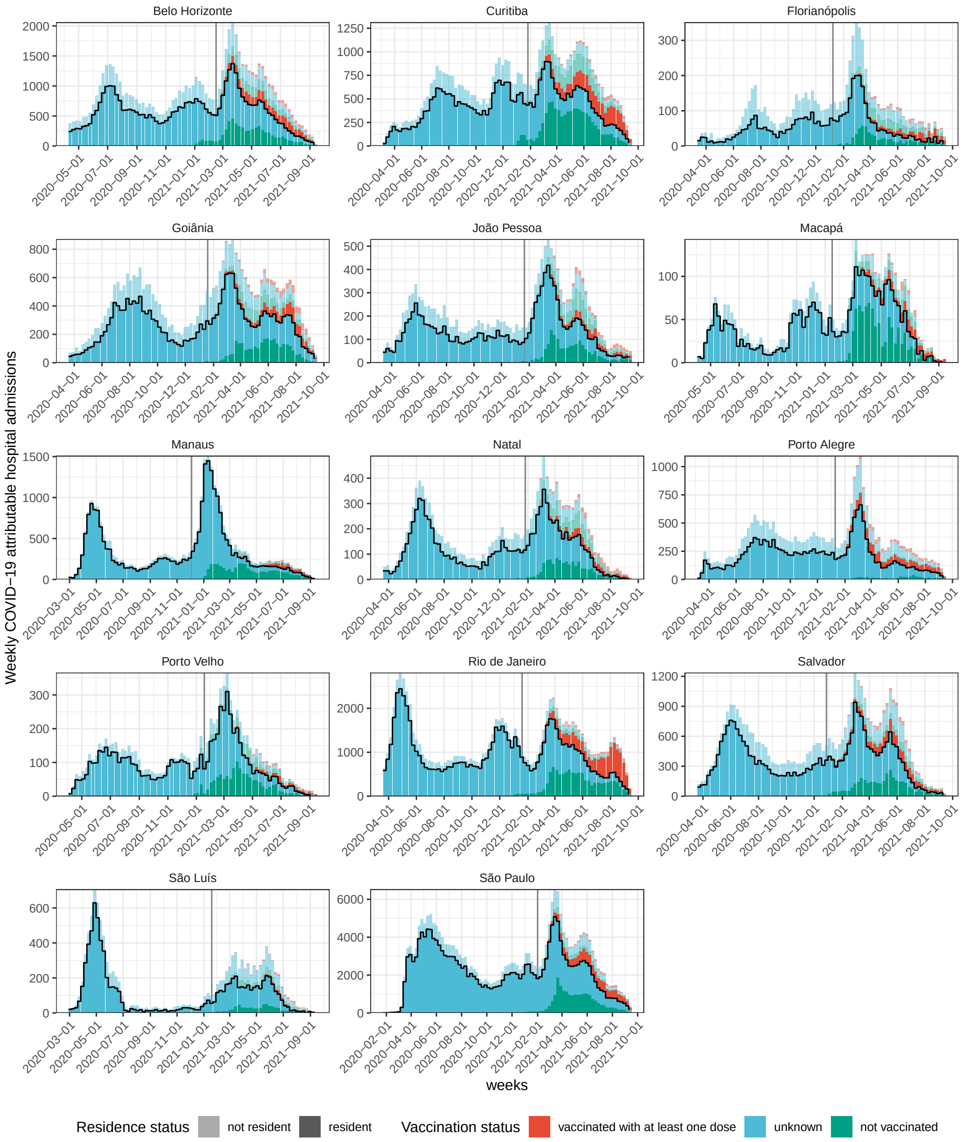

**Figure S20:** Weekly, COVID-19 attributable hospital admissions stratified by residence status (lighter shade for non-residents and darker shade for residents) and vaccination status (colours) in 14 cities in Brazil. Data are from the SIVEP-Gripe platform of the Brazilian Ministry of Health and as of 20 September 2021. The black line indicates the hospitalised patients selected in this study.

| | SIVEP-Gripe classification | | | | |
|---|---|---|---|---|---|
| City | Hospital admissions | 1-3 Other Cause | 4 Suspected COVID-19 | 5 Confirmed COVID-19 | Unknown |
| Belo Horizonte | 72221 | 0.79% | 42.56% | 51.77% | 4.87% |
| Curitiba | 54894 | 2.27% | 31.60% | 64.07% | 2.06% |
| Florianópolis | 8854 | 5.20% | 33.98% | 59.06% | 1.76% |
| Goiânia | 36083 | 1.29% | 20.95% | 72.84% | 4.92% |
| João Pessoa | 18447 | 0.49% | 26.57% | 63.26% | 9.68% |
| Macapá | 4436 | 0.47% | 4.26% | 94.32% | 0.95% |
| Manaus | 31404 | 1.37% | 20.82% | 76.77% | 1.03% |
| Natal | 15749 | 1.16% | 23.84% | 66.94% | 8.06% |
| Porto Alegre | 28793 | 1.67% | 23.88% | 73.21% | 1.24% |
| Porto Velho | 9889 | 1.22% | 14.71% | 78.11% | 5.96% |
| Rio de Janeiro | 105645 | 0.70% | 17.11% | 76.29% | 5.90% |
| Salvador | 43072 | 1.24% | 26.40% | 71.54% | 0.82% |
| São Luís | 15260 | 3.40% | 29.31% | 54.95% | 12.34% |
| São Paulo | 257673 | 1.39% | 27.98% | 65.49% | 5.14% |

**Table S5:** Reported SARI cases, hospital admissions, and SIVEP-Gripe classification.

Among the COVID-19 attributable hospital admissions, we excluded patients with evidence in the SIVEP-Gripe data of having had at least one vaccine dose administered before hospitalisation. For brevity, we refer to these patients as unvaccinated in the Supplementary Text. Here, we denote the COVID-19 attributable hospital admissions in unvaccinated patients in location $l$ and age band $a$ on day $t$ by $h_{l,a,t}$. Similarly, we denote the COVID-19 attributable hospital admissions in location $l$ and age band $a$ in week $w$ by

$$h_{l,a,w} = \sum_{t \in w} h_{l,a,t}. \tag{S2}$$

Figure S20 illustrates the weekly number of COVID-19 attributable hospital admissions by residency and vaccination status.

#### S1.6 COVID-19 attributable hospital admissions in residents without evidence of vaccination

The third data describes weekly, COVID-19 attributable hospital admissions and associated metadata during the observation period in unvaccinated residents. This is our primary data, and used to estimate COVID-19 in-hospital fatality rates. We focus on residents because our inferences are linked to further data on the city-level populations, i. e. individuals that are resident in each location.

Of the COVID-19 attributable hospital admissions in unvaccinated patients (Section S1.5), we retained those patients who were resident in each location. We denote the COVID-19 attributable, unvaccinated hospitalised residents in location $l$ and age band $a$ day $t$ by $h^{\text{res}}_{l,a,t}$. Similarly, we denote the COVID-19 attributable, unvaccinated hospitalised residents in location $l$ and age band $a$ in week $w$ by

$$h^{\text{res}}_{l,a,w} = \sum_{t \in w} h^{\text{res}}_{l,a,t}. \tag{S3}$$

Figure S20 shows that the contribution of non-residents to hospitalised patients differed substantially across locations.

#### S1.7 COVID-19 attributable deaths among residents

The SIVEP-Gripe platform reports deaths that occurred in-hospitals and, following reporting directives issued since the start of the COVID-19 pandemic [8, 9], deaths that occurred out of hospital, and that were known to data reporters. These data are used in Section S6 to adjust population denominators over time. We retained all reported COVID-19 attributable deaths in residents in location $l$ and age band $a$ that occurred in week $w$, and denote them by $d^{\text{res-SIVEP}}_{l,a,w}$.

#### S1.8 Resources in healthcare facilities

To characterise time trends in healthcare pressure in each location, we obtained monthly data on healthcare resources reported by healthcare facilities to the National Register of Health Facilities (Cadastro Nacional de Estabelecimentos de Saúde - CNES) [3]. Data on healthcare resources are mandatory to report by both public and private healthcare facilities. This section provides technical details on our definitions of healthcare resources.

Overall, we aggregated for each location monthly data on personnel (nurses, nurse assistants, physiotherapists, physicians and critical care specilists i. e. intensive care physicians) and equipment (critical care beds, critical care beds with ventilators, ICU beds, ventilators). The data are available in file inst/data/IPEA_ICUbeds_physicians_210928.csv in the github repository, and are as of September 15, 2021. Figure S5 summarises the healthcare resources per 100,000 population based on the 2020 population size projections described in Section S1.3. We use the data on healthcare resources to define the healthcare pressure indices shown in Figure 3.

To count resources for treatment of severe COVID-19, we considered several types of ICU beds reported to CNES. ICU type I beds (code 74) refer to an older standard that is phased out since 2017, and were not counted. Type II ICU beds (code 75) represent the minimum requirement for severe cases of COVID-19 requiring ventilation. Type III ICU beds (code 76) are, by regulation, reserved to ICU patients with multiple acute failures of vital organs, or to patients at risk of developing them, with an immediate threat to life [10]. Since March 2020, new ICU beds were created to try to minimise the immediate risk of healthcare system collapse (code 51). These ICU beds were intended exclusively for treatment of COVID-19, and are designated COVID-19 type II ICU beds. At least one microprocessor-controlled lung ventilator must be available for every two ICU type II, ICU type III, and COVID type II beds. Considering that both a ventilator and an ICU bed are necessary for adequate treatment of severe COVID-19, we only counted adult ICU beds with a ventilator per month and location, defined as the minimum number of available ventilators and the number of adult ICU type II, ICU type III, COVID-19 type II beds in the same healthcare facility that are reported to CNES, which we then aggregated across healthcare facilities in the same location.

We further considered critical care beds, with and without controlling for ventilators, to more broadly quantify beds used for the treatment of critical or severe cases of COVID-19 according to national guidelines ([11], p.19). Critical care beds summed, per month and location, the number of adult ICU type II, ICU type III beds, COVID-19 type II and intermediate care beds (code 95) reported to CNES. We considered both this sum and the minimum between the sum and the number of available ventilators in the same healthcare facility, which we then aggregated across healthcare facilities in the same location.

To guard against potential reporting differences or bias in reported bed types, we also considered monthly counts of available ventilators as resource. The number of reported ventilators (respirador or ventilador, code 64) does not include ventilators already held for ICU or critical care beds. Thus, for each healthcare facility, we counted the number of lung ventilators reported to CNES, and added one ventilator for every two reported ICU type II, type III, or COVID-19 type II beds, and added one ventilator for every three reported intermediate beds. We then again aggregated across healthcare facilities in the same location.

Physicians summed, per month and location, the number of medical professionals as identified by name and their professional health card number (Cartão Nacional de Saúde do professional – CNS_prof) within the occupational job families for medical doctors (families CBO 2231, 2251, 2252,2253) according to the 2002 Brazilian Classification of Occupations (Classificação Brasileira de Ocupações – CBO).

Critical care specialists (or intensive care specialists) summed, per month and location, the number of intensive care specialists reported within the occupational job family CBO 225150.

Nurses summed, per month and location, the number of nurses reported within the occupational job family CBO 2235. Nurse assistants summed, per month and location, the number of individuals reported within the occupational job family CBO 3222. In Brazil, the term nurses is used for professionals holding a university degree in Nursing, which is a 5-year training program, while the term technical nurse or nurse assistant is used for individuals that hold a nursing assistant certificate, obtained after a 18-24 months high-school level training program. Physiotherapists summed, per month and location, the number of physiotherapists reported within the occupational job family CBO 2236. In Brazil, physiotherapists are professionals holding a university degree in Physiotherapy, which is a 5-year training program.

#### S1.9 Excess deaths

To specify in the Bayesian multi-strain fatality rate model the population at risk of severe COVID-19, we obtained daily, age-specific, all-cause death records from the Brazilian Civil Registry [4], as well as daily, age-specific COVID-19 specific death records from the same data source. Data on the exact date, city and state where death occurred, age and sex of the deceased, and whether death occurred in or out of hospital were obtained from [12]. In the data, the cause of death is classified as COVID-19 related if COVID-19 or similar terms are mentioned anywhere in the death certificate.

The all-cause deaths reported in the Civil Registry are stratified by the age bands

$$\begin{aligned}\mathcal{B} = \big\{&0-9, 10-19, 20-29, 30-39, 40-49, 50-59, 60-69, 70-79,\\ &80-89, 90-99, 100+\big\}.\end{aligned} \tag{S4}$$

To match these age bands with our age bands (S1), we weighted the reported deaths in larger age brackets than ours by the population proportion in our age bracket. For example, we calculated

$$v^{\text{all-cause}}_{l,10-15,t} = \frac{n_{l,10-15}}{n_{l,10-19}}\, v^{\text{all-cause}}_{l,10-19,t}, \tag{S5}$$

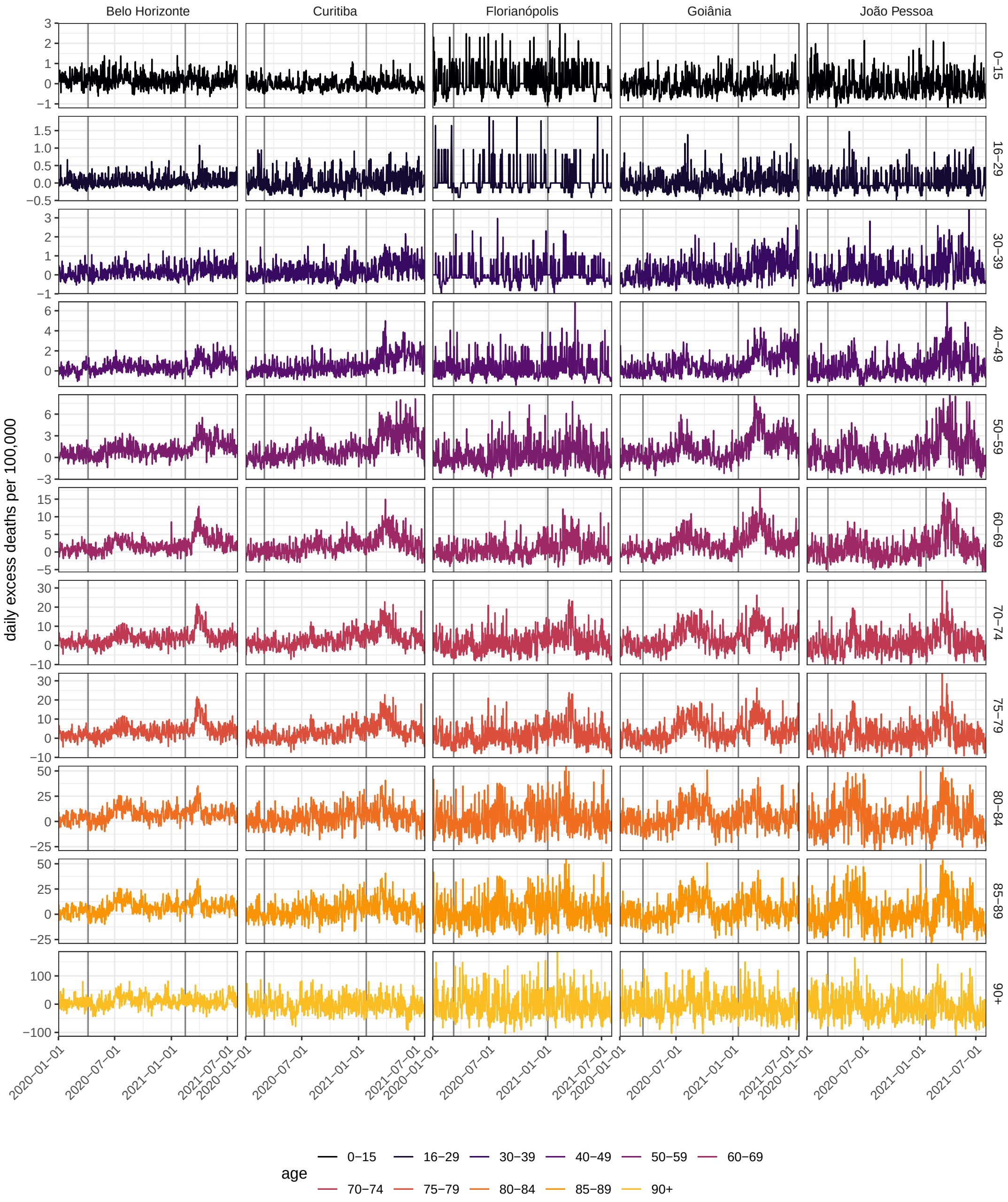

**Figure S21:** Daily age-specific excess deaths per 100,000 individuals in Belo Horizonte, Curitiba, Florianópolis, Goiânia, João Pessoa. Daily excess deaths were calculated from all-cause deaths reported by the Brazilian Civil Registry. The start date of the observation period and the date of Gamma's detection in each city are indicated as vertical dotted grey lines.

where $n_{l,10-15}$ and $n_{l,10-15}$ are the 2020 population projections for location $l$ in the corresponding age bands described in Section S1.3, and $v^{\text{all-cause}}_{l,10-19,t}$ are the all-cause deaths of age $10$-$19$ reported in the Civil Registry in location $l$. Records in smaller age brackets were summed. The COVID-19 related deaths reported in the Civil Registry were weighted and summed analogously to harmonise them into the age bands used in this study.

We then derived the excess deaths in location $l$, age band $a$ on day $t$ since January 1, 2020 by subtracting from the all-cause deaths $v^{\text{all-cause}}_{l,10-15,t}$ the smoothed, 7-day rolling mean of the all-cause deaths on the same day in 2019. Since 2020 was a leap year, we duplicated the all-cause death counts of February 28, 2019 for an artificial February 29, 2019. Specifically, we calculated

$$v^{\text{excess}}_{l,a,t} = v^{\text{all-cause}}_{l,a,t} - \frac{1}{7}\sum_{s=t-3}^{t+3} v^{\text{all-cause}}_{l,a,s-366}. \tag{S6}$$

Figure S21 illustrates the derived excess deaths for five cities, indicating that the excess deaths were in most state capitals higher after than before Gamma's detection (vertical grey line).

The daily excess deaths and COVID-19 related deaths reported by the Brazilian Civil Registry were then aggregated by week,

$$v^{\text{excess}}_{l,a,w} = \sum_{t\in w} v^{\text{excess}}_{l,a,t} \tag{S7}$$

$$v^{\text{covid}}_{l,a,w} = \sum_{t\in w} v^{\text{covid}}_{l,a,t}. \tag{S8}$$

#### S1.10 Administered vaccine doses among residents

To specify in the Bayesian multi-strain fatality rate model the population at risk of severe COVID-19, we further obtained individual-level data on administered vaccine doses from the Brazilian Ministry of Health database [13] on 05 August 21. This section provides technical details on how the data were pre-processed to estimate vaccine coverage in city populations. Our preprocessing steps are in file 'inst/utils/preprocess_vaccination_data_210804.R' and the preprocessed data are in file inst/data/saudegovbr_vaccinations_210805.rds in the github repository.

For all records, entries with an empty dose field were discarded. In one instance, the same vaccine type was reported with multiple names, and for this reason both "Covid-19-AstraZeneca" and "Vacina Covid-19 - Covishield" dose entries were classified as "Covishield". Considering records of each patient, we assumed that individual patient identifiers are unique. In the case the second dose was supposedly administered before the first dose, we decided to swap the dates of administration for the first and second dose. If the record for the first dose did not exist, we classified the second dose as the first. Similarly, if the date of administration of both doses coincided, the record for the second dose was ignored. For a small proportion of individuals, more than 2 administrations of the same dose were reported. In the case this issue concerned the first dose, only the first administration record was retained in chronological order. If the issue concerned the second dose, only the last administration record was retained. We found multiple administration records of the same dose on the same day. This issue was due to duplicate patient entries with non-unique date of birth or sex classification. In this case, entries were resolved by considering the most common value across multiple patient observations. When individuals were supposedly administered two different vaccines as first and second dose, we changed the vaccine type of the first dose to make it consistent with the second dose vaccine type.

To calculate vaccine coverage in residents in each location, the individual-level vaccine data were aggregated by location of residence into the number of individuals $v^{k,1}_{l,a,w}$ in location $l$ and age band $a$ that received a first dose of vaccine $k$ by week $w$, and similarly into the number of individuals $v^{k,2}_{l,a,w}$ in location $l$ and age band $a$ that received two doses of vaccine $k$ as of week $w$. We used the age of each patient on the first of January 2021 to stratify patients into our age bands. Our calculations to determine vaccine coverage are given in Section S6.

### S2 Sequence-based analyses to date Gamma's emergence and track Gamma's expansion

#### S2.1 Data selection for dating Gamma's emergence

To date the emergence of the SARS-CoV-2 Gamma variant in each location, only high coverage SARS-CoV-2 Gamma genome sequences with complete date of collection available on GISAID [14] by 14 June 2021 with dates of collection until 31 March 2021 were included. The metadata associated with the sequence data used for phylogenetic analysis thus corresponds to a subset of the metadata used to investigate replacement dynamics in each city, and a total of 2,212 were selected for the 14 states under study (Minas Gerais, Paraná, Santa Catarina, Goiás, Paraíba, Amapá, Amazonas, Rio Grande do Norte, Rio Grande do Sul, Rondônia, Rio de Janeiro, Bahia, Maranhão, and São Paulo). This data set included 5 sequences that had been recovered from the International Guarulhos Airport in São Paulo.

#### S2.2 Maximum likelihood phylogenetic tree estimation and data quality exploration

To confirm lineage classifications, all sequences were subjected to a Pango lineage classification version 3.0.6 (Pangolearn v.1.2.12 and scorpio lineage), and a few sequences were excluded from further analysis. Then, the reference strain WH04 (GISAID EPI_ISL_406801) was appended to the sequence dataset before multiple sequence alignment using MAFFT v7 [15]. After removing untranscribed terminal regions, the resulting multiple sequence alignment had a length of 29,409 nucleotides. Maximum likelihood phylogenetic trees were estimated using IQTree v2 [16] under the Jukes Cantor (JC69) substitution model [17]. We next used TempEst v.1.5.3 [18] to regress root-to-tip distances against sampling dates and identify data quality and data annotation problems prior to further phylogenetic analysis. Specifically, we discarded virus genomes characterized by a genetic distance to WH04 of more than 4 standard deviations from the epi-week mean genetic distance to WH04 [19]. A total of 10 sequences were excluded from subsequent phylogenetic analysis, as shown in Figure S22. The GISAID identifiers of the excluded sequences were EPI_ISL_1821206, EPI_ISL_2249444, EPI_ISL_1821208 (earliest available sequence from São Paulo, dated 2020-11-03), EPI_ISL_1821217, EPI_ISL_2249440 (earliest available sequence from Rio de Janeiro, dated 2020-11-18), EPI_ISL_2249443, EPI_ISL_2241496 (earliest available sequence from Paraíba, dated 2020-10-01), EPI_ISL_1715135, EPI_ISL_ 1821257, and EPI_ISL_2249437.

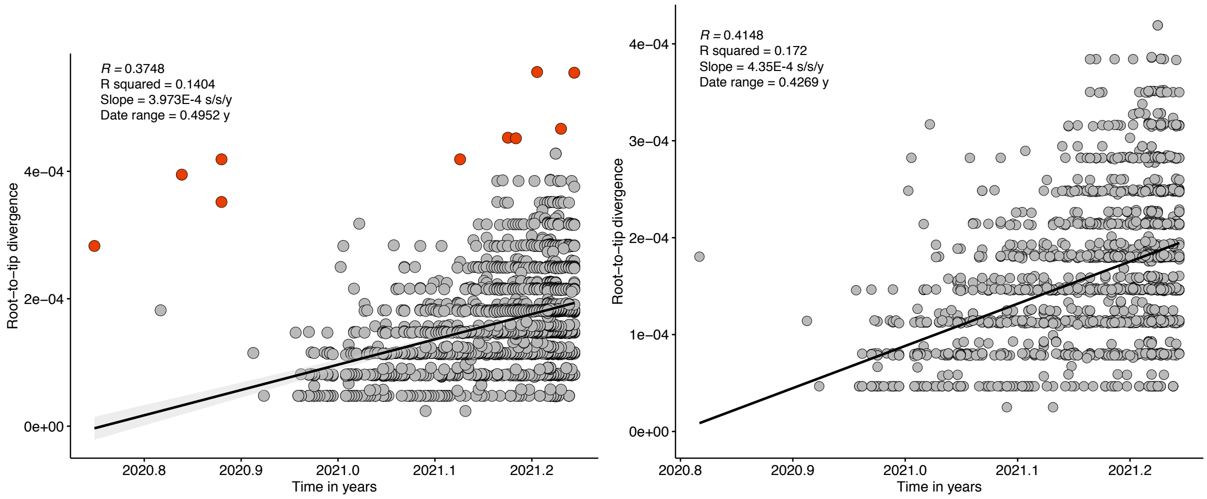

**(a)** With outliers **(b)** No outliers

**Figure S22:** Root-to-tip regression of genetic divergence against dates of sample collection. Red tips in panel a) correspond to the 10 sequences discarded from subsequent analysis. Grey tips in figure b) correspond to sequences kept for subsequent phylogenetic analysis.

#### S2.3 Bayesian phylogenetic analysis

Estimating time trees for large alignments can be computationally intractable. Thus we follow a computation strategy similar to du Plessis et al. [19] and Gutierrez et al. [20] that involves: (i) estimating an evolutionary rate using a subsample of the genome dataset of interest, and (ii) using a simpler computational approach to estimate time trees for the complete genome dataset. For step (i), we randomly selected a maximum of 20 sequences per state (except for Paraíba and Rondônia, which only had 15 and 9 sequences available during the study period). This generated a dataset of 264 genomes sequences. Sequences with earliest and latest dates of collection from each state were kept in the alignment to increase temporal signal of the resulting dataset. We used BEAST v.1.10.4 [21] to estimate an evolutionary rate under a Hasegawa Kishino-Yano (HKY) [22] substitution model and a strict molecular clock with a continuous-time Markov chain prior. We used a Bayesian skygrid with 10 grid points as a demographic tree prior [23]. The BEAST xml file is available at inst/utils/BEAST_thorney_P1.xml in our https://github.com/CADDE-CENTRE/covid19_brazil_hfr repository. Four Markov Chain Monte Carlo (MCMC) chains were run for 50 million steps, sampling parameters and trees every 50,000 steps. Convergence of the MCMC chains was assessed using Tracer v.1.7 [24]. For step (ii), the complete dataset was analysed using BEAST v1.1.0.5 [25] using a newly developed approach that significantly reduces computation time. This approach takes in a rooted phylogenetic ML tree (instead of an alignment) and rescales its branches into time units. The likelihood of each branch length is modelled as a Poisson distribution with a mean that is directly proportional to the clock rate [26, 27]. We used a rate of $4.864 \times 10^{-4}$ substitutions/site/year based on the median clock rate estimate obtained from step (i). We defined a coalescent Skygrid prior and used the best-fitting IQTree ML tree rooted in TempEst as a starting data tree. Two independent MCMC chains were run for 1,000 million MCMC steps and combined after discarding 10% of the run as burn-in to generate an empirical posterior tree distribution. Convergence was assessed using Tracer v.1.7 [24].

##### S2.4 Estimation of earliest date of SARS-CoV-2 Gamma circulation in each location

We used a 14-state asymmetric discrete Bayesian molecular clock phylogeographic approach [28] implemented in BEAST v.1.0.4 [21] to infer ancestral state locations on an empirical distribution of 500 posterior time-trees. For sequences with known travel history, we assigned the state of infection instead of state of reporting. We estimated unknown state locations for the sequences collected at the International Guarulhos Airport in São Paulo. We tracked the complete jump history of viral movement events between each pair of states [29, 30, 31]. We used a recently developed tool, the TreeMarkovJumpHistoryAnalyzer, which collects Markov jumps and their timings from a posterior tree distribution with Markov jump histories [32], available at inst/utils/P.1_MJumps_complete_history.xml in our https://github.com/CADDE-CENTRE/covid19_brazil_hfr repository. To date the most common recent ancestor of the earliest local transmission cluster in each state, we used a customised R script to summarise the posterior probability distribution densities for the earliest time of the introduction leading to $\geq$2 descendants in each of the 14 states. Throughout, we denote for each location the posterior median of the earliest week in which Gamma was estimated to circulate locally by $W_l^{\text{emerge}-\Gamma}$, and report the posterior median emergence dates in Supplementary Table S4.

##### S2.5 Tracking Gamma's expansion in each location

To assess the spatio-temporal expansion of Gamma in each of the 14 cities, we based our investigations on Gamma's variant frequency in SARS-CoV-2 sequence samples obtained in each location. All 7,221 sequences described in Section S1.2 were used for this analysis, and aggregated by week $w$ of sample collection in each location $l$. We assumed that the frequency of Gamma in state capitals is similar to the frequency of Gamma across that state, measured by data available in GISAID and metadata available from Rede Genômica FioCruz that provides more representative sampling [5]. We denote the week index in which Gamma was in each location first detected by $W_l^{\text{detect}-\Gamma}$ (see Supplementary Table S4), the number of sequenced genotypes in location $l$ and collection week $w$ by $s_{l,w}$, and the number sequenced genotypes attributed to the Gamma variant with Pangolin by $s_{l,w}^{\Gamma}$. Figure S18 indicates the sample sizes in each location, and Figure S1 shows the proportion of Gamma sequences over time in each location.

#### S3 Controlling for hospitalised patients with unknown outcomes

To characterise in-hospital fatality rates, we focused on the weekly COVID-19 attributable hospital admissions $h_{l,a,w}^{\text{res}}$ among residents with no evidence of vaccination prior to hospitalisation in each location $l$ and age band $a$, and their reported clinical outcomes as of 19 September 2021 (Section S1.6). Patients were either discharged alive, died, or had an unknown outcome, which respectively we denote by $h_{l,a,w}^{\text{res-A}}$, $h_{l,a,w}^{\text{res-D}}$ and $h_{l,a,w}^{\text{res-U}}$. Figure 2 in the main text and Figure S23 illustrate that a large number of hospitalised patients had unreported outcomes, typically since Gamma's detection in each location, which make interpretation of the raw data challenging. A

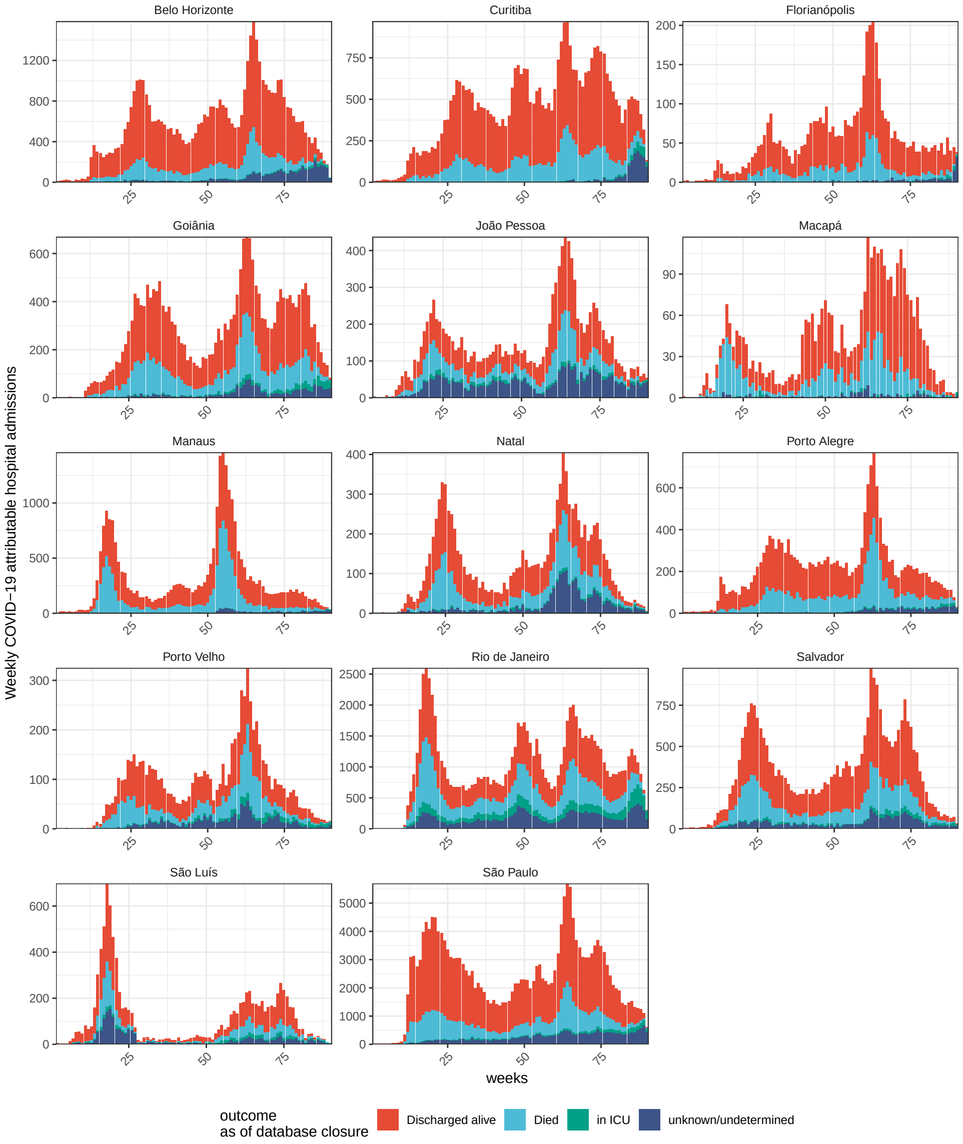

**Figure S23:** Clinical outcomes of COVID-19 attributable hospitalised admissions in unvaccinated residents in 14 cities in Brazil. Data are from the SIVEP-Gripe platform of the Brazilian Ministry of Health and as of 20 September 2021. The weekly COVID-19 attributable hospital admissions among residents are shown by outcome as of database closure.

subset of individuals withunknown outcome had an ICU start date and no ICU end date, and are in the figure shown separately. This section provides technical details on how death counts were adjusted for underreporting in hospitalised patients with unknown outcomes.

First, we calculated the proportion of deaths among patients with known outcomes, for each location and age band over the two weeks prior to the current week of interest. Second, we projected COVID-19 attributable deaths among COVID-19 attributable hospitalised residents with currently unknown outcomes based on this time-varying and age-specific proportion. Specifically, we calculated a 2-week rolling mean version of daily COVID-19 fatality rates among individuals with known outcomes in location $l$, age band $a$ and day $t$ via

$$z^{\text{known-rolling}}_{l,a,t} = \frac{\frac{1}{14}\sum_{s=t-14}^{t} h^{D}_{l,a,s}}{\frac{1}{14}\sum_{s=t-14}^{t} h^{A}_{l,a,s} + h^{D}_{l,a,s}}. \tag{S9}$$

Equation (S9) was calculated over all COVID-19 attributable hospitalised patients regardless of residency because of larger sample sizes in the numerator and denominator, and assuming that fatality rates among residents and non-residents are the same in the same location and same age band. Next, we used (S9) to project COVID-19 attributable deaths among COVID-19 attributable hospitalisations in residents in location $l$, age band $a$ and day $t$ who currently have an unknown outcome, and then summed for each week $w$,

$$h^{\text{res-proj-D}}_{l,a,w} = \sum_{t\in w} z^{\text{known-rolling}}_{l,a,t} h^{\text{res-U}}_{l,a,t}. \tag{S10}$$

This allows us to estimate the number of hospitalised residents in location $l$, age band $a$ and week $w$ with no evidence of vaccination prior to hospitalisation who are observed or expected to have a fatal outcome as

$$h^{\text{res-adj-D}}_{l,a,w} = h^{\text{res-D}}_{l,a,w} + h^{\text{res-proj-D}}_{l,a,w}, \tag{S11}$$

where $h^{\text{res-D}}_{l,a,w}$ are those with an observed fatal outcome (Section S1.6).

#### S4 Smoothed age-standardised COVID-19 in-hospital fatality rates

We describe the extent of spatiotemporal variation in the weekly COVID-19 fatality rate data with a simple measure, smoothed age-standardised in-hospital fatality rates. We age-standardise to summarise data across age bands into a simple time series statistic, and to control for differences in the age composition of city populations. This section presents technical details on how the smoothed, non-parametric estimates of age-standardised in-hospital fatality rates were constructed.

We start with the under-reporting adjusted COVID-19 attributable deaths in residents in location $l$, age band $a$ that were hospitalised in week $w$, $h^{\text{res-adj-D}}_{l,a,w}$, Equation (S11), and the COVID-19 attributable hospital admissions in residents without evidence of vaccination prior to hospital admission, $h^{\text{res}}_{l,a,w}$, (Section S1.6). Then, we define the age-specific in-hospital fatality rates in location $l$ and age band $a$ in week $w$ as

$$z_{l,a,w} = h^{\text{res-adj-D}}_{l,a,w} \Big/ h^{\text{res}}_{l,a,w} \tag{S12}$$

whenever the denominator $h^{\text{res}}_{l,a,w}$ is non-zero. Figure 1C in the main text illustrates the empirical in-hospital fatality rates in Manaus and in each age band. To standardise across age bands, we summed (S12) using as weights

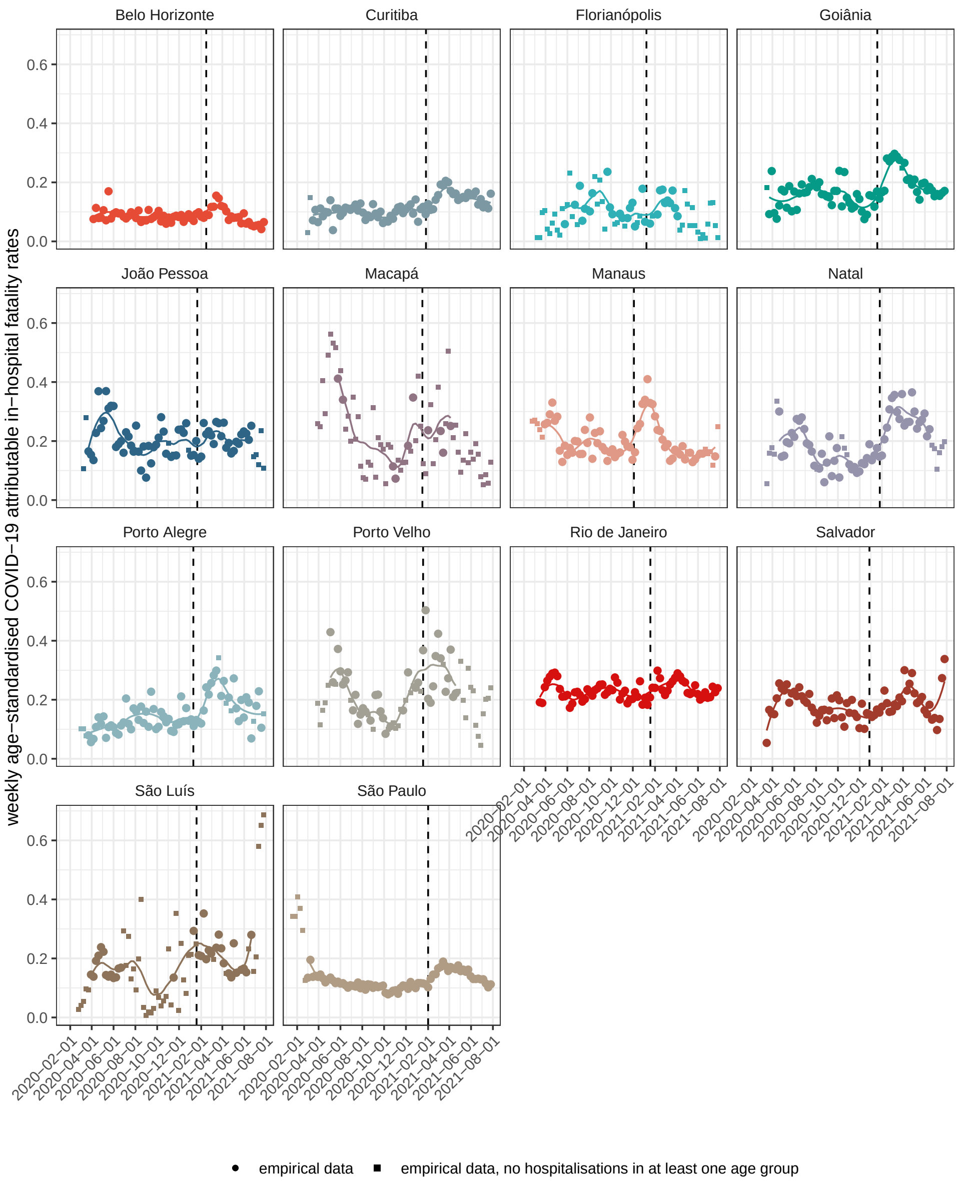

**Figure S24:** Age-standardised in-hospital COVID-19 fatality rates. The empirical, age-standardised fatality rates are shown as circles whenever the weekly age-specific fatality rates are well defined for all age groups (i. e. there was at least one weekly hospital admission for that given age group, week and location). Whenever there were undefined age-specific fatality rates, we computed the empirical age-standardised fatality rates by setting the undefined values to the closest defined value in time (squares). The smoothed, non-parametric estimates of the age-standardised in-hospital fatality rates are shown as a solid line. The date of Gamma's detection is indicated with vertical dashed lines.

the proportion of each age band in the population across all cities considered. Specifically, we calculate the unsmoothed, age-standardised COVID-19 attributable in-hospital fatality rate among residents in location $l$ in week $w$ as

$$z_{l,w} = \sum_a \frac{n_a^{\text{cities}}}{\sum_b n_b^{\text{cities}}} z_{l,a,w}, \tag{S13}$$

where $n_a^{\text{cities}}$ is the population in age band $a$ across all cities considered. If $z_{l,a,w}$ is not well defined for a given age group $a$, we instead use $z_{l,a,f_{l,a}(w)}$, where $f_{l,a}$ maps the input to the closest week with $h_{l,a,w}^{\text{res}} > 0$. In cities such as São Luís, most of the weekly age-standardised in-hospital fatality rates needed this adjustment, as can be observed in Supplementary Figure S24.

Due to the presence of weeks with zero or few COVID-19 attributable hospital admissions for specific age bands, the age-standardised in-hospital fatality rates were noisy. To obtain a more stable estimate of the age-standardised in-hospital fatality ratio, we first applied a non-parametric smoother to the age-specific in-hospital fatality rates, Equation (S12). Specifically, we used the loess smoother as implemented in the R package `stats` with input arguments span set to $0.3$. Second, we age-standardised the non-parametric smoothes in analogy to Equation (S13). To avoid extrapolating when computing extrema of the smoothed in-hospital fatality rates, we only considered estimates obtained in the window between the first and the last weeks with well-defined empirical age-standardised in-hospital fatality rates. Figure S24 illustrates the smoothed, age-standardised in-hospital fatality rates as a line.

#### S5 Healthcare pressure indices

In the context of substantial underfunding prior to the pandemic [33, 34] and disparities in health care resources across and within Brazil's states [35], we introduce pandemic healthcare pressure indices to monitor in-hospital health care load at city level. This section provides technical details of these healthcare pressure indices. All indices are constructed as a time series measure of healthcare demand per available resource.

We start by considering the number of adult ICU beds in location $l$ and week $w$, $r_{l,w}^{\text{ICU-beds}}$ (Section S1.8), and the number of ICU admissions among residents and non-residents in location $l$ and week $w$, which we denote by $h_{l,w}^{\text{ICU}}$, and which are reported in SIVEP-Gripe (Section S1.4). Note that here we consider all ICU admissions of residents and non-residents, because all ICU admissions add to demand in hospitals. Our first index, ICU admissions per ICU bed, was then defined by

$$x_{l,w}^{\text{ICU-adm-per-ICU-bed-00}} = h_{l,w}^{\text{ICU}} \Big/ r_{l,w}^{\text{ICU-beds}}. \tag{S14}$$

To account for the fact that additional ICU admissions in prevoius or subsequent weeks could have an adverse impact on fatality rates of patients admitted in the current week, we also considered rolling sums of the form

$$\begin{aligned} x_{l,w}^{\text{ICU-adm-per-ICU-bed-0j}} &= \Big(\sum_{i=0}^{j} h_{l,w+i}^{\text{ICU}}\Big) \Big/ r_{l,w}^{\text{ICU-beds}}, \\ x_{l,w}^{\text{ICU-adm-per-ICU-bed-j0}} &= \Big(\sum_{i=-j}^{0} h_{l,w+i}^{\text{ICU}}\Big) \Big/ r_{l,w}^{\text{ICU-beds}}, \\ x_{l,w}^{\text{ICU-adm-per-ICU-bed-jj}} &= \Big(\sum_{i=-j}^{j} h_{l,w+i}^{\text{ICU}}\Big) \Big/ r_{l,w}^{\text{ICU-beds}}, \end{aligned} \tag{S15}$$

for $j = 1, \ldots, 4$. Of these we retained for further analysis the version that correlated most strongly with bi-weekly age-standardised in-hospital fatality rate analogous to the weekly estimates in equation (S13). For the ICU admissions per ICU bed index, this was the rolling sum across the current and following two weeks.

All other healthcare pressure indices were constructed in the same way. These are the number of ICU admissions in this and the following 2 weeks per physician, the number of ICU admissions in this and the following 2 weeks per intensive care specialist, the number of ICU admissions in this and the following 2 weeks per nurse, the number of ICU admissions in this and the following 2 weeks per nurse assistant, the number of ICU admissions in this and the following 2 weeks per physiotherapist, and the number of ICU admissions in this and the following 2 weeks per ventilator, and the number of SARI hospital admissions in this and the following 2 weeks per critical care bed, the number of SARI hospital admissions in this and the following 2 weeks per critical care bed with a ventilator, the number of SARI hospital admissions in this and the following 2 weeks per ventilator, the number of SARI hospital admissions in this and the following 2 weeks per nurse, and the number of SARI hospital admissions in this and the following 2 weeks per physician, respectively defined by

$$x_{l,w}^{\text{ICU-adm-per-physician-02}} = \left( \sum_{i=0}^{2} h_{l,w+i}^{\text{ICU}} \right) \Big/ n_{l,w}^{\text{physicians}} \tag{S16a}$$

$$x_{l,w}^{\text{ICU-adm-per-intensivist-02}} = \left( \sum_{i=0}^{2} h_{l,w+i}^{\text{ICU}} \right) \Big/ n_{l,w}^{\text{intensivist}} \tag{S16b}$$

$$x_{l,w}^{\text{ICU-adm-per-nurse-02}} = \left( \sum_{i=0}^{2} h_{l,w+i}^{\text{ICU}} \right) \Big/ n_{l,w}^{\text{nurses}} \tag{S16c}$$

$$x_{l,w}^{\text{ICU-adm-per-nurse-assist-02}} = \left( \sum_{i=0}^{2} h_{l,w+i}^{\text{ICU}} \right) \Big/ n_{l,w}^{\text{nurse-assist}} \tag{S16d}$$

$$x_{l,w}^{\text{ICU-adm-per-physiotherapist-02}} = \left( \sum_{i=0}^{2} h_{l,w+i}^{\text{ICU}} \right) \Big/ n_{l,w}^{\text{physiotherapists}} \tag{S16e}$$

$$x_{l,w}^{\text{ICU-adm-per-ventilator-02}} = \left( \sum_{i=0}^{2} h_{l,w+i}^{\text{ICU}} \right) \Big/ n_{l,w}^{\text{ventilators}}, \tag{S16f}$$

and

$$x_{l,w}^{\text{SARI-adm-per-crit-care-bed-02}} = \left( \sum_{i=0}^{2} h_{l,w+i}^{\text{SARI}} \right) \Big/ n_{l,w}^{\text{crit-care-beds}} \tag{S17a}$$

$$x_{l,w}^{\text{SARI-adm-per-crit-care-bed-vent-02}} = \left( \sum_{i=0}^{2} h_{l,w+i}^{\text{SARI}} \right) \Big/ n_{l,w}^{\text{crit-care-beds-vent}} \tag{S17b}$$

$$x_{l,w}^{\text{SARI-adm-per-ventilator-02}} = \left( \sum_{i=0}^{2} h_{l,w+i}^{\text{SARI}} \right) \Big/ n_{l,w}^{\text{ventilators}} \tag{S17c}$$

$$x_{l,w}^{\text{SARI-adm-per-nurse-02}} = \left( \sum_{i=0}^{2} h_{l,w+i}^{\text{SARI}} \right) \Big/ n_{l,w}^{\text{nurses}} \tag{S17d}$$

$$x_{l,w}^{\text{SARI-adm-per-physician-02}} = \left( \sum_{i=0}^{2} h_{l,w+i}^{\text{SARI}} \right) \Big/ n_{l,w}^{\text{physicians}}, \tag{S17e}$$

where $h^{\text{SARI}}_{l,w}$ are the number of SARI hospital admissions among residents and non-residents in location $l$ and week $w$, $n^{\text{physicians}}_{l,w}$ are the number of physicians, $n^{\text{nurses}}_{l,w}$ are the number of nurses, $n^{\text{nurse-assist}}_{l,w}$ are the number of nurse assistants, $n^{\text{physiotherapists}}_{l,w}$ are the number of physiotherapists, $n^{\text{intensivist}}_{l,w}$ the number of intensive care specialists, $n^{\text{crit-care-beds}}_{l,w}$ the number of critical care beds, $n^{\text{crit-care-beds-vent}}_{l,w}$ the number of critical care beds controlled for ventilators and $n^{\text{ventilators}}_{l,w}$ the number of ventilators in location $l$ and week $w$. Figures S6 illustrate that most of these simple healthcare demand indicators correlate strongly with in-hospital fatality rates.

In our inference framework described in Section S7, we embed the indices (S16) within a variable selection framework to mitigate the fact that the indices that strongly correlated with each other. For this purpose, we standardised the indices for use in the Bayesian multi-strain fatality as follows. First, we identify for each location the week in which the smoothed, age-standardised in-hospital fatality rates (S13) prior to Gamma's detection were lowest,

$$\tilde{w}_l = \text{argmin}_{w=1:(W^{\text{detect}-\Gamma}_l-1}z_{l,w}, \tag{S18}$$

where $W^{\text{detect}-\Gamma}_l$ denotes the week index in which the Gamma variant was first detected in location $l$ (Section S1). Then, we standardise the $p$th indicator in (S16) according to

$$x^{\text{indicator-std}}_{l,p,w} = \left(x^{\text{indicator}}_{l,p,w} - x^{\text{indicator}}_{l,p,\tilde{w}_l}\right)\Big/\text{sd}\left(x^{\text{indicator}}_{l,p,w}\right), \tag{S19}$$

where $\text{sd}\left(x^{\text{indicator}}_{l,p,w}\right)$ denotes the standard deviation of the $p$th indicator across all weeks in location $l$. Thus, the standardised healthcare indicators evaluate to zero in week $\tilde{w}_l$, and are constructed to describe increases in in-hospital fatality rates relative to the minimum age-standardised in-hospital fatality rate that was achieved at some point prior to Gamma's detection in each location.

### S6 Controlling for pandemic associated mortality and vaccine roll-out

Figure S25 illustrates that the share of age groups among hospital admissions changed substantially over time, with the share of older age groups among hospital admissions declining steadily in recent months. To assess the extent to which changes in the age composition of deaths after Gamma's detection are driven by factors unrelated to Gamma, we further quantified time trends in the population at risk of COVID-19 hospitalisation, that were then used as input into the Bayesian multi-strain fatality model described in Section S7. First, to reflect high fatality rates among older individuals and resulting changes to the population age composition, we subtracted accumulating COVID-19 attributable deaths from the 2020 population denominators. Second, to reflect increased protection to COVID-19 hospitalisation after vaccination, we accounted for increasing proportions of the population that had received their first and second vaccine doses.

#### S6.1 Controlling for pandemic associated mortality

We adjusted the population at risk for cumulative COVID-19 attributable mortality in each location in several steps. First, we considered all COVID-19 attributable deaths in hospitals, which we obtained from the under-reporting adjusted death counts among hospitalised patients that are resident in each location, and which were reported to SIVEP-Gripe (Section S1.7). Second, we considered all COVID-19 attributable out-of-hospital deaths among

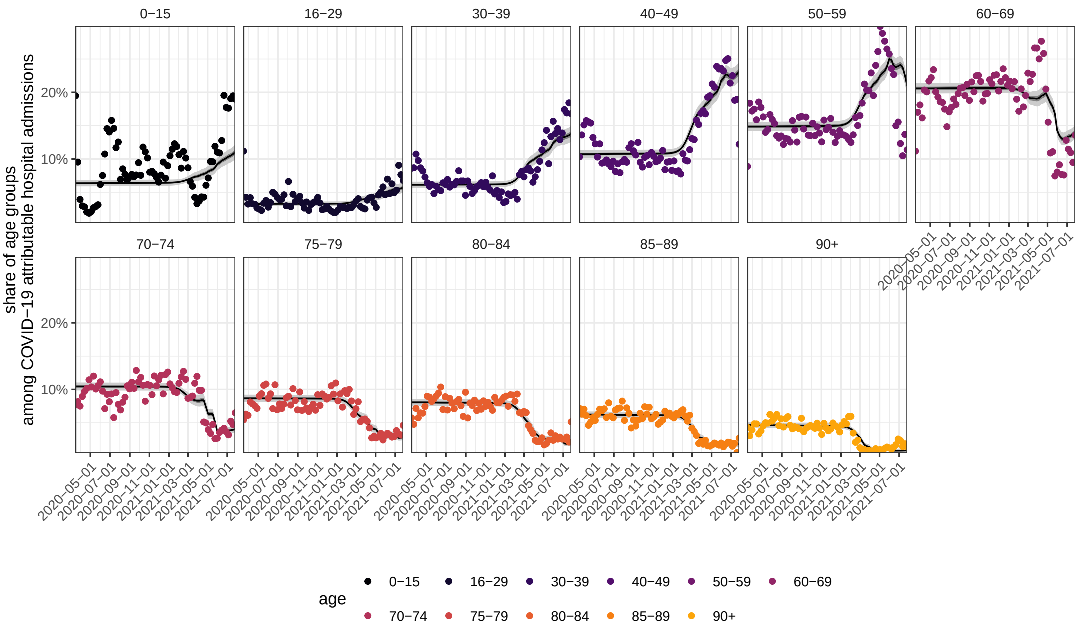

**Figure S25:** Temporal shifts in the age composition of COVID-19 attributable hospital admissions in Rio de Janeiro. The empirical, weekly proportions are shown as dots in colour. Posterior median estimates of the expected age composition of hospital admissions, as computed in Equation S37, are shown as a black line along with 95% credible intervals.

residents in each location, which were reported to SIVEP-Gripe. Third, we used records from Brazil's Civil Registry to calculate weekly excess deaths among residents of each location. Then, we compared the excess deaths to the in-hospital and out-of-hospital COVID-19 attributable deaths reported to SIVEP-Gripe, and used whichever number was larger in each week.

Starting with the in-hospital COVID-19 attributable deaths, we know all observed deaths in residents (Section S1.7), and calculated the expected number of additional deaths among hospitalised residents with unknown outcomes (Section S3). To attribute an expected death date to the expected number of additional deaths, we estimated for each city characteristic time distributions from the time of hospital admission to COVID-19 attributable death. The times from hospital admissions to deaths varied by age and location. For this reason we estimated the time distributions independently for each location-age group. Overall, Gamma distributions fitted by maximum likelihood with the `fitdist` function in the `fitdistrplus` package, version 1.0.14, provided the closest match to the empirical data in terms of quantile-quantile plots when compared to Lognormal, Weibull, and Pareto distributions. The resulting Gamma distributions $g^{\text{h2d}}(s;\eta^{\text{h2d}}_{l,a})$ with maximum likelihood parameter estimates $\hat{\eta}^{\text{h2d}}_{l,a}$ for each location and age band were then time-discretized and re-normalized using

$$g^{\text{h2d}}_{l,a,s} = \left(\int_s^{s+1} g^{\text{h2d}}(u;\hat{\eta}^{\text{h2d}}_{l,a})du\right) \Big/ \left(\int_1^{65+1} g^{\text{h2d}}(u;\hat{\eta}^{\text{h2d}}_{l,a})du\right) \quad \text{(S20)}$$

for $s = 1, \dots, 65$ days, where the integral in the denominator is the normalising constant. The resulting time-discretized Gamma distributions provided a good fit against the empirical data for some locations. The estimated distributions of the time from hospital admission to death sum to $1$, which we used in turn to attribute each projected death in location $l$, age band $a$ and day $t$ over future days $t+s$ according to $\delta_{l,a,t}\, h^{\text{res-U}}_{l,a,t}\, g^{\text{h2d}}_{l,a,s}$, where $s$ was in $s=1,\ldots,65$ days. The projected COVID-19 attributable deaths among hospitalised residents in location $l$ and age band $a$ in week $w$ with as of yet unknown outcomes were calculated through

$$d^{\text{res-hosp-unknown-outcome}}_{l,a,w} = \sum_{t\in w}\sum_{s=0}^{65} z^{\text{known-rolling}}_{l,a,t-s}\; h^{\text{res-U}}_{l,a,t-s}\, g^{\text{h2d}}_{l,a,s}, \tag{S21}$$

and added to the observed in-hospital and out-of-hospital COVID-19 attributable deaths that are reported to SIVEP-Gripe, and which are described in Section S1.7,

$$d^{\text{res-SIVEP-adj}}_{l,a,w} = d^{\text{res-SIVEP}}_{l,a,w} + d^{\text{res-hosp-unknown-outcome}}_{l,a,w}. \tag{S22}$$

So, while the deaths in (S11) are counted by week of hospital admission, the deaths in (S22) are counted by week of death. Figure S3 illustrates the expected deaths in hospitalised residents with unknown outcomes, Equation (S21), for each of the 14 cities in blue grey.

We further checked for additional under-reporting by comparing (S22) against the excess deaths in residents in each location, which we derived from longitudinal death records reported by Brazil's Civil Registry (see Section S1.9). The death records in the Brazilian Civil Registry include both residents and non-residents, and so could not be directly compared to the underreporting-adjusted, COVID-19 attributable deaths among residents that are derived from the SIVEP-Gripe data, equation (S22). We calculated the proportion $\rho^{\text{res-COVID-deaths}}_{l,a,w}$ of the number of COVID-19 attributable deaths in location $l$ and age band $a$ in week $w$ that occurred among residents,

$$\rho^{\text{res-COVID-deaths}}_{l,a,w} = d^{\text{res}}_{l,a,w}/d_{l,a,w}, \tag{S23}$$

where the COVID-19 attributable deaths among residents and non-residents, $d_{l,a,w}$, and among residents, $d^{\text{res}}_{l,a,w}$, are described in Section S1.7. The proportion was set to zero whenever $d_{l,a,w}$ was zero. We then compared the excess deaths and COVID-19 related deaths from Brazil's Civil Registry to the underreporting-adjusted, COVID-19 attributable deaths derived from SIVEP-Gripe in location $l$, age band $a$ and week $w$ through

$$\begin{aligned} d^{\text{res-under-reported}}_{l,a,w} = \max\Big(&\rho^{\text{res-COVID-deaths}}_{l,a,w} v^{\text{excess}}_{l,a,w},\; \rho^{\text{res-COVID-deaths}}_{l,a,w} v^{\text{covid}}_{l,a,w},\; d^{\text{res-SIVEP-adj}}_{l,a,w}\Big) \\ &- d^{\text{res-SIVEP-adj}}_{l,a,w}, \end{aligned} \tag{S24}$$

where $d^{\text{res-cens-adj}}_{l,a,w}$ are defined in (S22). Finally, we calculated the underreporting-adjusted COVID-19 attributable deaths among residents in location $l$, age band $a$ and week $w$ based on Civil Registry data through

$$d^{\text{res-SIVEP-Registry-adj}}_{l,a,w} = d^{\text{res-SIVEP-adj}}_{l,a,w} + d^{\text{res-under-reported}}_{l,a,w}. \tag{S25}$$

Figure S3 illustrates the expected number of COVID-19 attributable deaths that are derived by comparison to the Civil Registry records, equation (S24), in red.

To check the final estimate of COVID-19 attributable deaths in residents, Equation (S25), we sought to compare the all-cause deaths in the Brazilian civil registry data to other independent data sets reporting on the number of deaths in 2020 and 2021 in Brazilian state capitals. For Manaus, we were able to retrieve the daily number of public and private burials from the Mayor's office of Manaus. The daily data was published on the official website

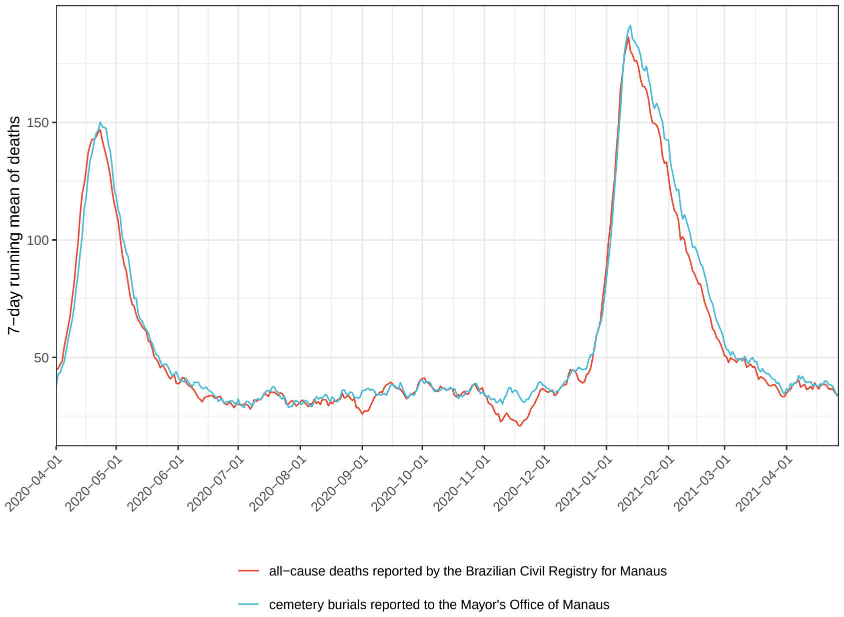

**Figure S26:** Reported all-cause deaths in Manaus. To check our adjustments to the reported COVID-19 attributable deaths in SIVEP-Gripe for likely under-reporting, we compared records of private and public burials to deaths reported in the Brazilian Civil Registry. Shown is the 7-day running average of all-cause deaths reported in the Brazilian Civil Registry, and the 7-day running average of burial records.

of Manaus' Mayor office [36] from April 1, 2020 to June 30, 2020, while it was published on the Amazonas State Health Surveillance Foundation website [37] starting from October 27, 2020. To fill the hiatus, we used data from the annual report on daily cemetery burials by the Mayor's office [38]. Figure S26 shows the 7-day running mean of the daily burials against the 7-day running mean of all-cause deaths reported in the Brazilian Civil Registry for Manaus. The burials lagged the registered deaths as can be expected by a few days, but otherwise were very similar in magnitude, indicating that at least for Manaus, our approach to accounting for likely underreporting of COVID-19 attributable deaths in the SIVEP-Gripe data is consistent with public and private burial records.

Finally, we downwards adjusted the 2020 population size projections $n_{l,a}$ (Section S1.3) by the expected, cumulated COVID-19 attributable deaths after adjusting for unknown outcomes in hospitalised residents and for differences to the Civil Registry data,

$$n^{A}_{l,a,w} = n_{l,a} - \sum_{s=1}^{w} d^{\text{res-SIVEP-Registry-adj}}_{l,a,s}, \tag{S26}$$

where $d^{\text{res-SIVEP-Registry-adj}}_{l,a,w}$ are defined in Equation (S25). Figure S27 illustrates the corresponding cumulated mortality rates, $\big(\sum_{s=1}^{w} d^{\text{res-SIVEP-Registry-adj}}_{l,a,s}\big)/n_{l,a}$, for each location and age group. As of 26 July 2021, we find cumulated mortality rates that exceed those in many other countries such as the US or the UK. Yet, the cumulated loss of life alone is too small to account for the much larger observed shifts in the age composition of deaths since Gamma's detection.

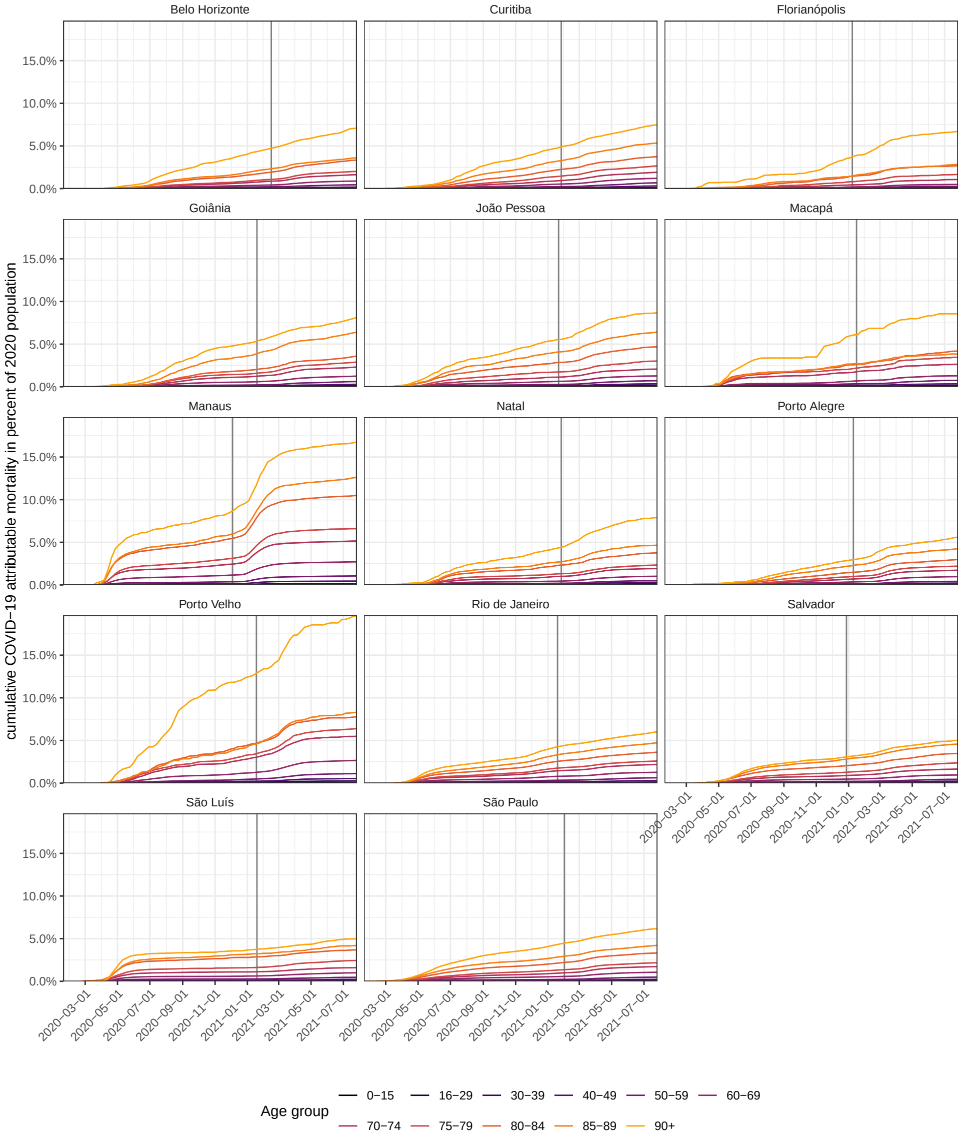

**Figure S27:** Cumulated COVID-19 attributable mortality in per cent of age-specific populations. For each location, the expected COVID-19 attributable deaths after adjusting for unknown outcomes in hospitalised residents and for differences to the Civil Registry data were cumulated over time and divided by 2020 population size estimates for each age band. The date of Gamma's detection is shown as grey vertical line.

| Vaccine | Dose | Estimated vaccine efficacy against symptomatic infection 14 days since administration | Notes and reference |
|---|---|---|---|
| Sinovac | 1 | 10.5% (-4.4-23.3%) | Performed between 17/01/21 to 29/04/21<br>In Brazil with presence of the Gamma variant.<br>Subjects were 70 years old or older. Ref [39]. |
| Sinovac | 1 | 49.6% (11.3-71.4%) | Performed between 19/01/21 to 25/03/21<br>In Brazil with presence of the Gamma variant.<br>Subjects were healthcare workers. Ref. [40]. |
| Sinovac | 2 | 41.6% (26.9-53.3%) | Performed between 17/01/21 to 29/04/21<br>In Brazil with presence of the Gamma variant.<br>Subjects were 70 years old or older. Ref. [39]. |
| Sinovac | 2 | 36.8% (-54.2-74.2%) | Performed between 19/01/21 to 25/03/21<br>In Brazil with presence of the Gamma variant.<br>Subjects were healthcare workers. Ref. [40]. |
| Sinovac | 2 | 50.7% (33.3-62.5%) | Performed between 23/02/2020 to 28/03/21<br>In Brazil with presence of the Gamma variant.<br>Subjects were healthcare workers. Ref. [41]. |
| Covishield | 1 | 76.0% (59-85.9%) | Performed between 23/02/2020 to 28/03/21<br>In UK, South Africa and Brazil,<br>But results do not concern Gamma variant<br>Subjects were 18 year old or older. Ref. [42]. |
| Covishield | 1, 2 | 21.9% (-49.9-59.8%) | Performed between 24/06/20 to 09/11/20<br>In South Africa with presence of the Gamma variant.<br>Subjects were 18 to 65 years old. Ref. [43]. |
| Covishield | 2 | 63.6% (-2.1-87%) | Performed between 06/21 to 02/21<br>In Brazil with presence of the Gamma variant.<br>Subjects were 18 years old or older. Ref. [44]. |
| Covishield | 2 | 56.9% (-15.5-83.9%) | Performed between 06/21 to 02/21<br>In Brazil with presence of the Gamma variant.<br>Subjects were 18 years old or older. Ref. [44]. |
| Covishield | 2 | 10.4% (-76.8-54.8%) | Performed between 24/06/20 to 09/11/20<br>In South Africa with presence of the Gamma variant.<br>Subjects were 18 to 65 years old. Ref [43]. |
| Covishield | 2 | 66.7% (57.4-74%) | Performed between 23/02/2020 to 28/03/21<br>In UK, South Africa and Brazil,<br>Results do not concern Gamma variant exclusively<br>Subjects were 18 year old or older. Ref. [42]. |

**Table S6:** COVID-19 vaccine efficacy estimates in populations in which SARS-CoV-2 Gamma is circulating.

#### S6.2 Controlling for vaccine roll-out

To estimate vaccine coverage in residents of each population, we calculated the proportion of vaccinated individuals in the estimated, mortality-adjusted populations in each location. Specifically, we calculated vaccine coverage with $i$ doses of vaccine $k$ in age group $a$ in location $l$ by week $w$ through

$$p_{l,a,w}^{\text{vaccinated-}k,i} = v_{l,a,w}^{k,i} / n_{l,a,w}^{A}. \tag{S27}$$

Supplementary Figure S7 shows that vaccine coverage increased rapidly over time and highlights qualitative differences in vaccination roll-out across cities. First, different vaccines were administered across cities. Second, within cities, different vaccines were administered to different age groups. Third, within age bands, the average time to the second dose varied substantially across locations.

We next sought to adjust the population at risk of severe illness and hospitalisation, which is to be used in the Bayesian fatality model, for the protective effect of Brazil's vaccine roll-out. Specifically, we subtracted from $n_{l,a,w}^{A}$, Equation (S26), a proportion of the corresponding populations that were vaccinated 2 weeks earlier with one or two doses,

$$n_{l,a,w}^{R} = n_{l,a,w}^{A} - \sum_{k\in\{\text{Covishield,Sinovac,Pfizer,Janssen}\}} \sum_{i=1}^{2} \eta^{k,i} v_{l,a,w-2}^{k,i}, \tag{S28}$$

where $n_{l,a,w}^{A}$ are defined in (S26), and $\eta^{k,i} v_{l,a,w-2}^{k,i}$ describe the proportion of the population in location $l$ and age band $a$ that was vaccinated with $i$ doses of vaccine $k$ two weeks earlier, and is in the model protected from a COVID-19 hospitalisation in week $w$.

To specify $\eta^{k,i}$, we considered published and unpublished data on vaccine effectiveness that were reviewed in the systematic analysis of Imai [45] , and data from a further 6 studies, that evaluated COVID-19 vaccine effectiveness in populations in which the SARS-CoV-2 Gamma variant was circulating. Table S6 lists the vaccine efficacy estimates found following 1 or 2 doses of Covishield and Sinovac. For Sinovac, we identified one study estimating vaccine effectiveness against death in São Paulo between January and April 2021 [39]. We assumed similar vaccine effectiveness against severe infection requiring hospitalisation, and set $\eta^{\text{Sinovac},i}$ to their estimates, which are 31.6% (95% CI: 7.1-49.7%) 2 weeks post first dose, and 71.4% (95% CI: 53.7-82.3%) 2 weeks post second dose. For Covishield, we could not identify studies on vaccine effectiveness against severe infection requiring hospitalisation or deaths under our search criteria. Instead we considered estimates of vaccine effectiveness against symptomatic infection, and then calibrated these estimates to expected vaccine effectiveness against a severe outcome using the relationship reported in Figure 3A of [46]. The resulting values of $\eta^{\text{Covishield},i}$ that we use in this study are 0.4% 2 weeks post first dose, and 0.91% 2 weeks post second dose. Figure S28 illustrates the resulting changes in the proportions of the populations in each location and age band that in our model are not at risk of COVID-19 hospitalisation, $1 - n_{l,a,w}^{R} / n_{l,a,w}^{A}$. By the end of the study period only 15.07% of administered doses in the 14 state capitals were Pfizer, and even less Janssen doses were administered. Due to the low frequency, we did not identify studies estimating the vaccine effectiveness in Brazil, and so for both Janssen and Pfizer, we used the same effectiveness values as for AstraZeneca.

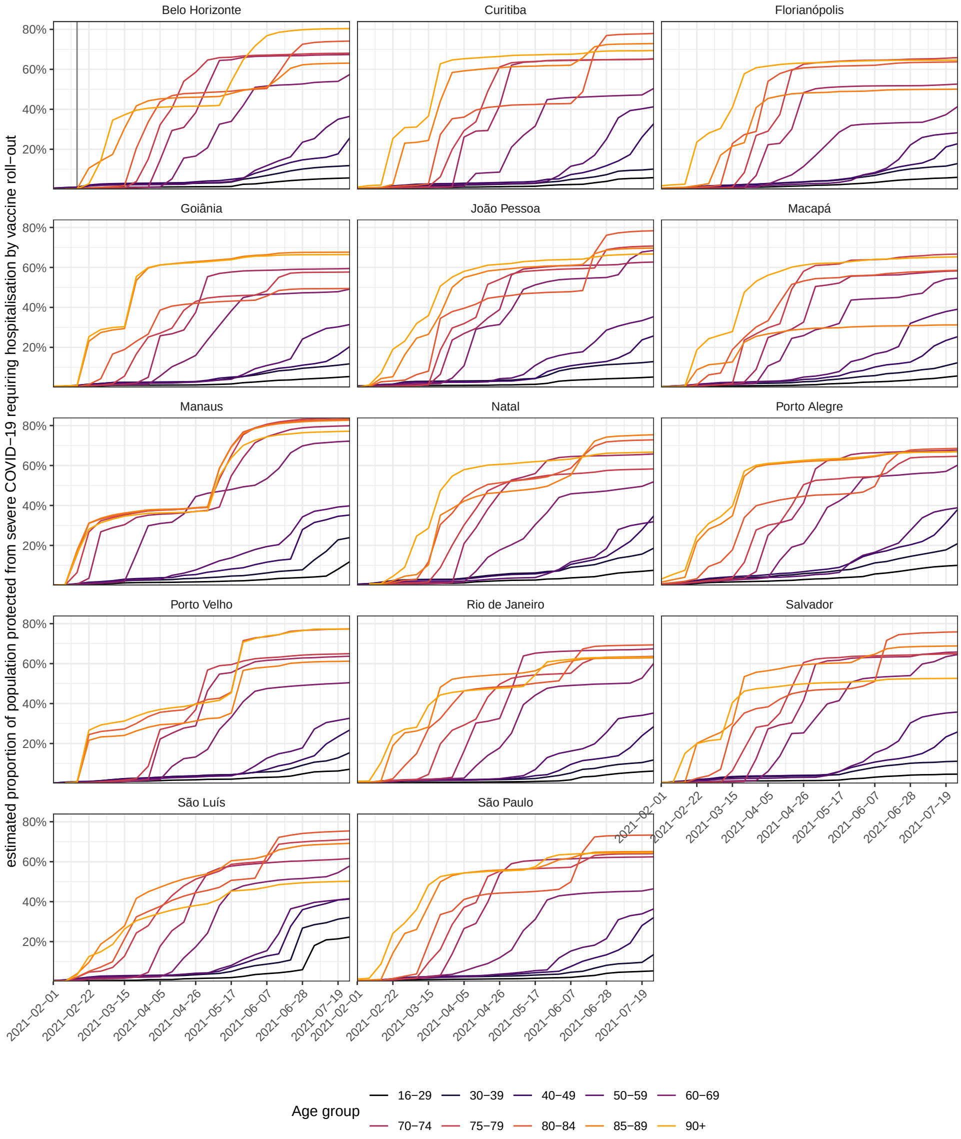

**Figure S28:** Estimated population at risk from severe COVID-19 requiring hospitalisation, after accounting for vaccine coverage. See Equation (S28) for details. Our calculations are based on individual-level data on vaccine administrations from the Brazilian Ministry of Health database, and published or calibrated estimates of vaccine effectiveness.

### S7 Statistical model to evaluate fluctuating in-hospital fatality rates

We developed a Bayesian multi-strain fatality model to disentangle the effects of pre-pandemic healthcare inequities, pandemic healthcare pressures and Gamma's disease severity on fluctuating COVID-19 in-hospital fatality rates. This section provides technical details on the model, inference, and generated quantities. Throughout, we denote by $\mathcal{A}$ the age stratification (S1), by $W_l$ the last week index in the observation period of location $l$, by $W_l^{\text{emerge-}\Gamma}$ the week index of the posterior median of the earliest week in which Gamma was estimated to circulate locally (Section S2), and by $W_l^{\text{detect-}\Gamma}$ the week index in which the Gamma variant was first detected in location $l$ (Section S1).

In Section S7.1, we describe how the model decomposes weekly, COVID-19 attributable hospital admissions by variant, based on Gamma's temporal expansion as measured by variant frequencies. The SARS-CoV-2 sequence data and sequence analyses were described in Sections S1.2 and S2. The COVID-19 attributable hospital admissions entering the model are restricted to residents in each location without evidence of vaccination prior to hospital admission, as described in Section S1.6. The hospital admissions are decomposed assuming a characteristic age profile of severe illness requiring hospitalisation, both for the Gamma and non-Gamma variants. We place this assumption in order to infer from the data if there are significant differences in the age profile of hospitalisations between variants. The primary source of information to decompose the weekly, COVID-19 attributable hospital admissions by variant are the genomic data on the SARS-CoV-2 Gamma variant frequencies over time in each location. We therefore conducted several sensitivity analyses using alternative sequence data sets to the describe local replacement dynamics, which had no discernible impact on our primary findings. See Sections S9.1 and S9.2.

Section S7.2 presents our approach for modelling associations between in-hospital fatality rates with non-parametric location effects, fixed effects associated with the healthcare pressure indices, and non-parametric virus variant effects associated with Gamma's replacement dynamics. In the model, the location effects account for social, cultural, economic or healthcare related factors that differentiate one city from another, and we interpret the ratio of fatality rates with and without Gamma's non-parametric effect as Gamma's effect size on in-hospital infection severity. We note that our list of regression predictors is not exhaustive, and so the Gamma specific effect size could be causally confounded, and merely reflects associations with fatality rates that are better explained with changes in variant frequency than with the observed healthcare pressure indices.

In Section S7.3 we explain how the model uses the linked data of weekly, COVID-19 attributable hospital admissions and the number of future deaths amongst these weekly hospitalisations to infer the extent to which location effects, pandemic healthcare pressure, and Gamma as measured on the population level through its replacement dynamics are associated with fluctuating in-hospital fatality rates. The model uses as input the underreporting-adjusted and age-specific deaths among weekly hospitalisations that are defined in (S11) in Section S3.

In Section S7.4, we then review the entire model. In Section S7.5, we present details on numerical inference of our Bayesian multi-strain model. In Section S7.6, we describe epidemiologically relevant quantities that we derive from the fitted model.

##### S7.1 Decomposition of COVID-19 hospital admissions by SARS-CoV-2 variant

To decompose COVID-19 attributable hospital admissions by variant, we model the proportion $\alpha_{l,w}$ of all-age COVID-19 attributable hospital admissions in location $l$ and week $w$ that are of variant Gamma through a logistic function, and fit the parameters of the logistic function through the number of Gamma positive samples $s^{\Gamma}_{l,w}$ out of $s_{l,w}$ sequenced or Gamma-genotyped samples over time. The model component describing the temporal expansion of Gamma in hospital admissions is

$$s^{\Gamma}_{l,w} \sim \text{Beta-Binomial}\Big(s_{l,w}, \alpha_{l,w}/\theta_1, (1-\alpha_{l,w})/\theta_1\Big) \tag{S29a}$$

$$\alpha_{l,w} = \frac{1}{1+\exp\Big(-\alpha^{\text{growth}}_l(w-\alpha^{\text{mid}}_l)\Big)} \tag{S29b}$$

$$\alpha^{\text{growth}}_l \sim \mathcal{N}(0, 0.2^2) \tag{S29c}$$

$$\alpha^{\text{mid}}_l \sim \mathcal{N}(\alpha^{\text{mid-mean}}_l, 3^2) \tag{S29d}$$

$$\alpha_{l,1:(W^{\text{emerge-}\Gamma}_l-1)} \sim \text{Normal-ccdf}(0, 0.0025^2) \tag{S29e}$$

$$\theta_1 \sim \text{Exponential}(20), \tag{S29f}$$

where the Beta-Binomial is specified in terms of the shape-shape parameterisation with mean $\alpha_{l,w}$ and variance equal to the Binomial variance component $s_{l,w}\alpha_{l,w}(1-\alpha_{l,w})$ multiplied by $\big(1+(s_{l,w}-1)\frac{1}{\theta_1^{-1}+1}\big)$ to allow for overdispersion. Time runs in units of weeks from the start of the first wave until the end of the observation period in the given location. The prior mean for the midpoint of the logistic function, $\alpha^{\text{mid-mean}}_l$, was set to the week in which the ratio $s^{\Gamma}_{l,w}/s_{l,w}$ was closest to $0.5$ in location $l$. We force Gamma's variant frequencies to close to zero before the week of Gamma's emergence in each location, which we estimate phylogenetically (Section S2). Technically this is implemented through the informative prior (S29e), where $W^{\text{emerge-}\Gamma}_l$ denotes the week including the posterior median estimate of the date of Gamma's emergence in each location, and Normal-ccdf denotes the survial function of the normal density that is parameterised by the mean and standard deviation. The prior in (S29f) was chosen to favour the least complex model with no overdispersion.

Next, we couple (S29b) to decompose the COVID-19 attributable hospital admissions $h^{\text{res}}_{l,a,w}$ among residents in location $l$ and age band $a$ in week $w$ with no evidence of vaccination prior to hospitalisation. Here, the age bands $a$ are specified in (S1) in Section S1.3, and $h^{\text{res}}_{l,a,w}$ are described in Section S1.6. We hypothesized that the Gamma and non-Gamma variants hospital admissions have a characteristic age composition, so that the mix of both characteristic age compositions according to the prevalence of Gamma and non-Gamma variants in the population provides an adequate description of the observed data $h^{\text{res}}_{l,a,w}$. Specifically, if we denote the observed sum of hospital admissions across age groups by $h^{\text{res-sum}}_{l,w} = \sum_a h^{\text{res}}_{l,a,w}$, we assume that the expected values of $h^{\text{res}}_{l,a,w}$ can be described by

$$\mathbb{E}\big(h^{\text{res}}_{l,a,w}\big) = \Big[\alpha_{l,w}\pi^{\Gamma}_a + (1-\alpha_{l,w})\pi^{\text{non-}\Gamma}_a\Big] h^{\text{res}}_{l,w}, \quad \forall a \in \mathcal{A}, \tag{S30}$$

where $\alpha_{l,w}$ is from (S29b), and the vectors $\boldsymbol{\pi}^{\Gamma} = \big(\pi^{\Gamma}_a\big)_{a\in\mathcal{A}}$ and $\boldsymbol{\pi}^{\text{non-}\Gamma} = \big(\pi^{\Gamma}_a\big)_{a\in\mathcal{A}}$ describe respectively the characteristic age compositions of the Gamma and non-Gamma variants in hospital admissions, subject to $\sum_a \pi^{\Gamma}_a = 1$ and $\sum_a \pi^{\text{non-}\Gamma}_a = 1$. However the age populations at risk of a severe outcome change over time in each location, which we described in $n^R_{l,a,w}$ in (S28) in Section S6. This leads us to model

$$\mathbb{E}\big(h^{\text{res}}_{l,a,w}\big) = \pi_{l,a,w} h^{\text{res}}_{l,w}, \quad \forall a \in \mathcal{A} \tag{S31a}$$
$$\pi_{l,a,w} = \alpha_{l,w}\pi^{\Gamma}_{l,a,w} + (1-\alpha_{l,w})\pi^{\text{non-}\Gamma}_{l,a,w} \tag{S31b}$$
$$\pi^{\Gamma}_{l,a,w} = \frac{\lambda^{\Gamma}_{l,a} n^R_{l,a,w}}{\sum_a \lambda^{\Gamma}_{l,a} n^R_{l,a,w}} \tag{S31c}$$
$$\pi^{\text{non-}\Gamma}_{l,a,w} = \frac{\lambda^{\text{non-}\Gamma}_{l,a} n^R_{l,a,w}}{\sum_a \lambda^{\text{non-}\Gamma}_{l,a} n^R_{l,a,w}}, \tag{S31d}$$

where the per-capita hospital admission rates $\lambda^{\Gamma}_{l,a}$, $\lambda^{\text{non-}\Gamma}_{l,a}$ are characteristic of the Gamma and non-Gamma variants, and constant over time. The rates $\lambda^{\Gamma}_{l,a}$, $\lambda^{\text{non-}\Gamma}_{l,a}$ may not be the same across locations, for example because of location-specific inequities in care, but we expect that the ratios $\lambda^{\Gamma}_{l,a}/\lambda^{\text{non-}\Gamma}_{l,a}$ should be constant for all age bands.

It is helpful to note that alternatively, we could also have modelled

$$\mathbb{E}\big(h^{\text{res}}_{l,a,w}\big) = \tilde{\alpha}_{l,w}\lambda^{\Gamma}_{l,a,w} n^R_{l,a,w} + (1-\tilde{\alpha}_{l,w})\lambda^{\text{non-}\Gamma}_{l,a,w} n^R_{l,a,w},$$

which leads in analogy to (S31b) to the equation

$$\pi_{l,a,w} = \tilde{\alpha}^{\Gamma}_{l,w}\frac{\lambda^{\Gamma}_{l,a} n^R_{l,a,w}}{C_{l,w}} + (1-\tilde{\alpha}^{\Gamma}_{l,w})\frac{\lambda^{\text{non-}}\Gamma_{l,a} n^R_{l,a,w}}{C_{l,w}}$$

where the normalising constant is $C_{l,w} = \sum_a \tilde{\alpha}^{\Gamma}_{l,w}\lambda^{\text{P1}}_{l,a} n^R_{l,a,w} + (1-\tilde{\alpha}^{\text{non-}}\Gamma_{l,w})\lambda^{\text{non-}\Gamma}_{l,a} n^R_{l,a,w}$. However in this parameterisation, the mixing parameters $\tilde{\alpha}_{l,w}$ do not directly correspond to the replacement dynamics $\alpha_{l,w}$ in (S29b), and for this reason we work with (S31). Finally, we allow for overdispersion in the observed admission counts and complete this model component with the same mean structure as in (S31) through

$$\boldsymbol{h}^{\text{res}}_{l,w} \sim \text{Dirichlet-Multinomial}\Big(h^{\text{res-sum}}_{l,w}, \phi_{l,w}\boldsymbol{\pi}_{l,w}\Big) \tag{S32a}$$
$$\pi_{l,a,w} = \alpha_{l,w}\pi^{\Gamma}_{l,a,w} + (1-\alpha_{l,w})\pi^{\text{non-}\Gamma}_{l,a,w} \tag{S32b}$$
$$\pi^{\Gamma}_{l,a,w} = \text{softmax}\Big(\log\lambda^{\Gamma}_{l,a} + \log n^R_{l,a,w}\Big) \tag{S32c}$$
$$\pi^{\text{non-}\Gamma}_{l,a,w} = \text{softmax}\Big(\log\lambda^{\text{non-}\Gamma}_{l,a} + \log n^R_{l,a,w}\Big) \tag{S32d}$$
$$\log\lambda^{\Gamma}_{l,a} \sim \mathcal{N}(0,1) \tag{S32e}$$
$$\log\lambda^{\text{non-}\Gamma}_{l,a} \sim \mathcal{N}(0,1) \tag{S32f}$$
$$\phi_{l,w} = (h^{\text{res-sum}}_{l,w} - 1)/(\theta_2 + 1) \tag{S32g}$$
$$\theta_2 \sim \text{Exponential}(20), \tag{S32h}$$

where we denote the vector of age-specific hospital admissions in location $l$ and week $w$ by $\boldsymbol{h}^{\text{res}}_{l,w} = \big(h^{\text{res}}_{l,a,w}\big)_{a\in\mathcal{A}}$ and the vector of the age composition of hospital admissions in location $l$ and week $w$ by $\boldsymbol{\pi}_{l,w} = \big(\pi_{l,a,w}\big)_{a\in\mathcal{A}}$, again such that $\sum_a \pi_{l,a,w} = 1$. The Dirichlet-Multinomial is in standard sample size-scale parameterisation such that the means are given by $h^{\text{res-sum}}_{l,w}\pi_{l,a,w}$ as in (S29b), and the scale parameter $\phi_{l,w}$ is conditional on the known $h^{\text{res-sum}}_{l,w}$ re-parameterised into the overdispersion parameter $\theta_2$, with $\theta_2 > 0$. The softmax transformations (S32c-S32d) allow for convenient prior specifications of the log hospital admission rates on the real line, and run over age bands $a$ for fixed location $l$ and fixed week $w$. The prior in (S32h) was chosen to favour the least complex model with no overdispersion.

#### S7.2 Decomposition of COVID-19 attributable in-hospital fatality rates

We model in-hospital fatality rates among hospital admissions in week $w$, location $l$ and age group $a$ that are respectively infected with non-Gamma and Gamma variants with the decomposition

$$\zeta^{\text{non-}\Gamma}_{l,a,w} = \text{logit}^{-1}\Big(\eta^{\text{non-}\Gamma}_{l,a} + X_{l,w}\beta_l\Big) \quad \text{(S33a)}$$

$$\zeta^{\Gamma}_{l,a,w} = \text{logit}^{-1}\Big(\eta^{\text{non-}\Gamma}_{l,a} + \eta^{\Gamma\text{-random-effect}}_{l,a} + X_{l,w}\beta_l\Big), \quad \text{(S33b)}$$

subject to the constraint $\beta_{l,i} \geq 0$ for all $l$ and $i = 1, \ldots, p$. In (S33), $X_{l,w}$ denote a set of $p$ healthcare pressure indices in location $l$ and week $w$ as described in Section S5, and $X_{l,w}\beta_l$ describes time trends in in-hospital fatality rates based on the $p$ healthcare pressure indices regardless of changes in virus variants.

#### S7.3 Estimating factors associated with fluctuating in-hospital fatality rates

To estimate the components of the in-hospital fatality rates that are attributable to each location, to the Gamma variant, and to changes in health-care demand and resource depletion, we note that in the model the proportion of all hospital admissions in week $w$ that occurs in age group $a$ with variant Gamma is $\alpha_{l,w}\pi^{\Gamma}_{l,a,w}$, and similarly for non-Gamma variants, see (S32b). Thus, the proportion of all hospital admissions in age $a$ and week $w$ that in the model is attributed to Gamma is

$$\frac{\alpha_{l,w}\pi^{\Gamma}_{l,a,w}}{\alpha_{l,w}\pi^{\Gamma}_{l,a,w} + (1-\alpha_{l,w})\pi^{\text{non-}\Gamma}_{l,a,w}}.$$

Of this proportion of hospital admissions in age $a$ in week $w$, the proportion of individuals with a fatal outcome is (S33a). Similarly, the proportion of all hospital admissions in age $i$ and week $w$ that is attributed to non-Gamma variants is $\big((1-\alpha_{l,w})\pi^{\text{non-}\Gamma}_{l,a,w}\big)/\big(\alpha_{l,w}\pi^{\Gamma}_{l,a,w} + (1-\alpha_{l,w})\pi^{\text{non-}\Gamma}_{l,a,w}\big)$, and in this proportion individuals face a fatal outcome with probability (S33b). Thus, in age group $a$ and week $w$ the in-hospital fatality rate across variants is given by the weighted average

$$\begin{aligned}\zeta_{l,a,w} = &\frac{(1-\alpha_{l,w})\pi^{\text{non-}\Gamma}_{l,a,w}}{\alpha_{l,w}\pi^{\Gamma}_{l,a,w} + (1-\alpha_{l,w})\pi^{\text{non-}\Gamma}_{l,a,w}}\,\zeta^{\text{non-}\Gamma}_{l,a,w} + \\ &\frac{\alpha_{l,w}\pi^{\Gamma}_{l,a,w}}{\alpha_{l,w}\pi^{\Gamma}_{l,a,w} + (1-\alpha_{l,w})\pi^{\text{non-}\Gamma}_{l,a,w}}\,\zeta^{\Gamma}_{l,a,w}.\end{aligned} \quad \text{(S34)}$$

We fit (S34) to the linked data of hospital admissions among residents of age $a$ in location $l$ and week $w$ and the corresponding, censoring-adjusted COVID-19 attributable deaths among these hospital admissions. This leads us to the model component

$$h^{\text{res-cens-adj-D}}_{l,a,w} \sim \text{Beta-Binomial}\Big(h^{\text{res}}_{l,a,w},\ \zeta_{l,a,w}/\theta_3,\ (1-\zeta_{l,a,w})/\theta_3\Big) \tag{S35a}$$

$$\zeta_{l,a,w} = \frac{(1-\alpha_{l,w})\pi^{\text{non-}\Gamma}_{l,a,w}}{\alpha_{l,w}\pi^{\Gamma}_{l,a,w} + (1-\alpha_{l,w})\pi^{\text{non-}\Gamma}_{l,a,w}}\ \zeta^{\text{non-}\Gamma}_{l,a,w} + \tag{S35b}$$

$$\frac{\alpha_{l,w}\pi^{\Gamma}_{l,a,w}}{\alpha_{l,w}\pi^{\Gamma}_{l,a,w} + (1-\alpha_{l,w})\pi^{\text{non-}\Gamma}_{l,a,w}}\ \zeta^{\Gamma}_{l,a,w} \tag{S35c}$$

$$\text{logit}\ \zeta^{\text{non-}\Gamma}_{l,a,w} = \eta^{\text{non-}\Gamma}_{l,a} + X_{l,w}\beta_l \tag{S35d}$$

$$\text{logit}\ \zeta^{\Gamma}_{l,a,w} = \eta^{\text{non-}\Gamma}_{l,a} + \eta^{\Gamma\text{-random-effect}}_{l,a} + X_{l,w}\beta_l \tag{S35e}$$

$$\eta^{\text{non-}\Gamma}_{l,a} \sim \mathcal{N}(-0.25, 1.5^2) \tag{S35f}$$

$$\eta^{\Gamma\text{-random-effect}}_{l,a} \sim \mathcal{N}(0, \sigma^2_\zeta) \tag{S35g}$$

$$\beta_{l,i} \sim \mathcal{N}_{[0,\infty]}(0, \kappa^2_{l,i}\tau^2) \tag{S35h}$$

$$\kappa_{l,i} \sim \text{Half-Cauchy}(0,1) \tag{S35i}$$

$$\tau \sim \text{Half-Cauchy}(0,0.01) \tag{S35j}$$

$$\sigma_\zeta \sim \text{Exponential}(2) \tag{S35k}$$

$$\theta_3 \sim \text{Exponential}(100), \tag{S35l}$$

where $w = 1,\dots,W_l$, and the age bands $a$ are specified in (S1) in Section S1.3. The Beta-Binomial is in the shape-shape parameterisation with mean $\zeta_{l,a,w}$ and variance equal to the Binomial variance component $h^{\text{res}}_{l,a,w}\zeta_{l,a,w}(1-\zeta_{l,a,w})$ multiplied by $\big(1+(h^{\text{res}}_{l,a,w}-1)\frac{1}{\theta_3^{-1}+1}\big)$ to allow for overdispersion. The priors in (S35f) were chosen to place the non-Gamma in-hospital fatality rate around the empirically observed range. In (S35g), we model the Gamma in-hospital fatality rate as a random effect around the non-Gamma in-hospital fatality rate. The priors in (S35h-S35j) were chosen to favour a priori no dependence of in-hospital fatality rates on hospital load predictors, and shrinkage towards zero using a horseshoe-type shrinkage prior. The prior in (S35l) was chosen to favour the least complex models with no overdispersion.

##### S7.4 Complete model

The complete multi-strain epidemic model consists of the model component described in (S29) to fit against the variant frequency data in location $l$ and week $w$, the model component in (S32) to fit against the age-specific COVID-19 attributable hospital admissions among residents in location $l$ and age band $a$ in week $w$, and the model component in (S35) to fit against the linked, underreporting adjusted, COVID-19 attributable deaths among hospital admissions in residents in location $l$, age band $a$ and week $w$ that had no evidence of vaccination prior to hospital admission.

##### S7.5 Numerical inference

Throughout, the model described through equations (S29-S35) was independently fitted to data from each location using cmdstanr version 0.3.0.9000 [47, 48]. Each inference was conducted in 4 Hamiltonian Monte Carlo

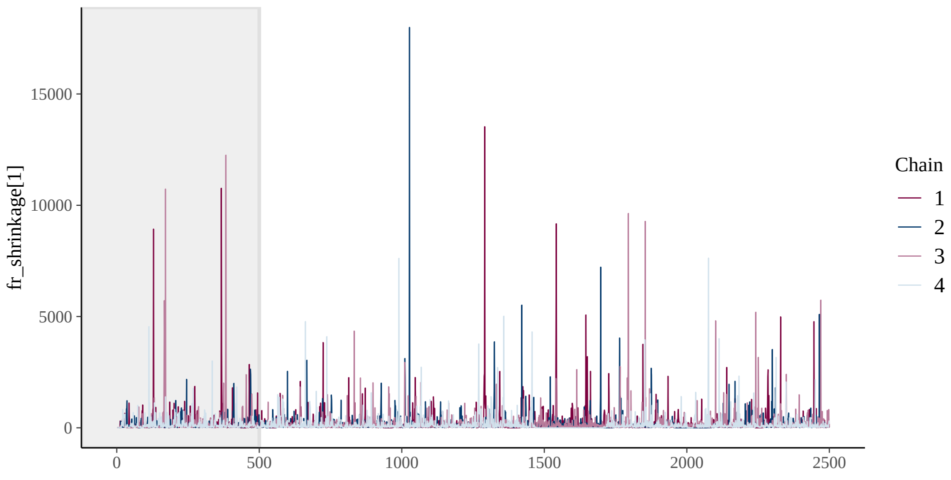

**Figure S29:** Trace plot of the parameter with lowest bulk Effective Sample Size. The minimum value achieved was 210 for the model run for Macapá.

chains, each over 500 warmup iterations, and 2,000 sampling iterations. There were no divergent transitions and the smallest bulk effective sample size was 210 . Figure S29 illustrates the Hamiltonian Monte Carlo chains for the parameter with lowest bulk effective sample size.

#### S7.6 Generated quantities

From the Monte Carlo samples of the joint posterior distribution of the fitted Bayesian multi-strain fatality model, we generate the following quantities.

**Hospital admissions.** We calculate the expected, COVID-19 attributable hospital admissions among residents in location $l$, age band $a$, and week $w$ for non-Gamma and Gamma variants that had no evidence of vaccinat prior to hospitalisation respectively by

$$\begin{aligned} h^{\text{res-non-}\Gamma}_{l,a,w} &= (1-\alpha_{l,w})\pi^{\text{non-}\Gamma}_{l,a,w} h^{\text{res}}_{l,a,w} \\ h^{\text{res-}\Gamma}_{l,a,w} &= \alpha_{l,w}\pi^{\Gamma}_{l,a,w} h^{\text{res}}_{l,a,w}, \end{aligned} \tag{S36}$$

where $h^{\text{res}}_{l,a,w}$ are observed and $\alpha_{l,w}$, $\pi^{\text{non-}\Gamma}_{l,a,w}$, $\pi^{\Gamma}_{l,a,w}$ are from the joint posterior. The expected, COVID-19 attributable hospital admissions among residents in location $l$, age band $a$, and week $w$ across all variants are

$$h^{\text{res-all}}_{l,a,w} = h^{\text{res-non-}\Gamma}_{l,a,w} + h^{\text{res-}\Gamma}_{l,a,w}. \tag{S37}$$

The expected share of age group $a$ among hospital admissions among residents of location $l$ in week $w$ with non-Gamma variants are $\frac{h^{\text{res-non-}\Gamma}_{l,a,w}}{\sum_b h^{\text{res-non-}\Gamma}_{l,b,w}} = \pi^{\text{non-}\Gamma}_{l,a,w}$, and similarly for Gamma. The expected share of age group $a$ among hospital admissions among residents of location $l$ in week $w$ across all variants is $\pi_{l,a,w} = (1-\alpha_{l,w})\pi^{\text{non-}\Gamma}_{l,a,w} +$

$\alpha_{l,w}\pi^{\Gamma}_{l,a,w}$. We calculate the expected ratio in the share of age group $a$ among Gamma hospital admissions among residents of location $l$ versus non-Gamma hospital admissions in the 2020 population by

$$\frac{\pi^{\Gamma}_{l,a,1}}{\pi^{\text{non-}\Gamma}_{l,a,1}} = \frac{\lambda^{\Gamma}_{l,a} n^{R}_{l,a,1}}{\sum_b \lambda^{\Gamma}_{l,b} n^{R}_{l,b,1}} \Big/ \frac{\lambda^{\text{non-}\Gamma}_{l,a} n^{R}_{l,a,1}}{\sum_b \lambda^{\text{non-}\Gamma}_{l,b} n^{R}_{l,b,1}} = \frac{\lambda^{\Gamma}_{l,a} n_{l,a}}{\sum_b \lambda^{\Gamma}_{l,b} n_{l,b}} \Big/ \frac{\lambda^{\text{non-}\Gamma}_{l,a} n_{l,a}}{\sum_b \lambda^{\text{non-}\Gamma}_{l,b} n_{l,b}}, \tag{S38}$$

where $n_{l,a}$ is the 2020 population size in location $l$ in age band $a$ (Section S1.3), and $\lambda^{\text{non-}\Gamma}_{l,b}$, $\lambda^{\Gamma}_{l,b}$ are from the joint posterior. Note that this ratio controls for changes in the populations susceptible to severe COVID-19 over time.

**Deaths following COVID-19 hospital admissions.** We calculate the expected, COVID-19 attributable deaths among hospital admissions in residents in location $l$, age band $a$, and week $w$ for non-Gamma and Gamma variants respectively by

$$\begin{aligned} h^{\text{res-non-}\Gamma\text{-D}}_{l,a,w} &= (1-\alpha_{l,w})\pi^{\text{non-}\Gamma}_{l,a,w} h^{\text{res}}_{l,a,w} \zeta^{\text{non-}\Gamma}_{l,a,w} \\ h^{\text{res-}\Gamma\text{-D}}_{l,a,w} &= \alpha_{l,w}\pi^{\Gamma}_{l,a,w} h^{\text{res}}_{l,a,w} \zeta^{\Gamma}_{l,a,w}, \end{aligned} \tag{S39}$$

where $h^{\text{res}}_{l,a,w}$ are observed and $\alpha_{l,w}$, $\pi^{\text{non-}\Gamma}_{l,a,w}$, $\pi^{\Gamma}_{l,a,w}$, $\zeta^{\text{non-}\Gamma}_{l,a,w}$, $\zeta^{\Gamma}_{l,a,w}$ are from the joint posterior. The expected, COVID-19 attributable deaths among hospital admissions in residents in location $l$, age band $a$, and week $w$ for all variants are

$$h^{\text{res-all-D}}_{l,a,w} = h^{\text{res-non-}\Gamma\text{-D}}_{l,a,w} + h^{\text{res-}\Gamma\text{-D}}_{l,a,w}. \tag{S40}$$

We calculate the expected share of age group $a$ among deaths in hospital admissions among residents of location $l$ in week $w$ with non-Gamma and Gamma variants respectively by

$$\begin{aligned} \frac{h^{\text{res-non-}\Gamma\text{-D}}_{l,a,w}}{\sum_b h^{\text{res-non}\Gamma\text{-D}}_{l,b,w}} &= \frac{\pi^{\text{non-}\Gamma}_{l,a,w}\zeta^{\text{non-}\Gamma}_{l,a,w}}{\sum_b \pi^{\text{non-}\Gamma}_{l,b,w}\zeta^{\text{non-}\Gamma}_{l,b,w}} \\ \frac{h^{\text{res-}\Gamma\text{-D}}_{l,a,w}}{\sum_b h^{\text{res-}\Gamma|text-D}_{l,b,w}} &= \frac{\pi^{\Gamma}_{l,a,w}\zeta^{\Gamma}_{l,a,w}}{\sum_b \pi^{\Gamma}_{l,b,w}\zeta^{\Gamma}_{l,b,w}}, \end{aligned} \tag{S41}$$

where $\pi^{\text{non-}\Gamma}_{l,a,w}$, $\pi^{\Gamma}_{l,a,w}$, $\zeta^{\text{non-}\Gamma}_{l,a,w}$, $\zeta^{\Gamma}_{l,a,w}$ are from the joint posterior. Thus, the expected share of age group $a$ among deaths following hospital admissions in residents in location $l$ in week $w$ regardless of variant is

$$\frac{h^{\text{res-D}}_{l,a,w}}{\sum_b h^{\text{res-D}}_{l,b,w}} = \frac{(1-\alpha_{l,w})\pi^{\text{non-}\Gamma}_{l,a,w}\zeta^{\text{non-}\Gamma}_{l,a,w} + \alpha_{l,w}\pi^{\Gamma}_{l,a,w}\zeta^{\Gamma}_{l,a,w}}{\sum_b (1-\alpha_{l,w})\pi^{\text{non-}\Gamma}_{l,b,w}\zeta^{\text{non-}\Gamma}_{l,b,w} + \alpha_{l,w}\pi^{\Gamma}_{l,b,w}\zeta^{\Gamma}_{l,b,w}}. \tag{S42}$$

We calculate the expected ratio in the share of age group $a$ among Gamma deaths after hospital admission in residents of location $l$ versus non-Gamma deaths in the 2020 population similar to (S38) by

$$\begin{aligned} &\left(\frac{h^{\text{res-}\Gamma\text{-D}}_{l,a,1}}{\sum_b h^{\text{res-}\Gamma\text{-D}}_{l,b,1}}\right) \Big/ \left(\frac{h^{\text{res-non-}\Gamma\text{-D}}_{l,a,1}}{\sum_b h^{\text{res-non-}\Gamma\text{-D}}_{l,b,1}}\right) = \\ &\left(\frac{\lambda^{\Gamma}_{l,a}\zeta^{\Gamma}_{l,a,1} n_{l,a}}{\sum_b \lambda^{\Gamma}_{l,b}\zeta^{\Gamma}_{l,b,1} n_{l,b}}\right) \Big/ \left(\frac{\lambda^{\text{non-}\Gamma}_{l,a}\zeta^{\text{non-}\Gamma}_{l,a,1} n_{l,a}}{\sum_b \lambda^{\text{non-}\Gamma}_{l,b}\zeta^{\text{non-}\Gamma}_{l,b,1} n_{l,b}}\right), \end{aligned} \tag{S43}$$

where $n_{l,a}$ is the 2020 population size in location $l$ in age band $a$ (Section S1.3), and $\lambda^{\text{non-}\Gamma}_{l,b}$, $\lambda^{\Gamma}_{l,b}$, $\zeta^{\text{non-}\Gamma}_{l,a,w}$, $\zeta^{\Gamma}_{l,a,w}$ are from the joint posterior. Note that this ratio controls for changes in the populations susceptible to severe COVID-19 over time, and for changes in hospital pressures over time.

**Contribution of location to COVID-19 in-hospital fatality rates.** To compare COVID-19 in-hospital fatality rates across locations, we define the overall in-hospital fatality rate in location $l$ and week $w$ in an age-standardised population that adjust for differences in age composition across locations. Specifically, we calculate

$$\zeta^{\text{age-std}}_{l,w} = \sum_a \frac{n^{\text{cities}}_a}{\sum_b n^{\text{cities}}_b} \zeta_{l,a,w} \tag{S44}$$

where $n^{\text{cities}}_a$ is the population size in age band $a$ across all cities considered, and $\zeta_{l,a,w}$ is from the joint posterior. Then, for each location, we define the week $w^\star_l$ with lowest in-hospital fatality rate as the week that minimises the posterior median of (S44),

$$w^\star_l = \text{argmin}_{w\in 1:W^{\text{detect-}\Gamma}_l} \big(\text{posterior median of } \zeta^{\text{age-std}}_{l,w}\big), \tag{S45}$$

and calculate the lowest, age standardised COVID-19 in-hospital fatality rates prior to Gamma's first detection through

$$\zeta^{\text{lowest}}_l = \zeta^{\text{age-std}}_{l,w^\star_l}. \tag{S46}$$

It is helpful to note that the healthcare pressure indices are standardised and evaluate to zero in each location's reference week, i. e. the week prior to Gamma's first detection with lowest empirical, age-standardised in-hospital fatality rate as described in Section S5. The week $w^\star_l$ typically corresponds to the week with lowest empirical, age-standardised in-hospital fatality rate, and so $\zeta^{\text{lowest}}_l$ is evaluated when the healthcare pressure effect $X_{l,w}\beta_l$ is zero. The estimated Gamma frequencies $\alpha_{l,w}$ are of course also very small prior to Gamma's first detection, and so the lowest, age standardised COVID-19 in-hospital fatality rates can be more simply expressed with

$$\zeta^{\text{lowest}}_l \approx \sum_a \frac{n^{\text{cities}}_a}{\sum_b n^{\text{cities}}_b} \text{logit}^{-1}\big(\eta^{\text{non-}\Gamma}_{l,a}\big), \tag{S47}$$

where $\eta^{\text{non-}\Gamma}_{l,a}$ are the intercept terms in our regression (S33) and from the joint posterior. Then, to compare (S46) across locations, we find the location with overall lowest age-standardised in-hospital fatality rate by

$$l^\star = \text{argmin}_l\, \zeta^{\text{lowest}}_l, \tag{S48}$$

and compute for all other locations $l$ the ratio

$$\zeta^{\text{lowest-ratio}}_l = \zeta^{\text{lowest}}_l / \zeta^{\text{lowest}}_{l^\star}. \tag{S49}$$

We interpret (S49) as the location effect on COVID-19 in-hospital fatality rates. Because of (S47), the location effect does not include contributions attributable to the healthcare pressure indices ($X_{l,w}\beta_l$), nor any contributions attributable to the non-parametric Gamma effects ($\eta^{\Gamma}_{l,a}$) on in-hospital fatality rates.

**Contribution of healthcare pressure to COVID-19 in-hospital fatality rates.** To compare time trends to in-hospital fatality rates in each location, we calculate for each location the multiplicative effect of changes in healthcare demand and resources in week $w$ in location $l$ by

$$\zeta^{\text{multiplier}}_{l,w} = \sum_a \frac{n^{\text{cities}}_a}{\sum_b n^{\text{cities}}_b} \big(\zeta^{\text{non-}\Gamma}_{l,a,w} / \zeta^{\text{non-}\Gamma}_{l,a,w^\star_l}\big), \tag{S50}$$

where $\zeta_{l,a,w}^{\text{non-}\Gamma}$ are from the joint posterior. It is helpful to recall Equation (S33a), which shows that we have more simply

$$\zeta_{l,w}^{\text{multiplier}} \approx \sum_a \frac{n_a^{\text{cities}}}{\sum_b n_b^{\text{cities}}} \frac{\text{logit}^{-1}\big(\eta_{l,a}^{\text{non-}\Gamma} + X_{l,w}\beta_l\big)}{\text{logit}^{-1}\big(\eta_{l,a}^{\text{non-}\Gamma}\big)}. \tag{S51}$$

Here, the approximation is because the Gamma frequencies $\alpha_{l,w}$ are very small but not exactly zero, and because the week in which the loess-smoothed fatality rates are lowest may not exactly coincide with the week $w_l^\star$ in which the model-based fatality rates are lowest. We interpret (S50) as the healthcare pressure effect on COVID-19 in-hospital fatality rates. Because of (S51), the healthcare pressure effect does not include contributions attributable to the non-parametric Gamma effects ($\eta_{l,a}^{\Gamma}$) on in-hospital fatality rates, and corresponds to the multiplier to the minimum fatality rates in each location that is associated with the healthcare pressure indices ($X_{l,w}\beta_l$).

**Contribution of Gamma to COVID-19 in-hospital fatality rates.** Finally we identify the effect of Gamma on in-hospital fatality rates as measured through population-level increases in Gamma's variant frequency in each location. Standardising across age bands, we calculate the ratio in Gamma versus non-Gamma in-hospital fatality rates in location $l$ by

$$\zeta_l^{\Gamma-\text{ratio}} = \sum_a \frac{n_a^{\text{cities}}}{\sum_b n_b^{\text{cities}}} \zeta_{l,a,w_l^\star}^{\Gamma} / \zeta_{l,a,w_l^\star}^{\text{non-}\Gamma}, \tag{S52}$$

where $\zeta_{l,a,w_l^\star}^{\Gamma}$, $\zeta_{l,a,w_l^\star}^{\text{non-}\Gamma}$ are from the joint posterior. We interpret (S52) as the Gamma effect on in-hospital fatality rates. It is again helpful to recall Equation (S33), which shows that we can approximate the Gamma effect through

$$\zeta_l^{\Gamma-\text{ratio}} \approx \sum_a \frac{n_a^{\text{cities}}}{\sum_b n_b^{\text{cities}}} \frac{\text{logit}^{-1}\big(\eta_{l,a}^{\text{non-}\Gamma} + \eta_{l,a}^{\Gamma}\big)}{\text{logit}^{-1}\big(\eta_{l,a}^{\text{non-}\Gamma}\big)}. \tag{S53}$$

This shows that $\zeta_l^{\Gamma-\text{ratio}}$ does not include contributions attributable to the healthcare pressure indices ($X_{l,w}\beta_l$), and corresponds to the multiplier to the minimum fatality rates in each location that is associated with the non-parametric Gamma effects ($\eta_{l,a}^{\text{non-}\Gamma}$).

### S8 Counterfactual analyses

To quantify the impact that the observed fluctuations in COVID-19 attributable in-hospital rates had on the death toll in the 14 state capitals, we performed two counterfactual analyses. The aim of the two counterfactuals was to estimate how many COVID-19 attributable deaths could have been avoided with sufficient healthcare resources so that healthcare pressures would not result in shocks in in-hospital fatality rates (Scenario 1), or so that healthcare pressures would not result in shocks in in-hospital fatality rates and in addition there would have been the same low baseline demand fatality rate across locations (Scenario 2). The first counterfactual is thus based on the minimum in-hospital fatality rates that were observed in each location, and which are thus achievable in each location. The second counterfactual in addition assumes that the lowest observed in-hospital fatality rate, which we observed in Belo Horizonte, could be achieved in all other locations. This section provides technical details on the implementation of these counterfactuals.

##### S8.1 Counterfactual 1

For each location $l$, we consider the estimated age-standardised in-hospital fatality rate (Equation (S44)) in the reference week, i. e. the week with the lowest age-standardised in-hospital fatality rate prior to Gamma's emergence in each location. In this counterfactual, we then compute the cumulated COVID-19 attributable deaths following hospital admissions among residents in location $l$ that had no evidence of vaccination prior to hospital admission, for each age band $a$ and across age bands as

$$\begin{aligned} h^{\text{hyp1-res-D}}_{l,a} &= \sum_{w=1}^{W_l} h^{\text{res}}_{l,a,w} \zeta_{l,a,w^\star_l} \\ h^{\text{hyp1-res-D}}_{l} &= \sum_{a\in\mathcal{A}} h^{\text{hyp1-res-D}}_{l,a}, \end{aligned} \tag{S54}$$

where the week indices range over the entire observation period up to the final week $W_l$ in each location. We define the expected COVID-19 attributable deaths that could have been avoided in the absence of healthcare pressures in location $l$ during the observation period relative to this hypothetical scenario by

$$h^{\text{hyp1-avoidable-D}}_{l} = \Big(\sum_{w=1}^{W_l}\sum_{a\in\mathcal{A}} h^{\text{res-adj-D}}_{l,a,w}\Big) - h^{\text{hyp1-res-D}}_{l}, \tag{S55}$$

where the observed $h^{\text{res-adj-D}}_{l,a,w}$ are described in Section S3. Similarly, we define the expected percentage reduction in COVID-19 attributable deaths relative to this hypothetical scenario by

$$1 - p^{\text{hyp1-avoidable-D}}_{l} = \frac{h^{\text{hyp1-avoidable-D}}_{l}}{\sum_{w=1}^{W_l}\sum_{a\in\mathcal{A}} h^{\text{res-adj-D}}_{l,a,w}}. \tag{S56}$$

##### S8.2 Counterfactual 2

In the second counterfactual analysis, we considered the location $l^{\star\star}$ in which we found the lowest age-standardised in-hospital fatality rate prior to Gamma's detection in each location,

$$l^{\star\star} = \text{argmin}_l\Big(\min_{w\in 1:W^{\text{detect-}\Gamma}_l}\big(\text{posterior median of } \xi^{\text{age-std}}_{l,w}\big)\Big). \tag{S57}$$

In this counterfactual, we then compute the total, cumulated COVID-19 attributable deaths following hospital admissions among residents in location $l$ that had no evidence of vaccination prior to hospital admission, for each age band $a$ and across age bands as

$$\begin{aligned} h^{\text{hyp2-res-D}}_{l,a} &= \sum_{w=1}^{W_l} h^{\text{res}}_{l,a,w} \zeta_{l^{\star\star},a,w^\star} \\ h^{\text{hyp2-res-D}}_{l} &= \sum_{a\in\mathcal{A}} h^{\text{hyp2-res-D}}_{l,a}, \end{aligned} \tag{S58}$$

where again the week indices range over the entire observation period up to the final week $W_l$ in each location. In analogy to the first counterfactual, we then calculate

$$h_l^{\text{hyp2-avoidable-D}} = \Big( \sum_{w=1}^{W_l} \sum_{a \in \mathcal{A}} h_{l,a,w}^{\text{res-adj-D}} \Big) - h_l^{\text{hyp2-res-D}}, \tag{S59a}$$

$$p_l^{\text{hyp2-avoidable-D}} = 1 - \frac{h_l^{\text{hyp2-res-D}}}{\sum_{w=1}^{W_l} \sum_{a \in \mathcal{A}} h_{l,a,w}^{\text{res-adj-D}}}. \tag{S59b}$$

### S9 Sensitivity analyses

#### S9.1 Using SARS-CoV-2 genomic sequence data obtained under controlled sampling

This section describes key study outcomes when in place of the SARS-CoV-2 sequence data from GISAID we instead used SARS-CoV-2 sequence data that was obtained under more controlled sequence sampling frames. We performed this sensitivity analysis because SARS-CoV-2 sequences are often obtained from patients with unusual clinical progression [49], and therefore may not be representative of Gamma's temporal expansion at population level.

Data were available from three locations, Manaus, Belo Horizonte, and São Paulo, and are shown in Supplementary Table S7. In Manaus, samples from PCR-positive residents testing in two private laboratories through nasal and oropharyngeal swabs were selected at random regardless of Ct values for sequencing. The samples were sequenced and processed using the ARTIC bioinformatics pipeline [50] as described in [51]. Viral genomes recovered from $147$ samples collected between November 1, 2020 and January 10, 2021 had sufficient genome coverage enabling lineage classification with pangolin version 2.2.1 [52, 53]. In Belo Horizonte, samples were selected at random from PCR-positive residents in three laboratories (Laboratório Hermes Pardini, Laboratório de Biologia Integrativa, UFMG, and Laboratório Municipal de Referência, PBH). The samples were sequenced on the Illumina MiSeq platform and processed using a custom pipeline. Idenfitied mutations were manually inspected, and the sequences were classified using Pangolin version 2.2.1 [52]. In total, $27$ samples were classified as P.1/Gamma, and $47$ samples to other lineages. For São Paulo, sequences were generated by the Adolf Lutz Institute (IAL), a national public health and reference laboratory for São Paulo State, Brazil, retrieved from GISAID (https://www.gisaid.org) for the period November 1, 2020, to March 31, 2021. In total, $76$ sequences were analysed between November 1, 2020, and March 31, 2021.

| Week | Manaus | | Belo Horizonte | | São Paulo | |
|---|---|---|---|---|---|---|
| | Genomes sampled | Gamma positive | Genomes sampled | Gamma positive | Genomes sampled | Gamma positive |
| 27/05/20 | 2 | 0 | - | - | - | - |
| 02/11/20 | 9 | 0 | - | - | 1 | 0 |
| 09/11/20 | 2 | 0 | - | - | - | - |
| 16/11/20 | 4 | 0 | - | - | - | - |
| 23/11/20 | - | - | - | - | 1 | 0 |
| 30/11/20 | 2 | 1 | - | - | - | - |
| 07/12/20 | 7 | 1 | - | - | - | - |
| 14/12/20 | 24 | 7 | - | - | - | - |
| 21/12/20 | 37 | 20 | - | - | 4 | 0 |
| 28/12/20 | 14 | 8 | - | - | 2 | 0 |
| 04/01/21 | 46 | 40 | 2 | 0 | 3 | 1 |
| 11/01/21 | - | - | 1 | 0 | 7 | 5 |
| 18/01/21 | - | - | - | - | 7 | 2 |
| 25/01/21 | - | - | 2 | 0 | 3 | 1 |
| 01/02/21 | - | - | 3 | 0 | 1 | 0 |
| 08/02/21 | - | - | 1 | 0 | 2 | 1 |
| 15/02/21 | - | - | 3 | 0 | 14 | 10 |
| 22/02/21 | - | - | 1 | 0 | 10 | 9 |
| 01/03/21 | - | - | 37 | 11 | 5 | 5 |
| 08/03/21 | - | - | 24 | 16 | 6 | 6 |
| 15/03/21 | - | - | - | - | 2 | 2 |
| 22/03/21 | - | - | - | - | 7 | 6 |
| 29/03/21 | - | - | - | - | 1 | 1 |

**Table S7:** SARS-CoV-2 sequence data from Manaus, Belo Horizonte, and São Paulo, obtained under controlled sequence sampling frames.

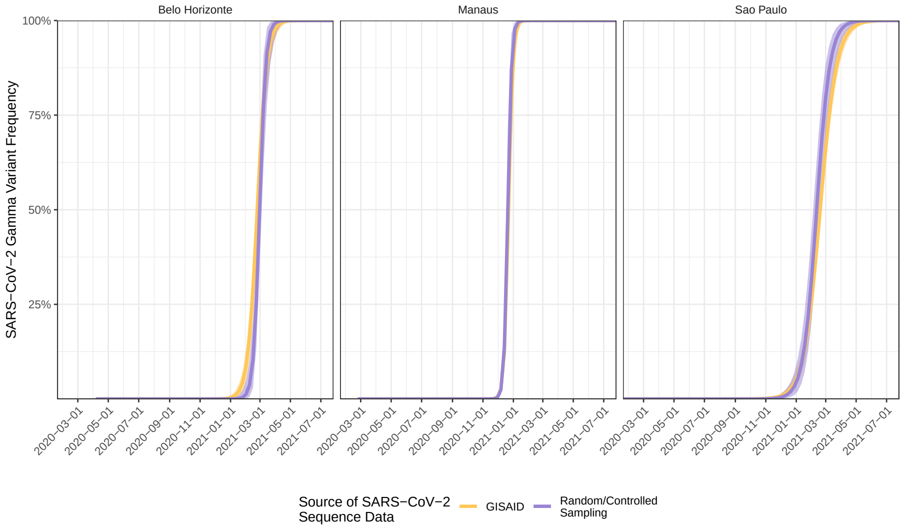

**Figure S30:** Estimated dynamics of Gamma's temporal expansion in Belo Horizonte, Manaus, and São Paulo, when using sequence data collected using controlled sampling methodologies. Posterior median estimates of Gamma's variant frequency (dots) are shown along with 95% credible intervals (linerange) for the central analysis (yellow) and sensitivity analysis (purple).

Supplementary Figure S30 compares Gamma's estimated replacement dynamics under the Bayesian multi-strain fatality model when using sequence data obtained under a random or controlled sampling frame compared to when using the GISAID data. The figure illustrates that Gamma's estimated replacement dynamics in the three cities are insensitive to the differences in the two sequence data sets, and indeed all other aspects of model fit showed similar levels of congruence (not shown). Supplementary Figure S31 compares the key study outcomes, the estimated location, Gamma, and healthcare pressure effect sizes, when using the sequence data obtained under a random or controlled sampling frame compared to when using the GISAID data. All estimated effect sizes were highly concordant, highlighting the robustness of the inferred conclusions using GISAID data, which are available for a wider range of locations.

#### S9.2 Using genomic sequence data obtained from Rede Genomica Fiocruz

This section describes key study outcomes when in place of the SARS-CoV-2 sequence data from GISAID we instead used sequence data publicly available via Rede Genomica Fiocruz. This sensitivity analysis was carried out across all 14 cities, and a detailed description of the data and its associated sampling methodologies is available at http://www.genomahcov.fiocruz.br. We selected only genome sequences obtained between November 2020 and May 2021, in-keeping with our approach to collating GISAID sequence data. This information was collected from data updated on June 15, 2021. We again assumed that the variant frequencies reported at state level are representative of the variant frequencies in state capitals. The raw counts for sampled genomes over time are presented in Supplementary Figure S32.

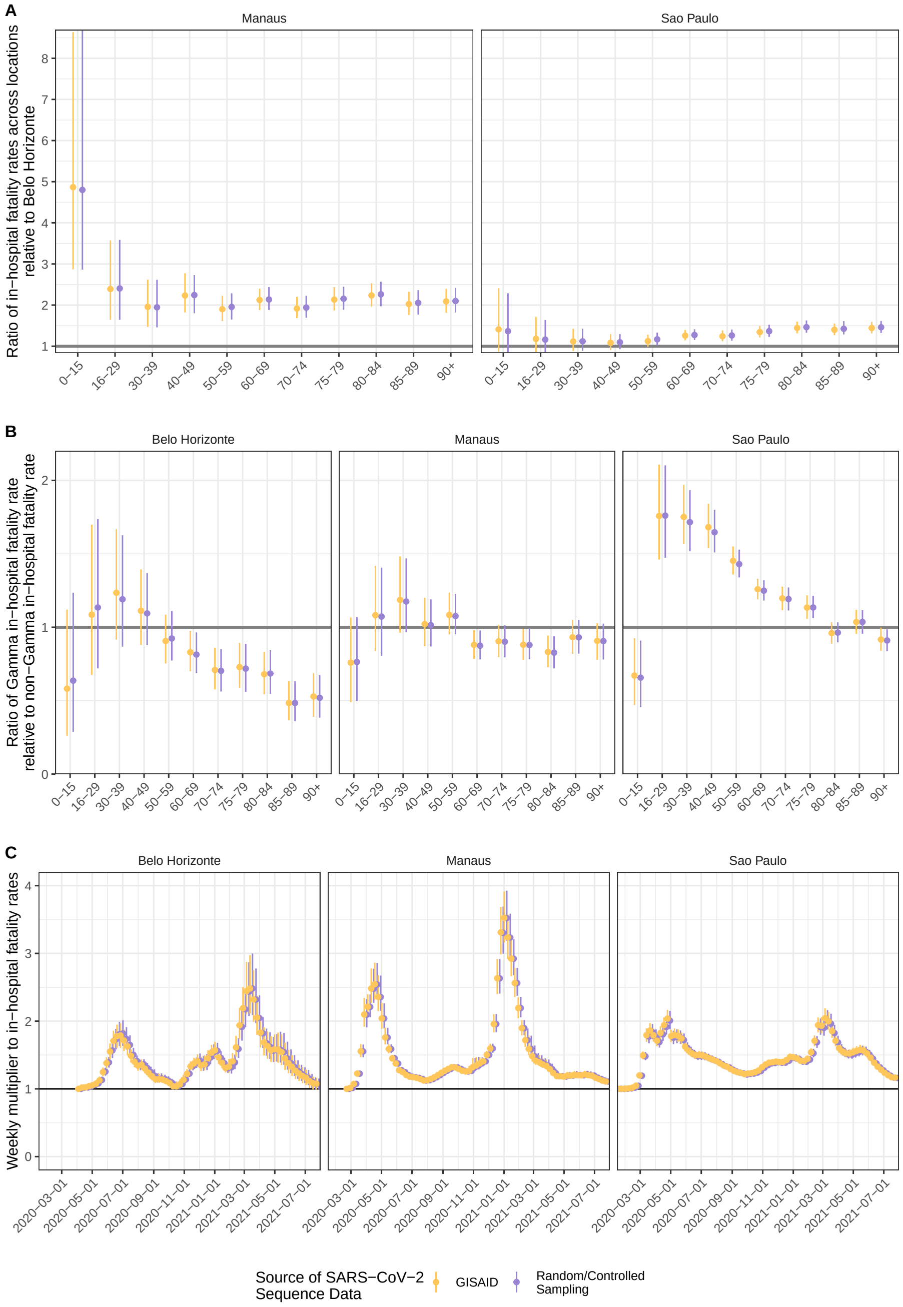

**Figure S31:** Estimated location, Gamma, and healthcare pressure effects when using sequence data collected using controlled sampling methodologies. (A) Estimated location effect, defined as in the main text and shown in Figure 5B. Posterior median estimates (dots) are shown along with 95% credible intervals (linerange) for the central analysis (yellow) and sensitivity analysis (purple). (B) Estimated Gamma effect, defined as in the main text and shown in Figure 5C. (C) Estimated healthcare pressure effect, defined as in the main text and shown in Figure 5D.

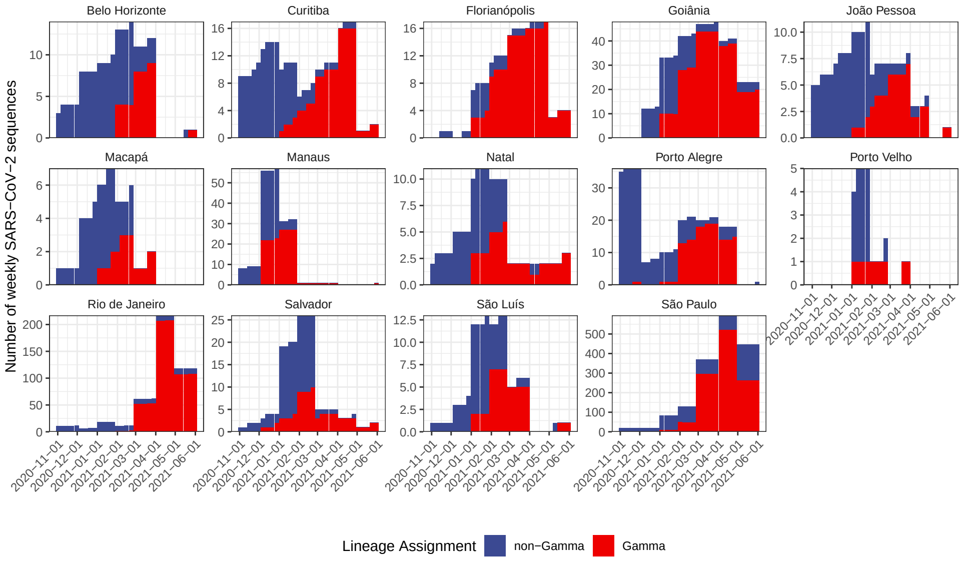

**Figure S32:** SARS-CoV-2 sequence data obtained from Rede Genomica Fiocruz in the 14 states in which the 14 state capitals are located. Data are shown by state capital, assuming that the variant frequencies reported at state level are representative of the variant frequencies in state capitals.

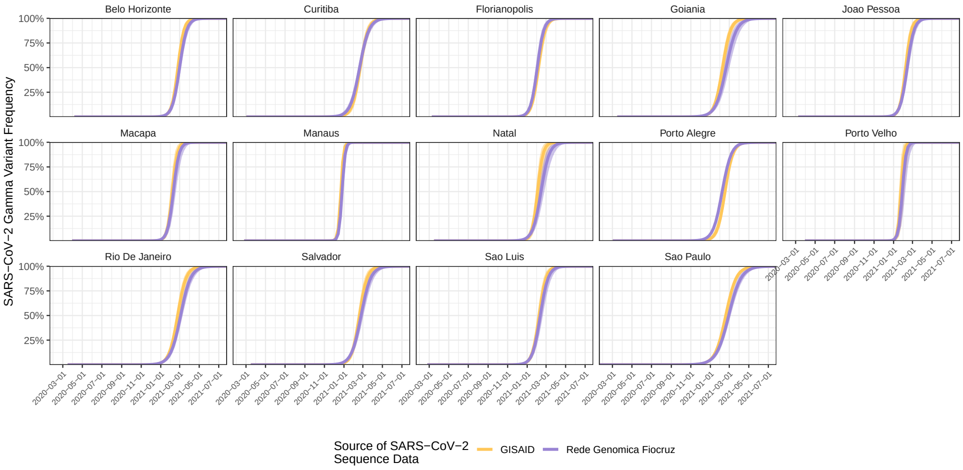

**Figure S33:** Estimated temporal dynamics of Gamma frequency across the 14 cities when using sequence data from Rede Genomica Fiocruz. Posterior median estimates (dots) are shown along with 95% credible intervals (linerange) for the central analysis (yellow) and sensitivity analysis (purple).

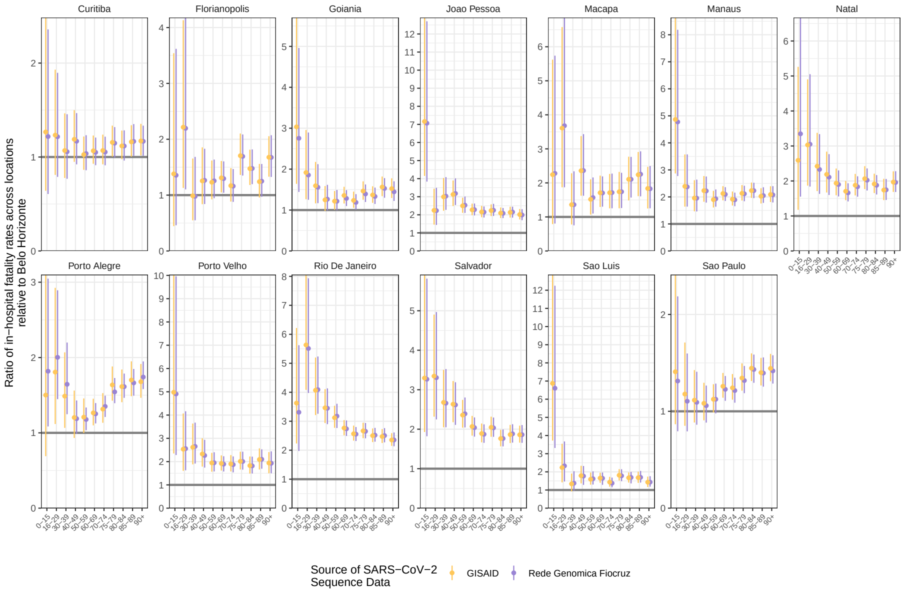

**Figure S34:** Estimated effect of location on in-hospital fatality rates when using sequence data from Rede Genomica Fiocruz. Posterior median estimates (dots) are shown along with 95% credible intervals (linerange) for the central analysis (yellow) and sensitivity analysis (purple).

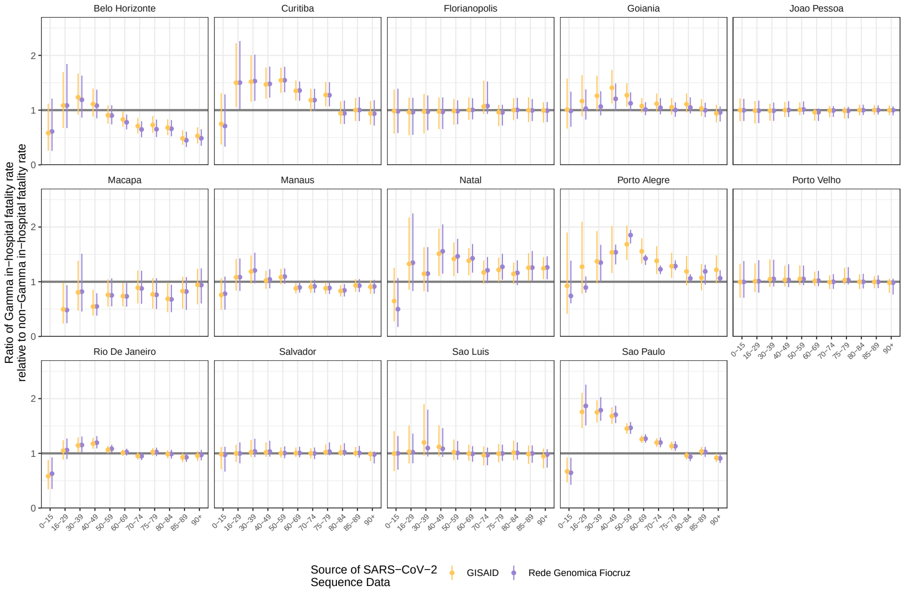

**Figure S35:** Estimated effect of Gamma infection on in-hospital fatality rates when using sequence data from Rede Genomica Fiocruz. Posterior median estimates (dots) are shown along with 95% credible intervals (linerange) for the central analysis (yellow) and sensitivity analysis (purple).

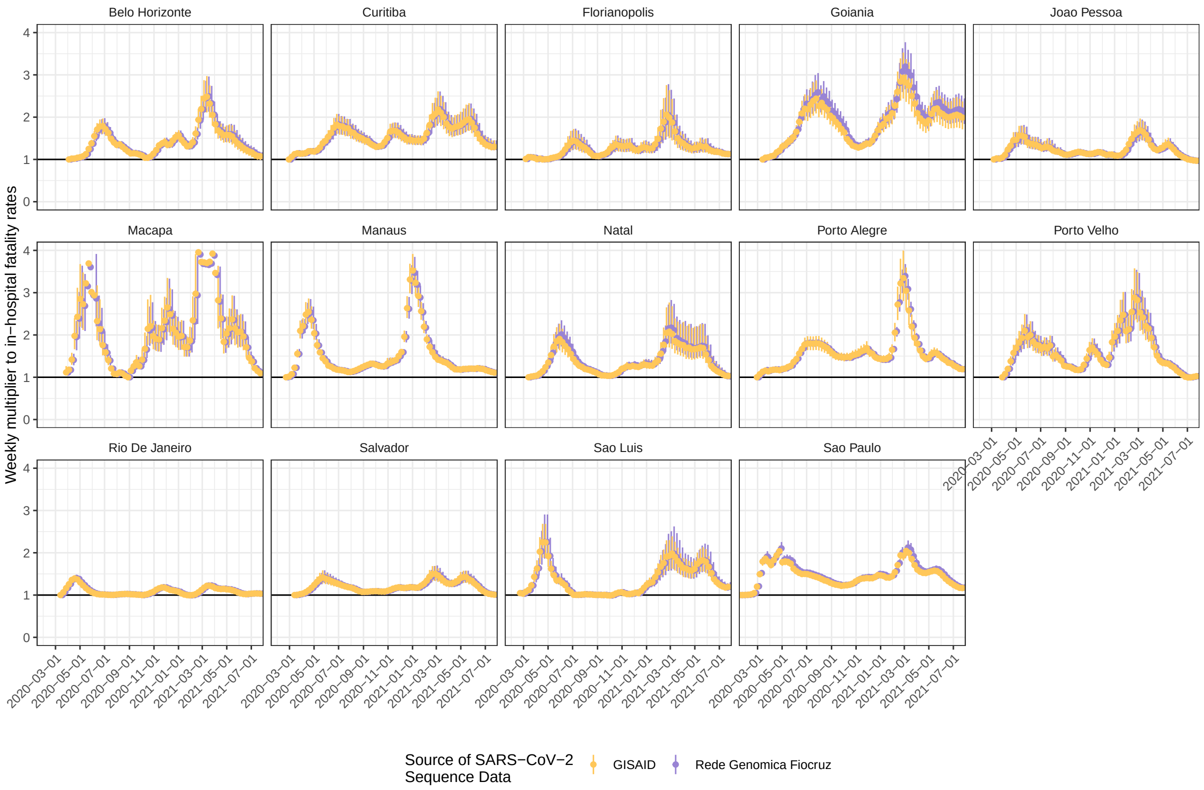

**Figure S36:** Estimated effect of healthcare pressure on in-hospital fatality rates when using sequence data from Rede Genomica Fiocruz. Posterior median estimates (dots) are shown along with 95% credible intervals (linerange) for the central analysis (yellow) and sensitivity analysis (purple).

Supplementary Figure S33 compares the modelled replacement dynamics of Gamma (as inferred by the Bayesian multi-strain fatality model) when using genomic data either from GISAID or from Rede Genomica Fiocruz. The inferred temporal dynamics of Gamma frequency across all 14 locations show only minor differences depending on the dataset utilised. This close concordance was also observed for the model-estimated effects of location (Supplementary Figure S34), infection with the Gamma variant (Supplementary Figure S35), and healthcare pressure (Supplementary Figure S36) on in-hospital fatality rates.

#### S9.3 Patients and resources in private hospitals

This section describes fluctuations in COVID-19 in-hospital fatality rates when the data were restricted to patients and resources in private hospitals. We performed this sensitivity analysis to investigate the extent to which fluctuations in COVID-19 in-hospital fatality rates were also observed in private hospital settings.

For São Paulo, we were able to classify hospitals based on their unique hospital identifier number reported in the SIVEP-Gripe [1] almost completely into “public” or “private” hospitals, or, where an assignment could not be

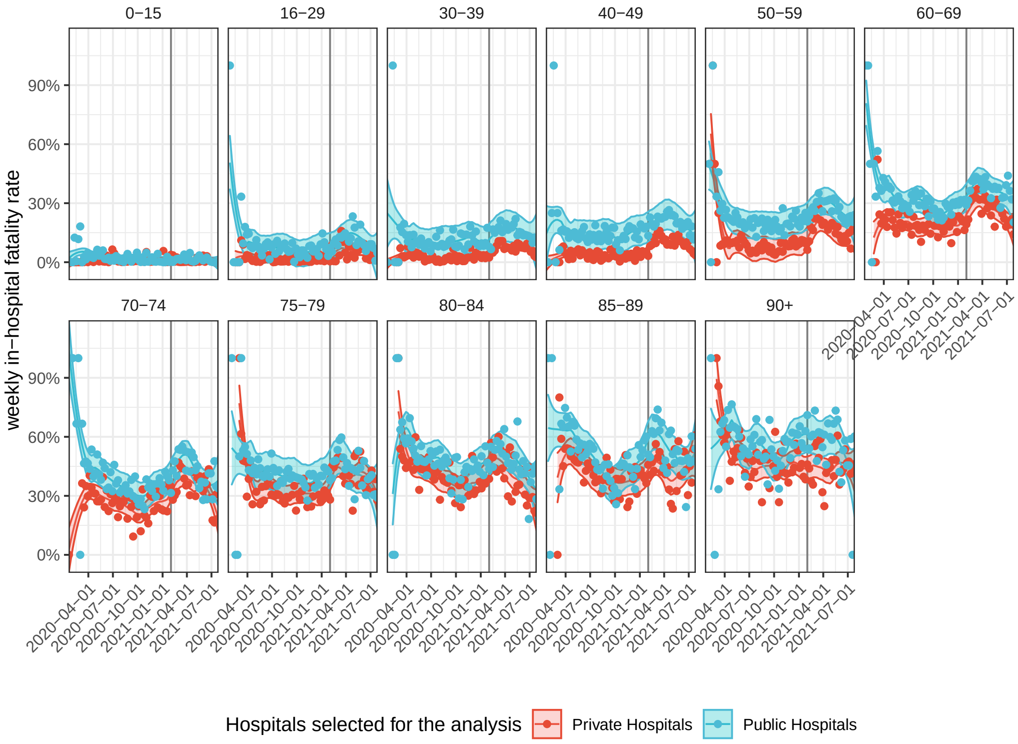

**Figure S37:** Non-parametric estimates of COVID-19 in-hospital fatality rates in public and private hospitals by age group in São Paulo. Nearly all of São Paulo's hospitals could be categorised into public or private healthcare facilities, and COVID-19 in-hospital fatality rates were calculated among unvaccinated patients in public hospitals (blue), and private hospitals (red). Non-parametric, smoothed loess estimates (line) are shown with 95% confidence intervals (ribbon). Weekly data are shown as dots, and the date of Gamma's first detection is indicated as a vertical dotted black line.

made based on available information, into "public or private" hospitals. The data are shown in Supplementary Figure S3.

We then calculated the non-parametric estimates of COVID-19 in-hospital fatality rates in each age group as described in Section S4 among unvaccinated, resident patients that were hospitalised in either private or public healthcare facilities in São Paulo. Figure S37 compares the COVID-19 in-hospital fatality rates in public versus private healthcare facilities in São Paulo for each age group. Overall, we find that COVID-19 in-hospital fatality rates were over time consistently and significantly lower in São Paulo's private healthcare facilities compared to São Paulo's public healthcare facilities. However, importantly, we found similar, synchronised fluctuations in private and public healthcare facilities over time. This suggests that the large impact of pandemic resource limitations on fatality rates that we characterise are common.

#### S9.4 Excluding patients without data on vaccination status

In Belo Horizonte, the age-standardised COVID-19 in-hospital fatality rates declined over the summer months of 2021 to levels well below those seen during earlier time periods (Figure 5A in the main text). We hypothesised that larger numbers of patients hospitalised in Belo Horizonte may have been vaccinated, with no vaccination record reported to SIVEP-Gripe. To address this possibility, we performed a sub-analysis in which we excluded patients with unknown vaccination status in the SIVEP-Gripe data set, and the results of this sensitivity analysis are reported in this Section.

From the denominator of COVID-19 attributable hospitalised residents $h^{\text{res}}_{l,a,w}$ described in Section S1.6, we further excluded patients with unknown vaccination status on or after March 29, 2021. We restricted exclusion of patients with unknown vaccination status to the time period on or after March 29, 2021 because all records prior to 2021 have unknown vaccination status (see Figure S38), and we were unable to generate robust smoothed trends with data in January and March excluded. We then re-calculated the non-parametric and age-standardised estimates of COVID-19 in-hospital fatality rates (Section S4). Figure S38 compares the in-hospital fatality rates in reported unvaccinated patients or patients with unknown vaccination status (blue) to those in reported unvaccinated patients only (red). For most state capitals, hospitalised patient denominators were too small to obtain reliable estimates of in-hospital fatality rates. In the remaining locations, our analysis suggests the estimated COVID-19 in-hospital fatality rates are not significantly different when patients with unknown vaccination status are excluded, and thus we expect that missing data on vaccination status has likely no substantial impact on our primary findings. However we note that Belo Horizonte remained an exception, with larger discrepancies obsserved, and so the COVID-19 in-hospital fatality rates reported in the main text could be confounded with unreported vaccination status.

#### S9.5 Alternative assumptions on COVID-19 attributable patients with unknown clinical outcome

The highest age-standardised COVID-19 in-hospital fatality rates were observed in Rio de Janeiro, against the national trend of declining rates in Brazil's South and Southeast macroregions. This prompted us to compare excess deaths derived from Brazilian's Civil Registry to the COVID- 19 attributable in hospital deaths (Section S1.9), which suggested that a smaller proportion of hospitalised patients with unknown clinical outcomes may have died, compared to what we expect based on our under-reporting adjustments (Section S3). In a sensitivity analyses, we assumed that all patients with unknown clinical outcomes survived, and the results of this analysis are reported in this section.

All analyses were performed a second time, with the only change being that all patients with unknown clinical outcomes survived. Figure S39 shows the resulting age-standardised COVID-19 in-hospital fatality rates, which are directly comparable to those in Figure 5A in the main text. We find that Rio de Janeiro's COVID-19 in-hospital fatality rates were also the highest among all 14 state capitals under this alternative assumption.

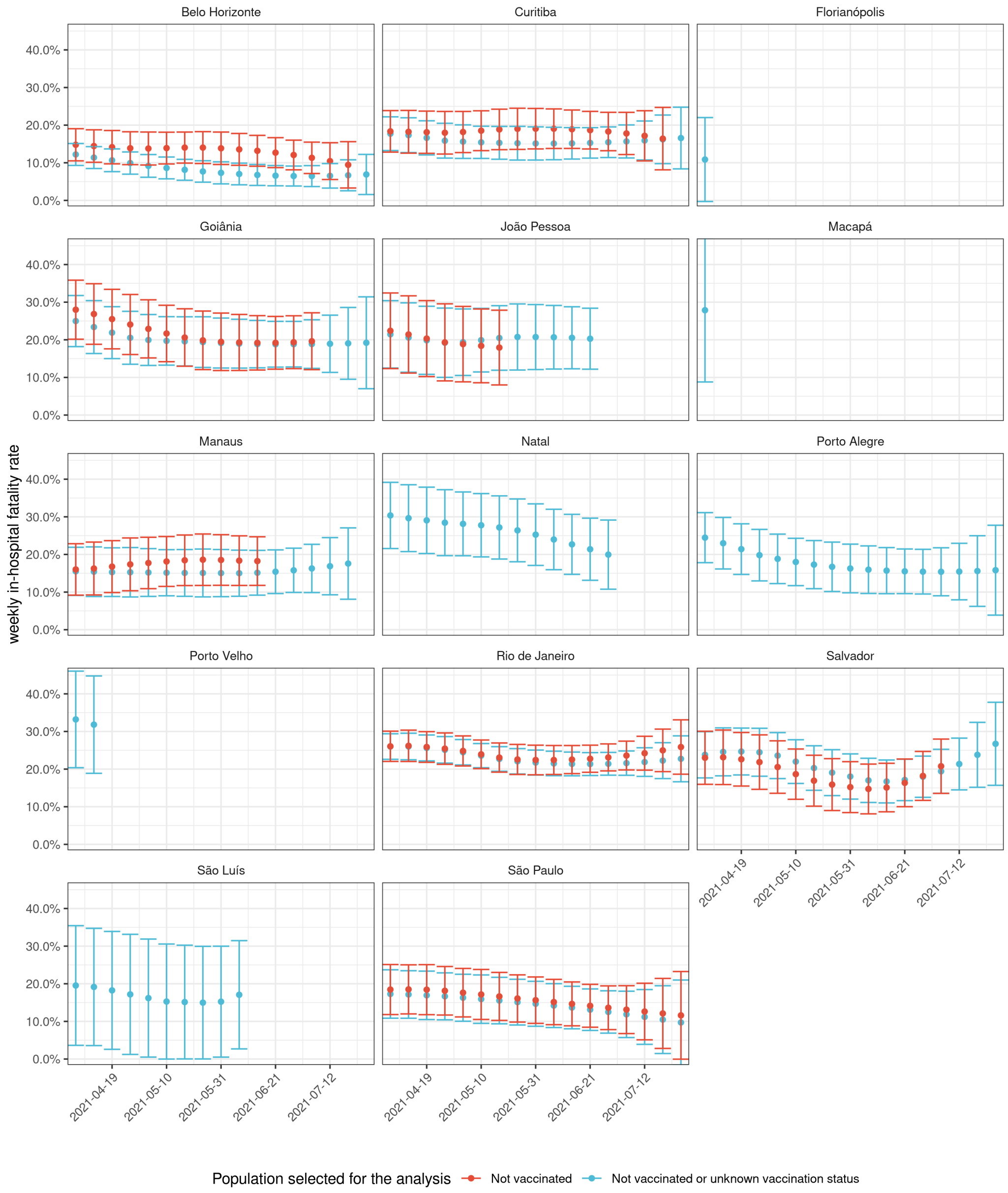

**Figure S38:** Age-standardised COVID-19 fatality rates in reported unvaccinated patients only. From the denominator of COVID-19 attributable hospitalised residents, we further excluded patients with unknown vaccination status on or after March 29, 2021, and re-calculated the non-parametric, smoothed estimates of age-standardised COVID-19 fatality rates. To avoid extrapolation, we only included estimates obtained prior to the last date for which there was at least one hospital admission per age group. Midpoint estimates (dots) are shown along with 95% confidence intervals (errorbars) for two patient groups depending on reported vaccination status (colour).

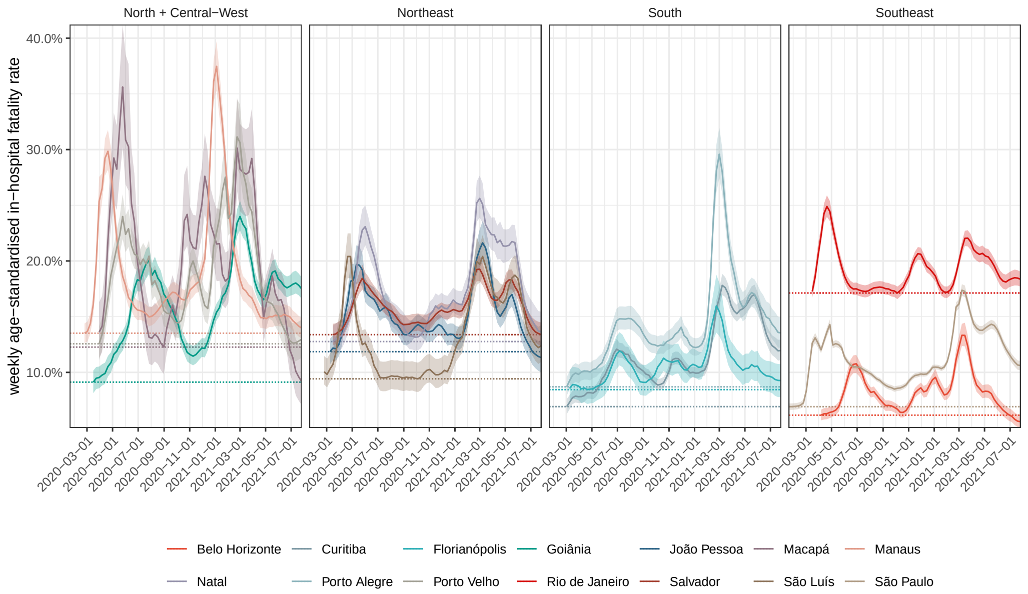

**Figure S39:** Estimated, weekly age-standardised COVID-19 in-hospital fatality rates, averaged across SARS-CoV-2 variants under the assumption that hospitalisations with unknown outcome were discharged. Posterior median estimates (line) are shown with 95%CrIs (ribbon), and the lowest estimated fatality rates during the observation period in each state capital (dotted horizontal line).

#### S9.6 Indicators of disease severity in COVID-19 attributable hospitalised patients

In-hospital fatality rates also depend on who, and under what circumstances, severely ill patients are admitted to hospitals. This prompted us to investigate if the observed fluctuations in COVID-19 fatality rates could in part be the result of concomitant changes in the profile of admitted patients. There is limited data on disease severity at time of hospital admission available in SIVEP-Gripe, however one indicator that can be readily calculated is the time between hospital admission and death in patients with a fatal outcome.

Figure S40 shows the weekly proportion of COVID-19 attributable hospitalisation in residents and non-residents without evidence of vaccination prior to hospitalisation and with a fatal outcome, by time to death. We stratified time to death by number of weeks, shown in fill colours. In Macapá, Manaus, Porto Alegre, and Porto Velho, we find substantially shorter times to death when the number of COVID-19 attributable hospital admissions peaked. This suggests that admitted patients may already have been at a more severe clinical stage when admitted, or that during times of peak demand healthcare pressure in hospitals both increased fatality rates and led to faster progression to death. Further data on out-of-hospital deaths shows that COVID-19 attributable out-of-hospital deaths typically occurred during times of peak demand, with the exception of Rio de Janeiro (Supplementary Figure S3). Together, these data suggest that increased healthcare pressure likely acts to shape in-hospital fatality rates through distinct mechanisms, through a combination of both a reduced ability to provide adequate care and an increase in the average severity of admitted patients.

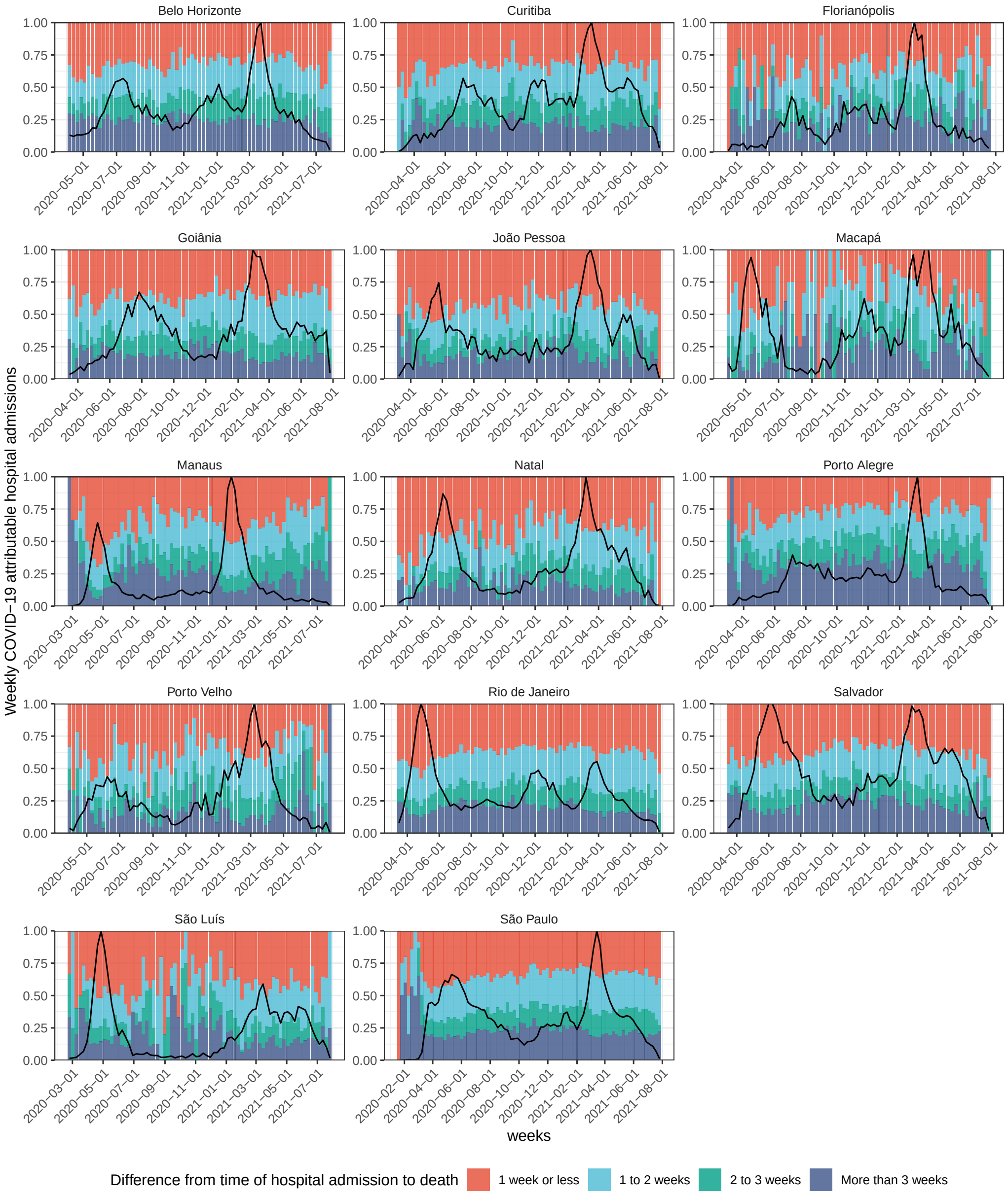

**Figure S40:** Proportion of COVID-19 attributable hospital admissions that died before 1, 2, 3 weeks or after 3 weeks since admissions date (colors). For reference, the time evolution of weekly hospital admissions is shown with the black line.
